# Home-use Photobiomodulation Device Treatment Outcomes for COVID-19

**DOI:** 10.1101/2022.06.16.22276503

**Authors:** Lew Lim, Nazanin Hosseinkhah, Mark V. Buskirk, Andrea Berk, Genane Loheswaran, Zara Abbaspour, Mahta Karimpoor, Alison Smith, Yoke N. Au, Kai F Ho, Abhiram Pushparaj, Michael Zahavi, Alexander White, Jonathan Rubine, Brian Zidel, Christopher Henderson, Russell G. Clayton, David R. Tingley, David J. Miller, Mahroo Karimpoor, Michael R. Hamblin

## Abstract

**BACKGROUND:** There is need for non-pharmaceutical treatments for COVID-19. A home-use photobiomodulation (PBM) device was tested as Treatment in a randomized clinical trial.

**METHODS:** 294 patients were randomized with equal allocation to Treatment or Standard of Care (Control). 199 qualified for efficacy analyses. The Treatment group self-treated for 20 minutes twice daily, for the first 5 days, and subsequently once daily for 30 days. A validated respiratory questionnaire was used, and patients were monitored remotely. The primary endpoint was the time-to-recovery (3 consecutive days of no sickness) for general sickness. The Kaplan-Meier method and the Cox Proportional Hazards model were primary methods of analyses.

**RESULTS:** Treatment patients with collective 0-12 days of symptoms, at moderate-to-severe level on Day 1 of Treatment, did not recover significantly faster than Control. However, for patients with 0-7 days of symptoms there was a significant mean difference of 3 days: Treatment, 18 days (95% CI, 13-20) vs. Control, 21 days (95% CI, 15-28), P=0.050. The Treatment:Control hazard ratio at 1.495 (95% CI, 0.996-2.243), P=0.054 exceeded the pre-trial target of 1.44. Treated patients exceeding 7 days symptoms duration were more tired and had lower energy. None of the patients in the Treatment group suffered death or hospitalization while the Control group had 1 death and 3 severe adverse events requiring hospitalization.

**CONCLUSIONS:** Patients with up to 7 days of symptoms at moderate-to-severe levels on first day of Treatment can expect faster recovery for general sickness and several respiratory symptoms. (Funded by Vielight Inc.; ClinicalTrials.gov number, NCT04418505.)

## INTRODUCTION

The National Institutes of Health reported that some people have sought “alternative” remedies to treat COVID-19^1^, also supported by other reports^2^; hence a need to consider devices for treatment. We report on a randomized clinical trial (RCT) of a non-pharmaceutical option to treat COVID-19, a home-use device based on photobiomodulation (PBM).

The PBM device delivers red and near infrared (NIR) light to selected areas of the body, stimulating mitochondrial activity^3^. The mechanisms include the release of nitric oxide (NO) in the mitochondria^4 3^ which has been shown to inhibit the replication of exposed coronavirus^5 6 7 8 9^ and support endothelial function^10 11^, beneficial to patients with acute respiratory distress syndrome (ARDS) and impaired pulmonary function^12 13 14^, which are features of acute COVID-19^15 16 17^.

PBM may attenuate inflammation observed in cases of COVID-19^18 19 9^. NIR light may reach damaged lungs to accelerate healing^20^. The elevated cell count in bronchoalveolar lavage, inflammatory cytokines and neutrophil numbers were reduced in PBM experiments^21^. Systematic reviews^22 23 24 25 26^ and case reports^27 28 29 30 31^ warrant this RCT.

See Appendix 2, Supplement 1.2.

## METHODS

### TRIAL DESIGN

The Control group received standard of care, whereas the Treatment group added self-administered home treatment with a PBM device, the “Vielight RX Plus”. The trial complied with the Declaration of Helsinki and the International Conference on Harmonization Guidelines for Good Clinical Practice. The protocol was approved by Health Canada and an institutional review board. Patients provided signed informed consent before enrollment. Information was posted on NIH National Library of Medicine website (ClinicalTrials.gov Identifier: NCT04418505).

The measures of COVID-19 improvement were based on the response to relevant questions (Q)1 through 43 on the Wisconsin Upper Respiratory Symptom Survey (WURSS)-44, scoring from 0 (not sick) to maximum 7 (severely sick) (Appendix 1 of the Protocol in Supplement 1).

Patients uploaded answers daily through the REDCap Cloud electronic data capture (EDC) platform over 30 days.

### PATIENTS AND PROCEDURES

All patients had tested positive for severe acute respiratory syndrome coronavirus 2 (SARS-CoV-2) infection with reverse transcriptase–polymerase chain reaction (PCR) tests. Qualifying patients scored 4-7 on WURSS Q1 on Day 1 of treatment. Patients were registered via EDC software and then randomized with equal allocation to the Treatment or Control group using the OxMAR minimization software^32^ (See Figure).

**Figure.**
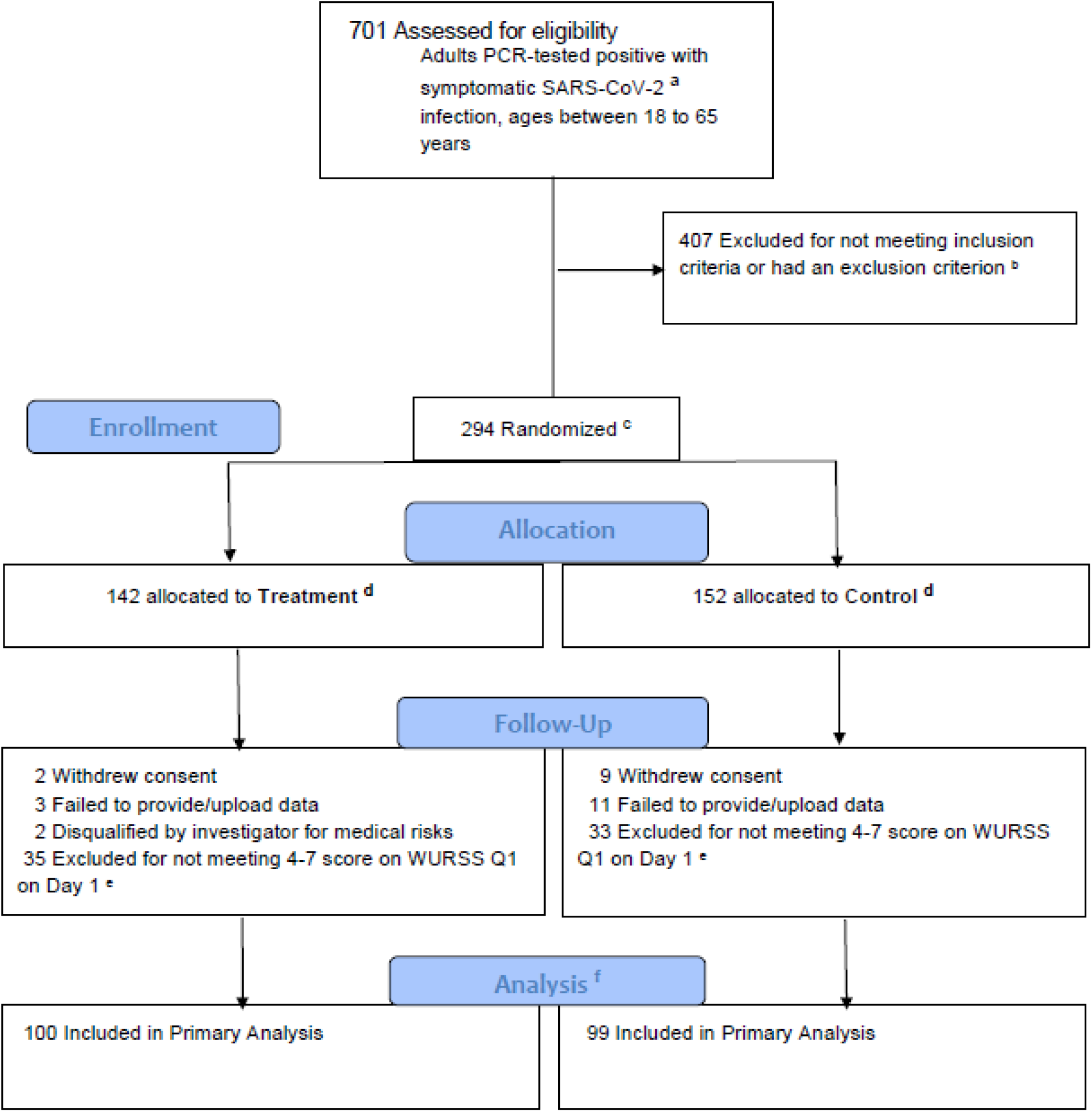
Patient Enrollment and Treatment Allocations. a. SARS-CoV-2 indicates severe acute respiratory syndrome coronavirus 2. b. The list of inclusion and exclusion criteria are presented in Sections 6.2 and 6.3 of the Protocol in Supplement 1. c. For enrollment and randomization, patients met the inclusion criteria, which included scores of 4-7 on the WURSS-44 Q1. Patients were allocated equally to Treatment or Control. d. Treatment involved following the standard of care (SoC) plus use of the Vielight RX Plus device, while Control only involved SoC. The allocations to Treatment and Control were managed by the OxMAR randomization software. e. Between Enrollment and Baseline (Day 1 of Treatment), shipping added a mean of 2 days before “Day 1”. 35 patients in Treatment and 33 in Control improved to the point that they no longer scored 4-7 for WURSS-44 Q1 and hence excluded from Baseline for analyses. However, they remained for safety monitoring with no bearing on efficacy. f. Primary analyses were carried out on patients who had 4-7 on WURSS-44 Q1 on Day 1 of Treatment (Baseline). The primary time-to-event analyses along with the “intention-to-treat” started at this point.

### TRIAL INTERVENTION AND MONITORING

The intervention was the “Vielight RX Plus” device, shipped to Treatment patients within 24 hours of randomization. The Treatment was self-administered for 20 minutes twice a day for the first 5 days, and subsequently once daily. A pulse oximeter was shipped to all patients to measure oxygen saturation. See Section 5.9 of Supplement 1, product specifications in Supplement 2.

This trial was monitored remotely by a contract research organization, principal investigators, qualified investigators, and study staff.

### EFFICACY OUTCOMES

The primary efficacy outcome was the time-to-recovery (days) for WURSS-44 Q1, “How sick do you feel today?”. Recovery was defined as the first day of 3 consecutive days with 0 (not sick) score.

Secondary efficacy outcomes included time-to-recovery (days) for WURSS-44 Q2-Q43, and number of days with mild symptoms (0-3 scores). For Safety assessments, the number and percentages of patients reporting adverse events (AEs) and daily oxygen saturation with pulse oximetry were reported. See details in Section 7 of Supplement 1.

The trial targeted to enroll 280 patients in 1:1 randomization. The study was designed to detect the minimum Treatment:Control hazard ratio (HR) of 1.44, with approximately 80% power with 5% type 1 error.

### STATISTICAL METHODS

Time-to-recovery (days) was estimated by the Kaplan-Meier (KM) method^33^ overall and for baseline strata of 0-5 days and 6-10 days symptoms duration established on enrollment. A stratified log-rank test compared the outcome distributions between Treatment and Control by symptoms duration. An unstratified KM method and log-rank test were used to evaluate time-to-recovery over strata with terms for treatment and symptoms duration strata. Subjects who did not recover were censored on Day 30.

Supportive analyses included stratified and unstratified Cox Proportional Hazards models^34^ with 95% confidence intervals (CI).

An analysis of variance (ANOVA) was used to compare mean days of mild symptoms with terms for treatment and symptoms strata. Unstratified analyses were by ANOVA with Treatment as the explanatory variable.

A linear mixed model repeated measures analysis of covariance^35^ was used to compare percentage changes in oxygen saturation in safety monitoring. Model terms included treatment, days (7, 14, 21, 28), symptom days strata treatment-by-day interaction and baseline covariate.

Frequency distributions of adverse events were presented. A Poisson regression model was used to compare the mean number of episodes of adverse event (AE) and patients with AEs, between Treatment and Control^36^.

The Statistical Analysis Plan and statistical methods are discussed in Supplement 3.1-3.3.

An interim analysis was conducted in January 2021. The results from 73 patients indicated that the study should continue and not stop for futility nor superiority (Supplements 5.1-5.2).

Missing data were not imputed. Kaplan-Meier estimates account for variable follow-up time under the assumption of non-informative censoring.

SAS software^37^ was used for statistical analysis. All P-values were two-sided and a P-value <0.050 was used to declare statistical significance.

Sensitivity analysis by multiple imputation was not performed as the data were uniformly complete.

## Results

### PATIENTS

Recruitment started in September 2020 and data collection completed in August 2021. 701 adults who tested positive for COVID-19 were assessed for eligibility. 407 failed the initial inclusion/exclusion criteria, leaving 294 patients for enrollment, randomization, and allocation at screening. For efficacy analysis, Baseline (“Day 1”) was established as the day of first-use of the Treatment device. Shipping added a mean of 2 days, extending the 0-5 days stratum to 0-7 days and 6-10 days stratum to 8-12 days. During this interval, 35 in Treatment and 33 in Control improved and scored below 4 on WURSS-44. Baseline with the intention-to-treat was then established with 199 patients (100 Treatment and 99 Control). See Figure, details in Supplements 4.1-4.3.

Patient demographics, baseline characteristics and WURSS-44 severity scores are presented in Table 1.

**Table 1.** Patient Demographics, Baseline Characteristics and Symptom Severity Scores.

| Characteristic | N |  |  |
| --- | --- | --- | --- |
|  | Treatment | Control | Combined |
|  | 100 | 99 | 199 |
| Age |  |  |  |
| Mean (SD) | 37.9 (13.16) | 35.3 (11.82) | 36.6 (12.54) |
| Median (IQR) | 37 (22.4) | 34 (18.7) | 36 (20.7) |
| Sex N (%) |  |  |  |
| Female | 31 (31.0) | 32 (32.3) | 63 (31.7) |
| Male | 68 (68.0) | 67 (67.7) | 135 (67.8) |
| Unstated | 1 (1.0) | 0 | 1 (0.5) |
| Ethnicity N (%) |  |  |  |
| American Indian / Alaskan | 2 (2.0) | 0 | 2 (1.0) |
| Black | 2 (2.0) | 5 (5.1) | 7 (3.5) |
| Hawaiian / Islander | 4 (4.0) | 4 (4.0) | 8 (4.0) |
| Caucasian | 82 (82.8) | 78 (78.8) | 160 (80.8) |
| Other | 9 (9.1) | 12 (12.1) | 21 (10.6) |
| Anthropometrics |  |  |  |
| Height (inches) |  |  |  |
| Mean (SD) | 66.7 (3.63) | 66.6 (4.23) | 66.7 (3.93) |
| Median (IQR) | 66.0 (5.0) | 66.0 (7.0) | 66.0 (5.0) |
| Weight (pounds) |  |  |  |
| Mean (SD) | 181.4 (54.24) | 173.0 (43.71) | 177.2 (49.31) |
| Median (IQR) | 172.0 (69.0) | 165.0 (55.0) | 170.0 (60.0) |
| BMI (kg/m<sup>2</sup>)<sup>a</sup> |  |  |  |
| Mean (SD) | 28.7 (9.26) | 27.3 (6.37) | 28.0 (7.39) |
| Median (IQR) | 27.0 (9.2) | 26.2 (8.2) | 26.6 (8.6) |
| Symptoms Duration Stratum at Baseline (%)<sup>b</sup> |  |  |  |
| 0-5 | 68 (68.0) | 68 (68.7) | 136 (68.3) |
| 6-10 | 32 (32.0) | 31 (31.3) | 63 (31.7) |
| 0-10 Combined Total | 100 (100.0) | 99 (100.0) | 199 (100.0) |
| WURSS-44 Severity Score at Baseline N (%)<sup>c</sup> |  |  |  |
| 4 | 32 (32.0) | 32 (31.3) | 63 (31.7) |
| 5 | 51 (51.0) | 48 (48.5) | 99 (49.7) |
| 6 | 15 (15.0) | 17 (17.2) | 32 (16.1) |
| 7 | 2 (2.0) | 3 (3.0) | 5 (2.5) |
| Residency N (%) |  |  |  |
| Canada | 8 (8.0) | 5 (5.1) | 13 (6.5) |
| United States | 92 (92.0) | 94 (94.9) | 186 (93.5) |

Abbreviations: BMI, body mass index; SD, standard deviation, IQR, interquartile range; WURSS, Wisconsin Upper Respiratory Symptoms Survey.
*a* Calculated as weight in kilograms divided by height in meters squared.
*b* Patients were stratified by symptoms duration strata 0-5 days or 6-10 days of having COVID-19 symptoms (established upon Enrollment).
*c* Qualifying patients at Baseline had WURSS-44 Q1 scores of 4-7.

### PRIMARY EFFICACY OUTCOMES

For the 199 patients at Baseline (0-10 days symptoms duration), the median time-to-recovery for Treatment was 19 days (95% CI, 16-22) vs. Control of 21 days (95% CI, 19-25), P=0.197, a median difference of 2 days.

For the 0-5 days symptoms duration stratum, the median time-to-recovery for Treatment was 18 days (95% CI, 13-20) vs. Control of 21 days (95% CI, 15-28), P=0.050, a median difference of 3 days.

For the 6-10 days stratum, the median time-to-recovery for Treatment was 23 days (95% CI, 19-27) vs. Control of 21 days (95% CI, 15-23), a median difference of -2 days (P=0.507).

In summary, the 0-5 days stratum demonstrated a significant (P=0.050) 3 day improvement to recovery. The 6-10 days stratum was worse (slower recovery) by 2 days (P=0.507).

Results are in Table 2, full table in Supplement 6.3.

**Table 2.**
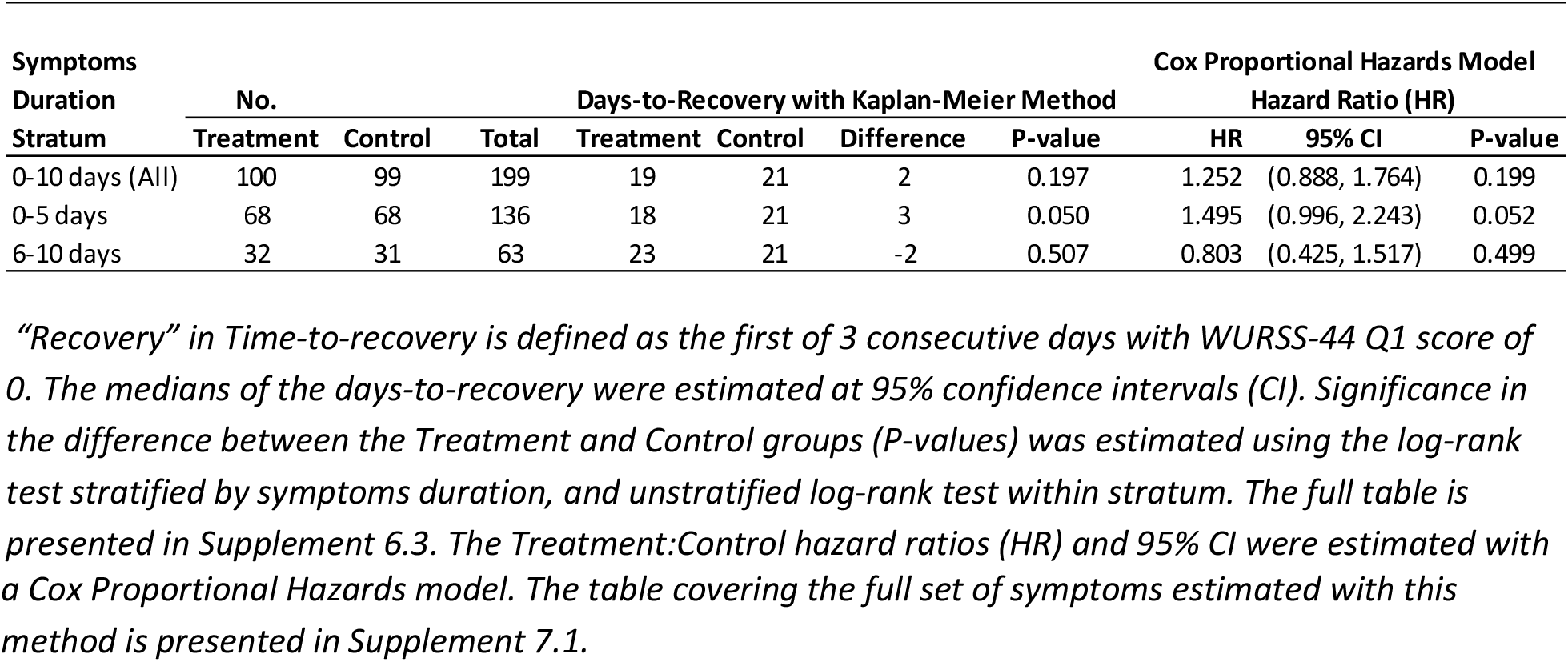
Primary Outcome – Time-to-Recovery and Hazard Ratio by Symptoms Duration Strata.

For all patients on Day 1, the hazard ratio (HR) was 1.252 (95% CI, 0,888-1.764), P=0.199. For the 0-5 days stratum, HR was 1.495 (95% CI, 0.996-2.243), P=0.052. For the 6-10 days stratum, HR was 0.803 (95% CI, 0.425-1.517), P=0.499. See Table 2 and Supplement 7.1. The HR of 1.495 for the 0-5 days stratum exceeded the pre-trial target, while the 6-10 days stratum and full population did not.

### SECONDARY EFFICACY OUTCOMES

As secondary efficacy outcomes, patients with WURSS-44 Q1 scores of 4-7 at Baseline were assessed for time-to-recovery for Q2-Q43 (Table 3).

**Table 3.**
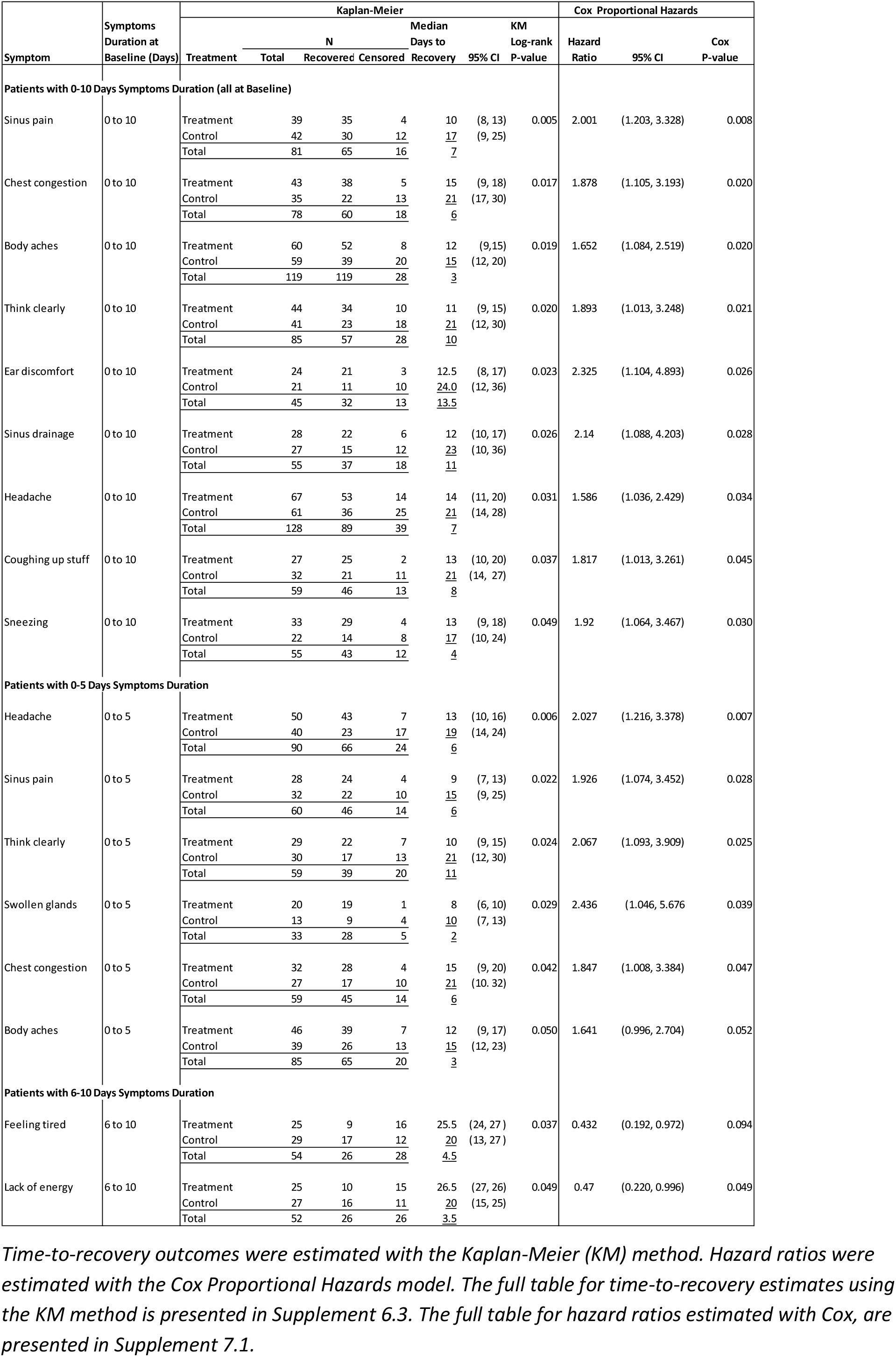
Secondary Outcomes of Patients with P<0.050 Log-rank for Time-to-recovery and Hazard Ratio.

For the full population, statistical significance favoring Treatment was observed for sinus pain, chest congestion, body aches, think clearly, ear discomfort, sinus drainage, headache, coughing up stuff and sneezing.

For the 0-5 days symptoms duration stratum, significance was observed for headache, sinus pain, thinking clearly, chest congestion and body aches.

For the 6–10 days stratum, the Treatment group recovered significantly more slowly for feeling tired and lack of energy.

We also assessed the Treatment effectiveness in reducing symptom severity, expressed as the mean number of days of mild symptoms (WURSS-44 Q1-43 scores of 0-3). For all patients, Treatment results were significantly better for runny nose, sneezing, body aches, irritability, and ear discomfort. For the 0-5 days stratum, Treatment showed significance for headache. For the 6-10 days stratum, Treatment showed significance for runny nose, sneezing, body aches, sinus drainage and plugged ears. See Supplement 8.

### SAFETY OUTCOMES

After initial follow-up, 267 enrolled patients (135 in Treatment and 132 in Control) were monitored for adverse events. The safety population included all randomized subjects with a response of 4+ to WURSS Q1 on enrollment.

None of the Treatment patients suffered death or severe adverse events (SAEs). In Control, there were 4 (3.0%) SAEs that required hospitalization, including 1 death (Tables 4(1)-4(2)).

**Table 4.**
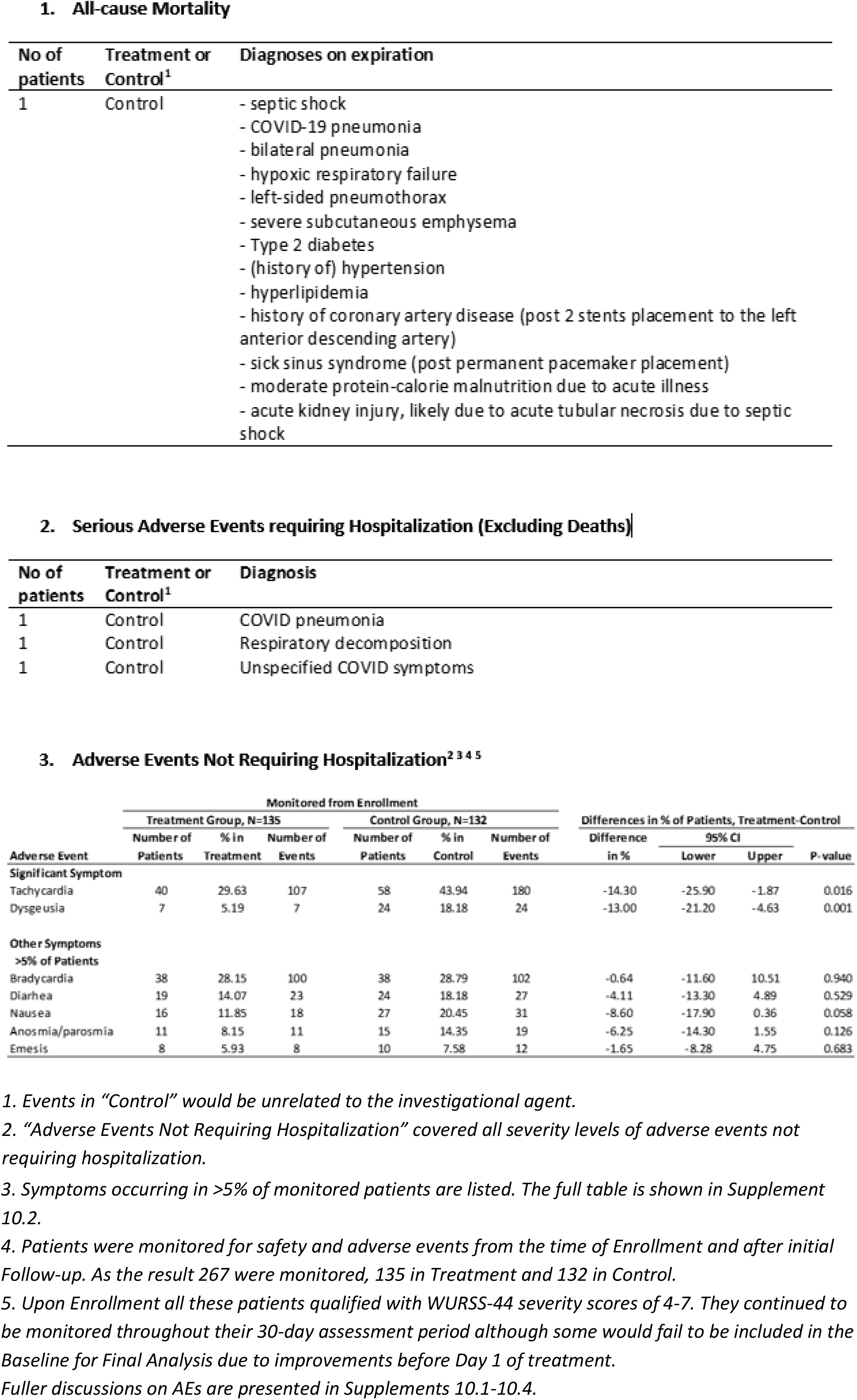
Summary Tables of Adverse Events.

AEs occurring in >5% of patients are listed in Table 4(3). Patients in Treatment had significantly lower AEs in Tachycardia and Dysgeusia but not for other AEs.

In the assessment of percentage changes in oxygen saturation, Treatment produced improvements with a mean difference of 0.32%, P=0.018 (Supplements 9.1-9.3).

## Discussion

The assessment of patients with combined symptoms duration of 0-10 days (established on enrollment) did not show significance for the primary outcome of time-to-recovery for general sickness. However, those with 0-5 days presented significant Treatment vs Control difference with P=0.050, supported by hazard ratios exceeding the pre-trial target.

The strata of 0-5 days and 6-10 days symptoms duration were reset to 0-7 days and 8-12 days respectively at Baseline due to device shipment time, allowing the same start for Control and Treatment patients for analyses.

By interpretation, patients with symptoms of up to 7 days can expect to recover more quickly than those with longer symptoms duration; and avoid the side effects of tiredness and energy deficits.

Patients with 0-7 days symptoms duration are also more likely to experience quicker recovery for headache, sinus pain, think clearly, swollen glands, and chest congestion; and experience more mild days with headache.

Fewer treated patients are expected to experience tachycardia and ageusia which were the most frequent adverse events reported.

There were several limitations. Firstly, the RCT was not double-blinded with a placebo device. Attempts at masking the efficacious visible red light of this device would likely fail with alert users. Secondly, the methodology was based on self-reporting. However, the WURSS-44 questionnaire had performed well as an illness-specific quality-of-life evaluative outcome instrument.^38^ Thirdly, the statistical power to detect differences in each WURSS-44 Q1-Q43 was reduced due to the sample size presenting with WURSS-44 4-7 severity scores for each item at Baseline.

## Supporting information

Supplementary Table of Contents

Protocol

Treatment Device Specifications

Statistical Analysis Plan and Methods

Patient Demographics, Information

Interim Analysis

Kaplan-Meier Method Results

Cox Proportional Hazards Estimates

Full Table ANOVA Mild Symptoms

Oxygen Saturation Analysis

Adverse Events

## Data Availability

All data produced in the present study are available upon reasonable request to the authors.

## Notes

### Competing Interest Statement

Lew Lim: Employee of sponsor, Vielight Inc.,
Nazanin Hosseinkhah: Employee of sponsor, Vielight Inc.,
Mark Van Buskirk: Paid consultant for the study,
Andrea Berk: Paid consultant for the study,
Genane Loheswaran: Employee of sponsor, Vielight Inc.,
Zara Loheswaran: Employee of sponsor, Vielight Inc.,
Alison Smith: Ex-employee of sponsor, Vielight Inc., was a co-investigator in the study,
Mahta Karimpoor: Ex-employee of sponsor, Vielight Inc., was a co-investigator in the study,
Yoke N Au: Employee of sponsor, Vielight Inc.,
Kai F Ho: Paid consultant for the study,
Abhiram Pushparaj: Paid consultant for the study,
Michael Zahavi: Paid investigator for the study,
Alexander White: Paid investigator for the study,
Jonathan Rubine: Paid investigator for the study,
Brian Zidel: Paid investigator for the study,
Christopher Henderson: Paid consultant for the study,
Russell G Clayton Sr: Paid investigator for the study,
David R Tingley: Paid investigator for the study,
David J Miller: Paid investigator for the study,
Mahroo Karimpoor: Employee of sponsor, Vielight Inc.,
Michael R Hamblin: Paid advisor to sponsor, Vielight Inc.

### Clinical Trial

NCT04418505

### Funding Statement

This study was funded by Vielight Inc.

### Author Declarations

Ethical approval granted by IRB: Advarra, Inc., and its affiliated entities, with a principal place of business at 6940 Columbia Gateway Drive, Suite 110, Columbia, MD 21046.

