## Supplementary Table of Contents for "Home-use Photobiomodulation Device Treatment Outcomes for COVID-19"

**SUPPLEMENTARY MATERIAL**

**Table of Contents**

Supplement 1: Protocol

1.1 Notes to Clinical Trial Protocol

1.2 Clinical Trial Protocol

1.3 Table of Amendments to Protocol

Supplement 2: Device Specifications

Treatment Device, Vielight RX Plus Specifications

Supplement 3: Statistical Analysis Plan and Statistical Methods

3.1 Notes to the Statistical Analysis Plan

3.2 Statistical Analysis Plan

3.3 Table of Amendments to Statistical Analysis Plan

3.4 Statistical Methods Used

Supplement 4: Patient Information

4.1 Chronology, Recruitment Numbers and Baseline Discussion

4.2 Full Table of Patient Demographics and Baseline Characteristics

4.3 Demographics and Baseline Characteristics of Patients with Baseline WURSS-44 Q1 Score of 4-7 and Summary Statistics for Days between Enrollment and Baseline

Supplement 5: Interim Analysis

5.1 Interim Analysis Report

5.2 Interim Efficacy Analysis Methods

Supplement 6: Kaplan-Meier Method

6.1 Introduction to the Kaplan-Meier Method Used

6.2 Outcomes Affecting >5% of Patients for Time-to-recovery and Hazard Ratio (P<0.050)

6.3 Full Table for Kaplan-Meier Estimates in Patients with a WURSS-44 Q1 Score of 4-7 at Baseline

6.4 Kaplan-Meier Curves of Primary Outcome

6.5 Kaplan-Meier Curves for Secondary Symptoms with Duration 0-5 days and 6-10 days at Screening (P≤0.050)

Supplement 7: Cox Proportional Hazards Estimates

7.1 Full Table for Cox Proportional Hazards Estimates

7.2 Patient Demographics and Baseline Characteristics Response to Treatment - Univariate Cox Proportional Hazards Model in Symptom Duration 0-5 Days at Screening with Baseline Score of 4-7 for WURSS Question 1

Supplement 8: Mild Symptoms Analysis of Variance

Full Table of Analysis of Variance for Days of Mild Symptoms - Comparison of Days with WURSS-44 Scores of 0-3 for All Symptoms in Patients with Baseline WURSS-44 Q1 Score of 4-7

Supplement 9: Oxygen Saturation Analysis

9.1 Summary of Oxygen Saturation Analysis

9.2 Mixed Model Repeated Measures Analysis of Covariance Treatment Comparison of Oxygen Saturation (%) in in All Patients with Oxygen Saturation Data from Enrollment

9.3 Chart of Change in Oxygen Levels with Standard Error Bars

Supplement 10: Adverse Events

10.1 Notes on Adverse Events

10.2 Non-severe Adverse Events Affecting More than 5% of Patients

10.3 Severe Adverse Events Report by Medical Monitor
