## Supplementary material for "Home-use Photobiomodulation Device Treatment Outcomes for COVID-19": Protocol

### Supplement 1:
### Protocol

**Table of Contents**

1.1 Notes to Protocol

1.2 Clinical Trial Protocol

1.3 Table of Amendments to Protocol

#### Supplement 1.1: Notes to Protocol

The clinical trial was conducted using version 4 of the Clinical Trial Protocol (Supplement 1.2) updated with amendments as tabled in this supplement (Supplement 1.3). Amendments to the original Protocol leading to version 4 had been recorded on pages 1 to 8 of the version 4 Protocol.

The amendments between version 1 to version 4 were summarized as follows:

- 2020-05-13 Initial protocol (April 2020)
- 2020-05-19 Protocol revised to elaborate on the role of Qualified Investigator along with the Principal Investigator
- 2020/08/05 Protocol revised to meet US regulatory requirements and include US-site Clinical Protocols (per 21 CFR 803 and 812)
- 2020/12/10 Protocol revised to add an exclusion criterion and add interim analysis

The selection of the few secondary efficacy endpoint items was made during early 2020 out of best-guess when little was known about COVID-19, and based on the false premise that it was necessary to restrict analysis to a few symptoms. It became apparent that there was no productive advantage in this narrow scope and far more useful to cover all the items on the WURSS-44 questionnaire; hence the amendments.

These would flow through to affect the Statistical Analysis Plan with similar amendments.

The primary endpoint was not affected.

**VIELIGHT RX PLUS FOR COVID-19**

### A Randomized Study Evaluating the Efficacy of the Vielight RX Plus in the Treatment of COVID-19 Respiratory Symptoms

---

Protocol No:

Revision date: 2020-12-10 V4.0

Study Products: Vielight RX Plus

**SPONSOR: Vielight Inc.**

**Contact: Lew Lim, PhD, MBA**

**346A Jarvis St.**

**Toronto, Ontario**

**M4Y 2G6, Canada.**

**1-855-875-6841**

****

***CONFIDENTIAL:*** *This protocol, its contents, and the information relating to it are the property of Vielight Inc. All information is to be kept confidential.*

#### REVISION HISTORY

2020-05-13 Initial protocol drafted by Vielight Research Team (April 2020)

2020-05-19 Protocol revised to elaborate on the role of Qualified Investigator along with the Principal Investigator

2020/08/05 Protocol revised to meet US regulatory requirements and include US-site Clinical Protocols (per 21 CFR 803 and 812)

2020/12/10 Protocol revised to add an exclusion criterion and add interim analysis

©2020 Vielight Inc.
All rights reserved

**Summary of changes, Protocol version V4.0**

| Item (Section) | Original Text | Revised Text | Rational for Change |
|---|---|---|---|
| Table of contents | | | Updated |
| 1.0 Protocol Summary | Qualified Investigator (QI) | Study Investigators | To generalize the role for US site addition |
| 1.0 Protocol Summary | | Participants within the same household | To exclude the same household participants to avoid switching treatments and contaminating the results |
| 2.2 COVID-19 Standard of Care Applied During Study | | The U.S. Government provides an Interim Clinical Guidance for Management of Patients with Confirmed Coronavirus Disease (COVID-19) by Centers for Disease Control and Prevention as an agency of the U.S. Department of Health and Human Services (Centers for Disease Control and Prevention, 2020a, 2020b). | The US Interim Clinical Guidance was added for US site addition |
| 3.1 Study Objective | Qualified Investigator (QI) | Investigators | To generalize the role for US site addition |
| 3.2 Study Design Rationale | (Centers for Disease and Control, 2020) | (Centers for Disease and Control, 2020c) | Reference updated |
| 4.0 Overview of Study Design | Qualified Investigator (QI) | Qualified Investigator (QI) in Canada and a Principal Investigator (PI) in the United States. | To generalize the role for US site addition |
| 5.2 Electronic Informed Consent Form | Qualified Investigator | -Investigators | To generalize the role for |

©2020 Vielight Inc.
All rights reserved

| Item (Section) | Original Text | Revised Text | Rational for Change |
|---|---|---|---|
| and Emergency Contact | | | US site addition |
| 5.4.1 Device Training | deal with potential issues followed by a comprehension test via the EDC | deal with potential issues. Instructional materials will also be included with each device shipment | Changed due to EDC capabilities |
| 5.4.1 Device Training | how to use the pulse oximeter will also be reviewed via the training module, followed by a comprehension test. | how to use the pulse oximeter will also be provided via an EDC module. | Changed due to EDC capabilities |
| 5.5 Baseline (Day 0) | Training comprehension test | Training review | |
| 5.5 Baseline (Day 0) | study investigators (i.e. QI, sub - investigators) via | study investigators | To generalize the role for US site addition |
| 8.6 Interim Analysis | | A single interim analysis will be conducted after 73 participants completed their study. To preserve the overall type I error rate at the nominal level of 0.05, the stopping boundary for efficacy will be set at $p<0.0051$ (O'Brien and Fleming, 1979). The stopping boundary for futility will be set at the conditional power of 50% (Lan et al, 1982). | To include an interim analysis at 73 participants |
| 9.0 CRO and Data Collection Methods | All study procedure | All study procedures | Typo |
| 10.1 Consent Process | Ethical Principles for Medical Research | Ethical Principles for Medical Research Involving Human Participants, and 21 CFR Part 50 Protection of Human Subjects. | To meet US regulatory requirements |

©2020 Vielight Inc.
All rights reserved

| Item (Section) | Original Text | Revised Text | Rational for Change |
|---|---|---|---|
| | Involving Human Participants. | | |
| 10.4 Study Suspension or Early Termination | | As defined in 21 CFR 813.3, an unanticipated adverse device effect means any serious adverse effect on health or safety or any life-threatening problem or death caused by, or associated with, a device, if that effect, problem, or death was not previously identified in nature, severity, or degree of incidence in the investigational plan or application (including a supplementary plan or application), or any other unanticipated serious problem associated with a device that relates to the rights, safety, or welfare of subjects.<br><br>As per 21 CFR 812.150, if Sponsor determines that an unanticipated adverse device effect presents an unreasonable risk to subjects, all investigations or parts of investigations presenting that risk shall be terminated, as soon as possible. Termination shall occur not later than 5 working days after Sponsor makes this determination and not later than 15 working days after the sponsor first received notice of the effect. The study shall not be resumed without FDA and IRB approval. | To meet US regulatory requirements |
| 12.1 Definition of Adverse Events | Investigators (which refers to the QI or designated co-investigators and study staff) | Investigators | To generalize the role for US site addition |
| 12.1 Definition of Adverse Events | | An AE is defined as being serious (SAE) if it results in death or meets the definition of serious injury, as defined in Title 21 of the Code of Federal Regulations, Section 803.3 (21 CFR | To meet US regulatory requirements |

| Item (Section) | Original Text | Revised Text | Rational for Change |
|---|---|---|---|
| | | 803.3), which means an injury or illness that:<br>i. Is life-threatening,<br>ii. Results in permanent impairment of a body function or permanent damage to a body structure; or<br>iii. Necessitates medical or surgical intervention to preclude permanent impairment of a body function or permanent damage to a body structure. Permanent means irreversible impairment or damage to a body structure or function, excluding trivial impairment or damage.<br><br>As defined in 21 CFR 803.3, caused or contributed means that the adverse device effect was or may have been attributed to the study device, or may have been a factor in in the event, including events occurring as a result of:<br><br>○ Failure,<br>○ Malfunction, | |

All rights reserved

| Item (Section) | Original Text | Revised Text | Rational for Change |
|---|---|---|---|
| | | ○ Improper or inadequate design,<br>○ Manufacture,<br>○ Labeling, or<br>○ User error. | |
| 12.3 Reporting Adverse Events and Device Incidents | The Sponsor will be responsible for submitting incident information to Health Canada within the specified timeframe.<br>The Investigator will report all of the above to the reviewing IRB (as applicable) according to their reporting requirements. Reports must identify subjects using the study's unique identifier to protect subject's confidentiality. | As per 21 CFR 812.150, this incident reporting shall not be later than 10 working days after the investigator first learns of the effect. The Investigator will report all of the above to the reviewing IRB (as applicable) according to their reporting requirements. Reports must identify subjects using the study's unique identifier to protect subject's confidentiality.<br><br>The Sponsor will be responsible for submitting incident information to Health Canada and FDA within the specified timeframe. As per 21 CFR 812.46, Sponsor shall immediately conduct an evaluation of any unanticipated adverse device effect. Sponsor shall report the results of such evaluation to FDA and to the reviewing IRB and participating investigators within 10 working days after receiving notice of the effect. Thereafter the sponsor shall submit such additional reports concerning the effect as FDA requests. | To meet US regulatory requirements |
| 12.6 Incidental Findings | QI | QI/PI | To generalize the role for US site addition |
| 12.7 Protocol Deviations | QI | QI/PI | To generalize the role for |

All rights reserved

| Item (Section) | Original Text | Revised Text | Rational for Change |
|---|---|---|---|
| | | | US site addition |
| 12.7 Protocol Deviations | Qualified Investigator. | QI/PI. | To generalize the role for US site addition |
| 13.2 Investigator Responsibilities | applicable Health Canada regulations, | applicable Health Canada regulations, applicable FDA regulations, | To meet US regulatory requirements |
| 13.3 Required Documents from the Investigators | QI Investigators | Investigators | To generalize the role for US site addition |
| 13.4 Investigator Records | whichever date is later. | whichever date is later, per 21 CFR 812.140(d). | To meet US regulatory requirements |
| 13.4 Investigator Records | | Records must be retained in designated administrative files at each Investigational Site and/or retained by the study sponsor<br>○ Trial protocol and all amendments<br>○ Ethics Committee Approval Letter(s) and approved informed consent(s) (including any revisions)<br>○ Device Instructions for Use<br>○ CVs of all investigators<br>○ Site Subject Log<br>○ Monitoring reports<br>○ Site Authorized Personnel Signature List | To meet US regulatory requirements |

©2020 Vielight Inc.
All rights reserved

| Item (Section) | Original Text | Revised Text | Rational for Change |
|---|---|---|---|
| | | ○ Reports (includes Adverse Event reports and final reports from investigator and sponsor) | |
| 13.5 Specific Sponsor Responsibilities | qualified investigators | investigators that are qualified by training and education | Change is sentence structure |
| 13.8 Medical Monitoring | the qualified investigator | the investigator | To generalize the role for US site addition |
| 16.0 References | | Centers for Disease and Control (CDC). "What to Do If You Are Sick" Online: https://www.cdc.gov/coronavirus/2019-ncov/if-you-are-sick/steps-when-sick.html. Accessed. May 8, 2020b.<br><br>Centers for Disease and Control (CDC). "Discontinuation of Transmission-Based Precautions and Disposition of Patients with COVID-19 in Healthcare Settings (Interim Guidance)." Online: https://www.cdc.gov/coronavirus/2019-ncov/hcp/disposition-hospitalized-patients.html. Accessed. April 11, 2020c. | References to CDC were added to describe the SOC in US |

©2020 Vielight Inc.
All rights reserved

**Table of Contents**

| | | |
|---|---|---|
| 1.0 | Protocol Summary | 12 |
| 2.0 | Introduction | 16 |
| 2.1 | COVID-19 Pathology and Study Focus | 16 |
| 2.2 | COVID-19 Standard of Care Applied During Study | 16 |
| 2.3 | Vielight RX Plus Device Description | 17 |
| 2.3.1 | Design Principles | 17 |
| 2.3.2 | Regulatory Status | 17 |
| 2.4 | Mechanisms of Action of the Vielight RX Plus | 18 |
| 2.4.1 | Fundamental cellular mechanisms | 18 |
| 2.4.2 | Targeting the Thymus Gland, Sternal Bone Marrow and Lungs | 18 |
| 2.4.3 | Targeting the Respiratory Tract | 19 |
| 2.4.4 | Intranasal PBM on Respiratory Infections | 19 |
| 2.4.5 | The Systemic Effect of Irradiating the Free-Floating Mitochondria | 20 |
| 2.4.6 | Directly Inhibiting Coronavirus through Nitric Oxide Release | 20 |
| 2.4.7 | Direct Photobiomodulation of the Lower Respiratory Tract | 20 |
| 2.4.8 | Calming the Cytokine Storm | 20 |
| 2.5 | Clinical Data and Summary of Related Evidence to Date | 20 |
| 2.5.1 | Inhibition of coronavirus replication in the nasal passages | 21 |
| 2.5.2 | Development of T-lymphocytes | 21 |
| 2.5.3 | Attacking pathogens in the respiratory tracks, including the lungs | 21 |
| 2.5.4 | Production of nitric oxide in the respiratory tract tissues | 21 |
| 2.5.5 | Support recovery of damaged cells | 21 |
| 2.5.6 | Management of inflammation | 21 |
| 2.5.7 | Vielight devices have produced evidence of tissue responses | 21 |
| 2.5.8 | Track record of efficacy in other clinical studies | 21 |
| 3.0 | Study Objective and Rationale | 22 |
| 3.1 | Study Objective | 22 |
| 3.2 | Study Design Rationale | 22 |
| 4.0 | Overview of Study Design | 22 |
| 5.0 | Study Procedures | 23 |
| 5.2 | Electronic Informed Consent Form and Emergency Contact | 24 |
| 5.3 | Registration and Randomization | 24 |
| 5.4 | Shipment of Devices | 25 |

©2020 Vielight Inc.
All rights reserved

| | | |
|---|---|---|
| 5.5 | Baseline (Day 0) | 25 |
| 5.6 | Days 1 to 30 | 25 |
| 5.7 | Final, Day 30 | 26 |
| 5.8 | Subject Withdrawal | 26 |
| 5.9 | Treatment Protocol | 26 |
| 6.0 | Eligibility Criteria and Recruitment | 27 |
| 6.1 | Recruitment Strategy | 27 |
| 6.2 | Inclusion Criteria | 27 |
| 6.3 | Exclusion Criteria | 27 |
| 7.0 | Study Endpoints | 27 |
| 7.1 | Primary Endpoint | 27 |
| 7.2 | Secondary Endpoints | 27 |
| 8.0 | Statistical Methods | 29 |
| 8.1 | Sample Size Justification | 29 |
| 8.2 | Analysis Populations | 29 |
| 8.3 | Primary Endpoint: Recovery from 4-7 to 0 in WURSS-44 mean score | 29 |
| 8.4 | Secondary Endpoints | 29 |
| 8.4.1 | Outcomes based on mean number of days | 29 |
| 8.4.2 | Outcomes based on time-to-event | 30 |
| 8.4.3 | Descriptive Statistics | 30 |
| 8.5 | Assessment of Device Safety | 30 |
| 8.6 | Interim Analysis | 31 |
| 9.0 | CRO and Data Collection Methods | 31 |
| 9.1 | Study Duration | 31 |
| 10.0 | Ethical Considerations | 31 |
| 10.1 | Consent Process | 31 |
| 10.2 | Institutional Review Board (IRB) and Research Ethics Board (REB) Approvals | 31 |
| 10.3 | Data Protection | 31 |
| 10.4 | Study Suspension or Early Termination | 32 |
| 11.0 | Investigational Devices | 32 |
| 12.0 | Safety and Adverse Events | 33 |
| 12.1 | Definition of Adverse Events | 33 |
| 12.2 | Definition of Device Incident | 34 |
| 12.3 | Reporting Adverse Events and Device Incidents | 34 |

©2020 Vielight Inc.
All rights reserved

| | | |
|---|---|---|
| 12.4 | Adverse Event Follow-up | 35 |
| 12.5 | Anticipated Adverse Events | 35 |
| 12.6 | Incidental Findings | 35 |
| 12.7 | Protocol Deviations | 35 |
| 13.0 | Study Management | 36 |
| 13.1 | Sponsor Overall Responsibility | 36 |
| 13.2 | Investigator Responsibilities | 36 |
| 13.3 | Required Documents from the Investigator | 37 |
| 13.4 | Investigator Records | 37 |
| 13.5 | Specific Sponsor Responsibilities | 38 |
| 13.6 | Sponsor Records | 39 |
| 13.7 | Study Monitoring Plan | 39 |
| 13.10 | Protection of Subject Confidentiality | 40 |
| 13.11 | Quality Assurance and Supervision by Authorities and Privacy | 40 |
| 13.12 | Final Report | 40 |
| 13.13 | Information Confidentiality | 41 |
| 13.14 | Trial Registration | 41 |
| 14.0 | Study Close-out | 41 |
| 14.1 | Timeline of Close-out | 41 |
| 14.2 | Record Storage and Retention | 41 |
| 14.3 | Discontinuation of Study | 41 |
| 15.0 | Definitions and Acronyms | 41 |
| 16.0 | References | 45 |
| Appendix 1 – Wisconsin Upper Respiratory Symptom Survey (WURSS-44) | | 48 |
| Appendix 2 – White Paper | | 50 |

©2020 Vielight Inc.
All rights reserved

#### 1.0 Protocol Summary

| | |
|---|---|
| Study Title | **A Randomized Study Evaluating the Efficacy of the Vielight RX Plus in the Treatment of COVID-19 Respiratory Symptoms** |
| Methodology | Prospective, randomized trial |
| Study Duration | 30 days from the first use of the device. Duration of trial (from first subjects enrolled to completion of final participant) is expected to be approximately 6 months. |
| Study Site(s) | Participants are monitored remotely by the Contract Research Organization (CRO) and Study Investigators |
| Number of Subjects | 280 subjects (140 Standard of Care (SOC); 140 SOC + Vielight RX Plus) |
| Major Inclusion Criteria | • Confirmation of COVID-19 infection<br>• Score of 4-7 for WURSS-44 Question 1 (moderate to severe sickness)<br>• Between 18-65 years of age |
| Major Exclusion Criteria | • Need for hospitalization at the time of diagnosis<br>• Current need for supplemental oxygen or positive pressure support and/or has required supplemental oxygen or positive pressure support for >or= 24 hours<br>• > 10 days since symptom onset<br>• Diagnosis of Chronic Obstructive Pulmonary Disease (COPD)<br>• Pregnant<br>• Positive for Hepatitis C Virus (HCV), Hepatitis B Virus (HBV) or Human Immunodeficiency Virus<br>• Inability to electronically complete WURSS-44 daily (in English)<br>• Participants within the same household |
| Treatment | Vielight RX Plus administered for 20 minutes twice a day, separated by at least 6 hours for the first 5 days, and subsequently once daily, banded NIR LED module on the upper sternum and intranasal applicator inside the left or right nostril for the study duration |

©2020 Vielight Inc.
All rights reserved

| | |
|---|---|
| Study Design and Measures | • COVID-19 positive participants will be screened and consented through an electronic informed consent form (ICF) while they are at home in self-isolation via the electronic data capture system (EDC).<br>• Pre-existing health conditions, medication use, WURSS-44, Fitzpatrick scale, race/ ethnicity and sex, are recorded at baseline<br>• Participants will be randomized to SOC or SOC + Vielight RX Plus (Treatment) Groups using minimization software (OxMAR).<br>• Participants will complete an electronic shipping address form via the EDC at screening. Devices will be express couriered to participants; all participants will receive a pulse oximeter. Only participants in the Treatment group will receive a Vielight RX Plus.<br>• Participants will complete the self-report measure of symptoms (WURSS-44) daily via the EDC.<br>• Oxygen saturation levels at rest will be measured daily by participants and entered into the EDC.<br>• Participants will use a daily diary and checklist to record:<br> ○ treatment administration (compliance)<br> ○ complications related to the use of the device (e.g. nasal irritation)<br> ○ technical complications (e.g. device not turning on etc).<br> ○ change in medication or treatment<br> ○ emergent medical condition requiring medical attention (safety)<br> ○ hospitalization (safety)<br> ○ adverse events |
| Primary Endpoint | Time to overall recovery in days as measured by item 1 of the WURSS-44 “How sick do you feel today”. Recovery is defined as a rating of 0 (not sick) that is confirmed over 3 consecutive days. |

©2020 Vielight Inc.
All rights reserved

| | |
|---|---|
| Secondary Endpoints | A. Time to elimination of symptoms in days on the following items in the WURSS-44, where elimination of symptoms is defined as a rating of 0 (do not have this symptom or not at all) that is confirmed over 3 consecutive days:<br>o Item 2 “Cough”<br>o Item 12 “Body aches”<br>o Item 18 “Feeling tired”<br>o Item 37 “Breathe easily”<br>o Item 16 “Feeling feverish”<br>B. Mean number of days with a rating of 0, 1, 2 or 3 for the following items:<br>o Item 1 “How sick do you feel today”<br>o Item 2 “Cough”<br>o Item 12 “Body aches”<br>o Item 18 “Feeling tired”<br>o Item 37 “Breathe easily”<br>o Item 16 “Feeling feverish”<br>C. Mean number of days with mild overall respiratory symptoms as measured by a total WURSS-44 score of <=129<br>D. Time to reduction in symptoms in days as measured by item 1 of the WURSS-44 “How sick do you feel today”. Reduction in symptoms is defined as a rating of 3 or less that is confirmed over 3 days.<br>E. Time to elimination of symptoms in days on the following items in the WURSS-44, where elimination of symptoms is defined as a rating of 0 (do not have this symptom or not at all) that is confirmed over 3 consecutive days:<br>o Item 16 “Feeling feverish”<br>o Item 37 “Breathe easily”<br>F. Average number of days spent with mild respiratory symptoms for the 7 questions relating to respiratory symptoms defined below (WURSS-44 respiratory score < or = 21) in treatment group compared to SOC.<br>WURSS-44 rating on past 24hrs for<br>o ‘cough’,<br>o ‘coughing stuff up’,<br>o ‘cough interfering with sleep’,<br>o ‘chest congestion’,<br>o ‘chest tightness’,<br>o ‘heaviness in chest’<br>o ‘breathe easily’<br>G. Oxygen Saturation by Pulse Oximetry [Time Frame: 30 Days]<br>H. Proportion of subjects with serious adverse events in treatment group compared to SOC.<br>I. Mortality rate in Treatment group compared to SOC |

©2020 Vielight Inc.
All rights reserved

| | |
|---|---|
| Statistical Methodology | **Time-to-event outcomes**<br>Time-to-event is defined as the interval from the date of first use of the device to the start date of the improvement. Where a durable improvement over 3 days is indicated, the start date of the improvement will be taken to be the first day where the criteria is met. Subjects who have not met the definition of improvement will be censored as of their last available assessment. Subjects without a baseline assessment or post-baseline assessment will be censored at the date of randomization.<br>Time-to-improvement will be estimated using Kaplan-Meier methods. The proportion of subjects with an improvement will be reported by treatment group. The overall group comparison will be done using a log-rank test stratified by time since onset of symptoms (0-5 days vs. 5-10 days). The hazard ratio and its associated 95% confidence interval will be calculated based on a stratified Cox proportional hazards model.<br>The study will enroll 280 patients in a 1:1 randomization and the primary analysis will be performed when all patients have completed treatment, regardless of the number of recovery events observed. If, as expected, nearly all patients should recover within the 30-day treatment period, then a hazard ratio of 1.40 can be detected with approximately 80% power with a two-sided significance level of 5%. If a substantial proportion of patients (i.e. approximately 15%) do not record a recovery within the 30-day treatment period, then a hazard ratio of 1.44 can be detected with approximately 80% power with a two-sided significance level of 5%. |

©2020 Vielight Inc.
All rights reserved

#### 2.0 Introduction

##### 2.1 COVID-19 Pathology and Study Focus

COVID-19 is caused by the novel coronavirus, severe acute respiratory syndrome coronavirus 2 (SARS-CoV-2). The pathogenesis of COVID-19 is not fully understood. Some people infected with the virus have no symptoms. When the virus does cause symptoms, common ones include fever, dry cough, fatigue, loss of appetite, loss of smell, and body ache. In some people, COVID-19 causes more severe symptoms like high fever, severe cough, and shortness of breath, which often indicates pneumonia (Harvard, 2020). Experience in China found that approximately 14% developed severe disease requiring hospitalization and oxygen support and 5% required admission to an intensive care unit (ICU) (Team NCPERE, 2020). In severe cases, COVID-19 can be complicated by acute respiratory distress syndrome (ARDS); sepsis and septic shock; and multiorgan failure, including acute kidney injury and cardiac injury (Yang et al, 2020). However, respiratory failure is the most common factor and main cause of fatality (Ruan et al, 2020). For this reason, this clinical study will focus on respiratory-related symptoms.

##### 2.2 COVID-19 Standard of Care Applied During Study

The Canadian Government provides an Interim Guidance for clinical management of patients with moderate to severe COVID-19 (Government of Canada, 2020), which will serve as the reference for Standard of Care (SOC) adopted in this study. The Interim Guidance calls for patients with the abovementioned symptoms to be screened and triaged, the first point of contact being the emergency department or outpatient department/clinic. COVID-19 would be considered as a possible etiology in patients presenting with acute respiratory illness.

The U.S. Government provides an Interim Clinical Guidance for Management of Patients with Confirmed Coronavirus Disease (COVID-19) by Centers for Disease Control and Prevention as an agency of the U.S. Department of Health and Human Services (Centers for Disease Control and Prevention, 2020a, 2020b).

According to the Interim Guidance, for those with mild illness, hospitalization is not required unless there is concern about rapid deterioration or inability to return promptly to hospital. “Mild illness” is defined as, “Patients with uncomplicated upper respiratory tract viral infection that may have non-specific symptoms such as fever, fatigue, cough (with or without sputum production), anorexia, malaise, muscle pain, sore throat, dyspnea, nasal congestion, or headache. Patients may also present with diarrhea, abdominal pain, nausea and vomiting. Many are afebrile or have low-grade fever. The elderly and immunocompromised may present with atypical symptoms. Symptoms due to physiologic adaptations of pregnancy or adverse pregnancy events, e.g., dyspnea, fever, GI symptoms or fatigue, may overlap with COVID-19 symptoms.” (Government of Canada, 2020). “In admitted patients with suspected COVID-19, the diagnosis should be first attempted with an upper respiratory specimen, preferably a nasopharyngeal swab. However, a nasal and/or throat swab can be collected as an alternative if nasopharyngeal swabs are not available.” (Government of Canada, 2020). All patients outside hospital care are instructed to follow public health protocols for self-isolation and return to hospital if symptoms worsen. Patients are provided with information on how to treat mild symptoms. If they develop signs and symptoms of worsening disease, like difficulty breathing, pain or pressure in the chest, confusion, drowsiness, or weakness, they should seek follow-up care.

The definition of “mild” used to triage the patients is a label set at the discretion of the healthcare providers at the point of contact. Due to the burden of managing a pandemic, many patients will be asked to return home and follow the SOC protocol above. This study will utilize the WURSS-

©2020 Vielight Inc.
All rights reserved

44, a validated self-report measure of cold symptoms and severity (described further in Section 4). Question 1 of the WURSS-44 is a self-report Likert scale of ‘how sick do you feel today’, which ranges from a score of 0 (asymptomatic) to 7 (severe). Non-hospitalized patients reporting a WURSS-44 (Question 1) score of ≥ 4 will be recruited to allow for capture of any significant changes between the Treatment group against the SOC-only group.

#### 2.3 Vielight RX Plus Device Description

##### 2.3.1 Design Principles

The Vielight RX Plus (Figure 1) is a home-use photobiomodulation (PBM) device designed to deliver near-infrared (810 nm) and red (633 nm) light (or photons) to the sternal bone marrow, thymus gland, lungs and nasal tissues. It is hypothesized that PBM action can inhibit nasal coronavirus replication, elevate immune cell function, control inflammation, and thus act to accelerate recovery and reduce viral infection morbidity. PBM involves the application of red (600-700 nm) or near-infrared (760-1200 nm) light to modulate body functions. Light within this electromagnetic spectrum is absorbed by cellular chromophores, which triggers signaling pathways that then activate transcription factors and change protein expression, all of which can positively affect physiological functions in the body (de Freitas et al, 2016).

PBM has the potential to assist impaired cellular-mediated immunity: one of the hallmarks of fighting COVID-19 infection. PBM can be applied to the whole body or discrete areas through light emitting diodes (LEDs). The Vielight RX Plus is designed to deliver red and near-infrared light via LEDs placed directly inside the nose or on a discrete area. In this case, the RX Plus provides one LED module to be placed on the manubrium of the sternum (which covers the thymus gland) and inside one nostril (Figure 1). The photons of the near infrared (NIR) 810 nm light from the sternal LED is also expected to benefit the lungs through PBM effects. Another applicator is applied inside one of the nostrils to inhibit coronavirus replication. This is discussed further in Section 2.4 below.

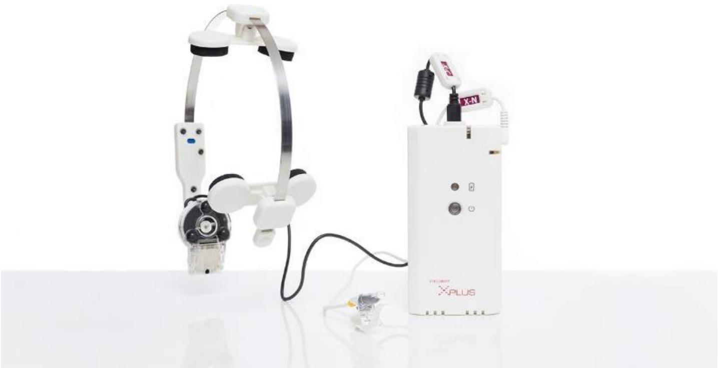

Figure 1: The Vielight RX Plus with sternal LED and intranasal LED

##### 2.3.2 Regulatory Status

The Vielight RX Plus has been available in its current commercialized form as the “Vielight X-Plus”: a low risk general wellness device with no medical claims attached. It has the status of a

©2020 Vielight Inc.
All rights reserved

Class II medical device registered with the United States Food and Drug Administration (FDA) but has not yet been available commercially under this status.

Sales of the commercial Vielight X-Plus has been suspended voluntarily and will re-emerge as the renamed “Vielight RX Plus” if cleared by Health Canada as a medical device to treat COVID-19.

##### 2.4 Mechanisms of Action of the Vielight RX Plus

A white paper, “Can the Vielight X-Plus be a Therapeutic Intervention for COVID-19 Infection?”, co-authored with Prof. Michael Hamblin of Massachusetts General Hospital and Harvard Medical School provides full explanation of the mechanisms underlying therapy with the Vielight RX Plus. The discussion below condenses the relevant information. The paper accompanies this protocol in Appendix 2.

###### 2.4.1 Fundamental cellular mechanisms

The fundamental mechanisms of PBM are based on the absorption of photons by the mitochondria to modulate cellular functions. The RX Plus delivers light of specific wavelengths (633 nm and 810 nm), power and duration to the body to achieve this. The biological process involves numerous interacting mechanisms that modulate bodily functions (de Freitas et al, 2020). One result of PBM is the regulation of the immune system (Novoselova et al, 2006). PBM can benefit a weakened immune system, and in healthy cases, could be prophylactic. The central mechanisms of PBM revolve around stimulating the mitochondrial respiratory chain where a transient release of non-cytotoxic levels of reactive oxygen species (ROS) leads to positive modulation of the immune response. PBM has a regulatory role via crosstalk with nuclear factor kappa-light-chain-enhancer of activated B cells (NF-κB) for the management of various conditions, including immune-related ones (Rajendran et al, 2019). In an immuno-compromised system, PBM leads to an increased production of white blood cells, while managing inflammation. In short, the appropriate dose of PBM directed to the mitochondria could positively modulate the immune system to address viral infections.

###### 2.4.2 Targeting the Thymus Gland, Sternal Bone Marrow and Lungs

Figure 2 below shows how the LED of the RX Plus is positioned to treat the thymus gland, sternal bone marrow and lungs. The LED is placed over the manubrium of the sternum.

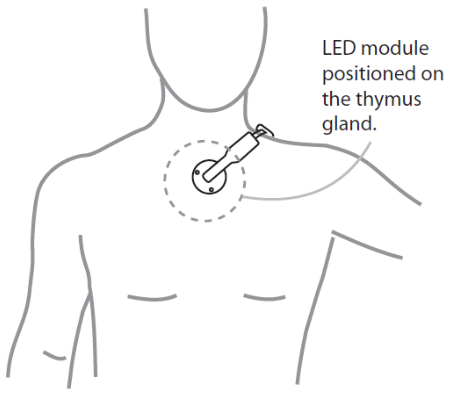

Figure 2: LED module of the RX Plus positioned over the sternum and thymus gland

©2020 Vielight Inc.
All rights reserved

The RX Plus has a NIR LED module placed on the manubrium of the sternum, and from that position, concurrently irradiates the thymus gland, sternal bone marrow and the lungs.

PBM activates stem cell mobilization that leads to tissue repair and production of white blood cells to support the immune system (Khorsandi et al, 2020). In the case of T-lymphocytes, these are produced in the bone marrow and mature in the thymus gland. Controlled doses of PBM to the thymus gland are expected to improve its function and hence improve maturation of T-cells. Although, the thymus gland loses density as we age, it is rational to believe that it remains active throughout adult life (Bertho et al, 1997; Kendall 1984). Hence the opportunity for thymic stimulation is never lost. It was demonstrated in an animal study that PBM irradiation with short treatments at a low power level showed elevated IL-2 cytokines, which are products of activated CD4+ T-lymphocytes (Novoselova et al, 2006).

###### 2.4.3 Targeting the Respiratory Tract

Viral respiratory infections, such as COVID-19 affects both the upper respiratory tract (nasal cavity, pharynx, larynx) and the lower respiratory tract (trachea, primary bronchi, lungs). COVID-19 has the ability to migrate to the lung and access the host type II alveolar cells, the most abundant type of alveoli where gas exchange takes place. As the alveolar disease progresses, respiratory failure ensues and death could follow (Xu et al, 2020).

The Vielight RX Plus could interrupt the passage of coronavirus from the upper to lower respiratory tracts in two ways, by application of:

1. an intranasal LED device in the nasal cavity
2. an LED module positioned on the manubrium of the sternum

Both components of the RX Plus may also contribute an additional beneficial systemic effects that are characteristic of PBM.

###### 2.4.4 Intranasal PBM on Respiratory Infections

Discussion on the effect of intranasal PBM here involves two broad pathways: 1) The systemic effect through irradiating the free-floating mitochondria and 2) direct inhibition of the virus through the production of nitric oxide (NO). The intranasal applicator of the RX Plus is used as shown in Figure 3, below. Its LED delivers red light at 633 nm.

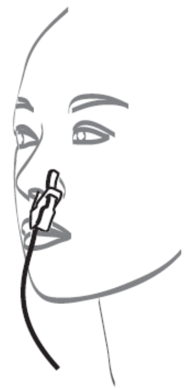

Figure 3: Intranasal applicator in use

©2020 Vielight Inc.
All rights reserved

###### 2.4.5 The Systemic Effect of Irradiating the Free-Floating Mitochondria

The nasal cavity has been chosen to position an LED for access to the dense local blood capillary networks which are shielded by a very thin light-permeable membrane in the nasal mucosa. This makes it relatively easy for light from the nasal LED to reach the blood circulatory system. PBM has a body-wide systemic effect that could be mediated by free-floating mitochondria (Al Amir Dache et al, 2020).

###### 2.4.6 Directly Inhibiting Coronavirus through Nitric Oxide Release

Nitric oxide (NO) is typically produced in the PBM process involving the mitochondria (de Freitas et al, 2016). The available mitochondria in the nasal passages produces NO, which is possibly part of the defense system against bacterial and viral infections (Lundberg et al, 1995). In viral infections, NO effects are complex and can be protective or deleterious (Uehara et al, 2015). The presence of NO has been found to inhibit the replication of coronavirus (Akerström et al, 2005). A PBM nasal applicator, as used by the RX Plus, would therefore present an opportunity to stop the infection before its migration to the lower respiratory tract where it causes the main damage.

###### 2.4.7 Direct Photobiomodulation of the Lower Respiratory Tract

When positioned on the manubrium of the sternum, the NIR light from the LED module of the RX-Plus may reach the lungs (Figure 2). The earlier discussion on NO having the beneficial effect of inhibiting the replication cycle of coronavirus (Akerström et al, 2005) is also relevant here.

In addition, it has also been found that direct PBM therapy of viral-infected cells has an inhibitory effect when compared to uninfected cells (Lugongolo et al, 2017). This could have been facilitated by the receptivity of the virus to NIR wavelength (Liu et al, 2003). Several studies showed that PBM can lead to increased count of various components of the white blood cell complex, that includes macrophage and T-lymphocyte concentration (Zheng et, 1992, Lim et al, 2009). This points to the benefits of irradiating the lungs directly.

The lungs can become acutely weakened as COVID-19 infection progresses. A parallel can be drawn in chronic obstructive pulmonary disease (COPD). In a double-blind study involving COPD patients, PBM therapy of the respiratory muscles was effective in improving acute functional capacity in the patients (de Souza et al, 2019).

In summary, positioning the RX Plus module on the manubrium allows the benefit of activating the functions of the bone marrow, thymus gland and protecting the lungs. The LED module of the RX Plus for this area emits light of 810 nm. This wavelength was chosen because it has widely been recognized for its penetration depth into mammalian tissues (Byrnes et al, 2005).

###### 2.4.8 Calming the Cytokine Storm

In studies, PBM has been shown to promote inflammation through NF-$_{\kappa}$B proteins in normal cells. However, in the presence of excessive inflammatory markers, PBM has been shown to behave differently. It has been shown to be anti-inflammatory (Hamblin, 2017). The anti-inflammatory characteristic of PBM is expected to calm a potential cytokine storm.

##### 2.5 Clinical Data and Summary of Related Evidence to Date

Presently, there is no direct clinical data supporting the Vielight RX Plus as an effective treatment for COVID-19, hence the reason for this pilot study. However, as presented in the preceding

©2020 Vielight Inc.
All rights reserved

paragraphs, the technology is hypothesized to enhance the immune system. Benefits include inhibiting coronavirus replication; elevating the immune system; and managing inflammation. From the Vielight device side, there is evidence of tissues (mainly neuronal) responding to its PBM devices. These features are summarized below:

###### 2.5.1 Inhibition of coronavirus replication in the nasal passages

The action of nitric oxide (NO) released in PBM (de Freitas et al, 2016) directly inhibits the replication of coronavirus in the nasal passages (Lundberg et al, 1995).

###### 2.5.2 Development of T-lymphocytes

PBM mobilizes stem cells in the sternal bone marrow (Oron, 2019), which migrate to the thymus gland for maturation. The NIR light is also expected to support the activity of the thymus gland (Odinokov, 2019).

###### 2.5.3 Attacking pathogens in the respiratory tracks, including the lungs

Lymphoid stem cells are available in the respiratory tracts for activation into mature lymphocytes by PBM to attack pathogens (Al Musawi et al, 2017).

###### 2.5.4 Production of nitric oxide in the respiratory tract tissues

In the same way for the nasal passage as described above, NO is produced when near infrared photons reach the tissues of the respiratory tracts, including the lungs to inhibit coronavirus replication (de Freitas et al, 2016; Akerström et al, 2005).

###### 2.5.5 Support recovery of damaged cells

PBM has established mechanisms in the production of growth factors to promote cellular regeneration (de Freitas et al, 2016)

###### 2.5.6 Management of inflammation

PBM has been shown to control inflammation generated through the activities of the cytokines mitigating the risk of a cytokine storm (Hamblin, 2017)

###### 2.5.7 Vielight devices have produced evidence of tissue responses

Vielight devices have demonstrated in peer-reviewed literature that tissues such as those in the brain respond to NIR. This is measured with electroencephalography (EEG) (Zomorrodi et al, 2019) and functional magnetic resonance imaging (fMRI) (Chao, 2019). Numerous other studies have shown that different tissues have positive responses to PBM (de Freitas et al, 2017), which can potentially be related to a device such as the Vielight RX Plus.

###### 2.5.8 Track record of efficacy in other clinical studies

PBM with Vielight devices were used in other clinical studies, producing positive clinical outcomes. These include positive clinical outcomes for dementia (Saltmarche et al, 2017; Chao, 2019) and traumatic brain injury (Naeser et al, 2016). These devices share the same platform as the RX Plus proposed in this COVID-19 study.

#### 3.0 Study Objective and Rationale

##### 3.1 Study Objective

The objective of this study is to obtain data on the efficacy of the Vielight RX Plus in decreasing time to recovery of symptoms in subjects with COVID-19.

The study will be conducted among COVID-19 positive subjects at home in self-isolation via an EDC. There will be no physical contact between the subjects and the Investigators or other study staff. This study aims to demonstrate that the Vielight RX Plus is a useful adjunct to SOC.

##### 3.2 Study Design Rationale

The study design is aimed at obtaining data to assess the safety and efficacy of the Vielight RX Plus while recognizing the safety needs and practical feasibility under the present highly restrictive environment. There are several objective biomarker assays that may be conducted to assess illness progression, such as measures of immune response (e.g. interleukin levels and lymphocyte count), and viral load. However, due to self-isolation procedures, and limited supply of medical equipment required to collect and assess these biomarkers, a self-report measure of symptoms and severity is most practical and reduces the risk of spreading the infection. The WURSS-44, although designed for symptoms of the common cold, contains questions for variables that cover respiratory symptoms that are characteristic of COVID-19. By utilizing a self-report measure, the study will not require the participant to break self-isolation and social-distancing measures and allows for daily report of illness progression.

Currently there are no vaccines, monoclonal antibodies, or drugs available to treat the virus, SARS-CoV-2 (Centers for Disease and Control, 2020c). Due to the novelty of the virus, the existence of any viable treatment remains an unanswered question, although drug discovery efforts have dominated the search for a cure. Medical devices as therapy have remained largely unexplored. Although several factors support the proposal that the Vielight RX Plus is a device that could be viable for addressing COVID-19, safety and efficacy have not yet been established, and warrants further investigation.

#### 4.0 Overview of Study Design

The study will be managed by an independent clinical research organization (CRO), supporting a Qualified Investigator (QI) in Canada and a Principal Investigator (PI) in the United States. Vielight Inc will supply the RX Plus devices free of charge and will sponsor the study. The study is not dependent on the success of achieving external financial support.

A diagram of study flow is shown in Figure 4.

©2020 Vielight Inc.
All rights reserved

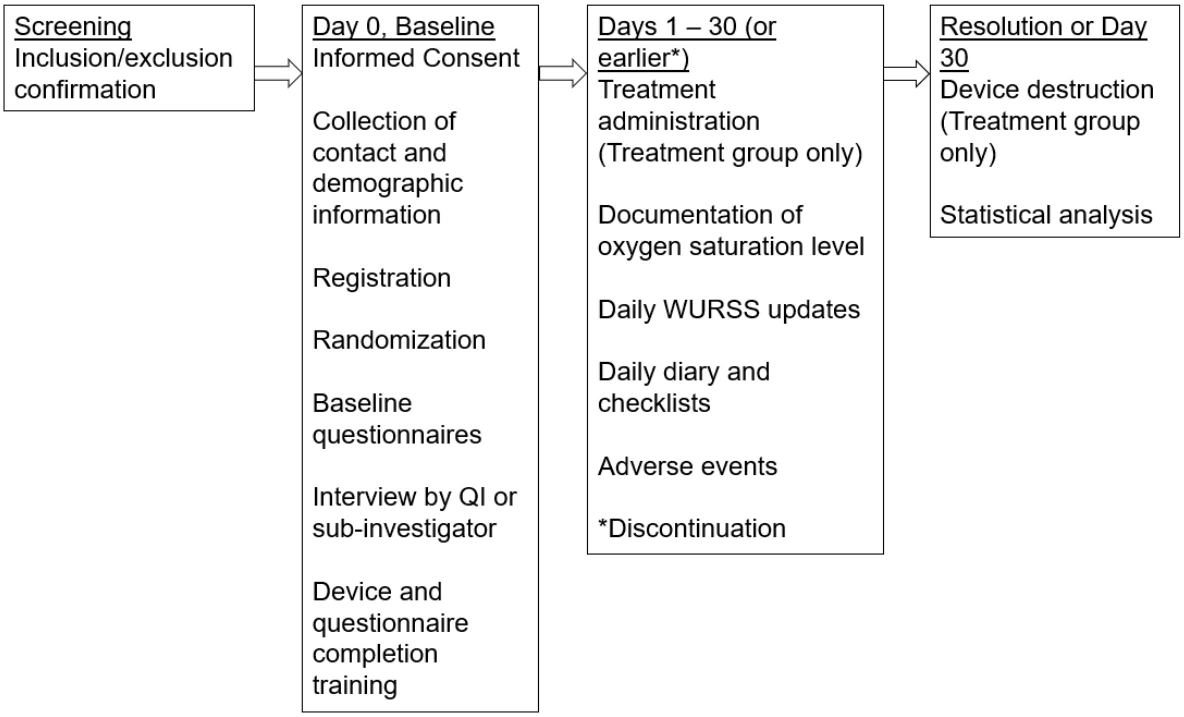

Figure 4. Study Flow Chart

#### 5.0 Study Procedures

##### 5.1 Screening

The study information and pre-screening form will be available on the study website. Potential participants will be prompted to complete a screening form if they are interested in participating. The Uniform Resource Locator (URL) for the study website will be included on all recruitment methods. The pre-screening form will capture the following information:

1. Participant age range (18-30, 31-45, 46-55, 56-65)
2. Range in days since symptom onset (i.e. 0-5 days, 6-10 days, or greater than 10 days),
3. WURSS-44 Question 1 (0 – 7)
4. Setting (at home or hospitalized)
5. Need for supplemental oxygen in past 24 hours (Yes or No)
6. Other medical condition such as Chronic Obstructive Pulmonary Disease (COPD), Hepatitis C Virus (HCV), Hepatitis B Virus (HBV) or Human Immunodeficiency Virus (Yes or No)
7. Daily access to an electronic device (laptop, desktop, tablet, cell phone) and internet (Yes or No).
8. COVID-19 positive test (Yes or No)

The potential subject will be asked to upload a copy of their positive COVID-19 infection confirmation report and a valid ID to the secure file sharing platform if they select ‘Yes’ to having a positive COVID-19 test.

©2020 Vielight Inc.
All rights reserved

The EDC will generate for each individual a unique Screen ID number and generate a Screening log. The Screening log will include the full name, address, telephone number, and any other pertinent subject information. This log is the primary way in which the subject identification for consented subjects will be correlated to the actual identity of the subject. Study staff will review the Screening log and information submitted by the potential participant. The Screening log will have access restricted to only study staff, ensuring confidentiality is strictly maintained.

##### 5.2 Electronic Informed Consent Form and Emergency Contact

The potential participant will be prompted to complete the informed consent form (ICF) module which will include review of study procedures, overview of the medical device, their rights and responsibilities and all aspects of the ICF, followed by a comprehension test. Potential participants will be prompted to contact study staff if they have any questions regarding the device and/or study procedures. Potential participants will also be asked if they have any additional questions through a prompt once they have completed the study overview module. Study staff will contact the potential participant to discuss any questions via a telemedicine platform call and will document that all participant questions have been answered.

The electronic informed consent form will be completed via the EDC by all participants prior to completing study procedures. The ICF used under this study protocol will be consistent with applicable elements of ISO 14155: 2011 Good Clinical Practice Guidelines and the Tri-Council Policy Statement: Ethical Conduct for Research Involving Humans, December 2010, Canada's Medical Device Regulations Part 3(81)(k) and 21 CFR Part 50 Protection of Human Subjects.

Subjects who have signed the ICF will be given a unique subject number that shall be used to identify the subject on all study related documents. The EDC will automatically generate a Study ID and generate an enrollment log.

Participants will be requested to provide emergency contact information and consent to the - Investigators and study staff. This information will be used to reach out to the emergency contact in the event that the subject stops responding to questionnaires and study staff are unable to contact the participant to conduct a safety assessment. Study information, training materials and ICF will reiterate that the participant is responsible for their health, that Vielight Inc., and study staff are not monitoring their health, and that they are instructed to follow local public health guidance for seeking medical assistance and care.

##### 5.3 Registration and Randomization

This is a prospective, randomized study, with no blinding. Subjects who provide informed consent will be registered on the enrolment log via the EDC with a unique Study ID.

Study staff will complete a randomization form using OxMAR minimization software. Participants will be randomly allocated to the Treatment group or SOC group (1:1) based on the minimization algorithm which aims to reduce the differences in the two groups based on baseline factors (i.e. duration of days since symptom onset (< or = 5 or 6-10 days) and WURSS Q1 score (4-5 or 6-7). Once baseline information has been entered into the OxMAR minimization software study staff will be notified which arm the participant has been allocated to via email.

Participants will receive a notification from the EDC regarding group assignment and details of the study requirements. Each participant will also be provided with a link to a secure private portal unique to them. All questionnaires, diaries and checklists will be available in the portal for baseline and daily access.

©2020 Vielight Inc.
All rights reserved

##### 5.4 Shipment of Devices

Vielight RX Plus device and/or fingertip pulse oximeter will be delivered to participant within 24 hours of randomization. Participants will be provided with a shipment notification, tracking information and will confirm receipt of the device via the EDC. Once confirmation of the device has been indicated on the EDC, the participant will be prompted to complete a scheduling form to select a date and time for completing the device training and baseline call with study staff.

###### 5.4.1 Device Training

Training will be provided for the device(s) via an EDC module. Training for the Vielight RX Plus will be provided in instructional online/digital material on how to use the device and deal with potential issues. Instructional materials will also be included with each device shipment. There will also be instructions for how to destroy the device at the end of the study. Instructions and training on how to use the pulse oximeter will also be provided via an EDC module. Training and baseline information will be reviewed by study staff with participants via a telemedicine platform call prior to initiating intervention procedures.

##### 5.5 Baseline (Day 0)

Prior to conducting any study procedures or interventions, the participant will complete study training module which includes videos and instructions on completion of study forms, use of the device(s), and how to contact study staff.

All participants will complete the following baseline forms:

- WURSS-44
- Race/ ethnicity questionnaire
- Fitzpatrick scale (It is a scale to capture skin tone in addition to the ethnicity and race self-report)
- Medical history and concomitant medications questionnaire
- Training review

Following completion of baseline forms the participant will complete an interview with study staff to review baseline information, review device training, and address any additional questions regarding the study procedures. Study staff will confirm with participants that they are to seek medical attention according to their local public health guidance, and confirm that Vielight and study staff are not taking the place of the participant's health care provider.

- Participant will then be asked to record baseline oxygen saturation using the pulse oximeter.

Participants will be scheduled for an introductory call with the study investigators a telemedicine platform, who will make the final assessment of subject's study eligibility.

##### 5.6 Days 1 to 30

The participant will be instructed to complete the electronic WURSS-44 form, daily diary and Oxygen saturation at around the same time of day (exact time and date recorded) to have consistency of readings via the EDC portal.

The primary endpoint for the subject is completed at 30 days, or when there is discontinuation because of emergency hospitalization, death or a voluntary withdrawal by the subject. However, a positive resolution does not discontinue the study procedures as data continues to be collected

©2020 Vielight Inc.
All rights reserved

until Day 30. A positive resolution is when the subject scores 0 on the WURSS-44 form (Question 1) for 3 consecutive days, which indicates that they no longer have symptoms.

In the event that there is missing data that does not meet the threshold for subject withdrawal from the study per definition of non-compliance, the missing interval will use the score from the last available observation (see Section 12.7 for non-compliance and protocol deviations).

During the study, research staff will be available during normal business hours to provide any requested assistance. An after-hour contact number (24 hour) will be provided to the participant for any safety events.

In the event that a participant does not complete a daily questionnaire, they will be sent a notification from the EDC. Study staff will also contact them via phone call within 24 hours to confirm that participant is aware of the study tasks, and whether any safety event(s) have occurred. If the participant is not reachable, study staff will attempt to contact the listed emergency contact to confirm the participants' safety. In the event that a safety event has occurred, study staff will speak with the participant or emergency contact to obtain any details regarding this event.

##### 5.7 Final, Day 30

Day 30 is the final day for self-reporting and data collection by subject. Day 30 will include an additional CRF collecting participant disposition as detailed in Section 7.2. The subject in treatment group will be given instruction about device destruction. Participants will submit proof of device destruction via the EDC.

##### 5.8 Subject Withdrawal

Participants may be withdrawn from the study for any of the following reasons:

1. Subject's choice to withdraw their consent to participate. Any participant has the right to withdraw from the study at any time and for any reasons without prejudice to future medical care by the physician or the institution. This will be considered a screening failure if they withdraw from the study prior to being randomized. Participants who withdraw consent after randomization will have any available data evaluated, up to the time of withdrawal.
2. Investigator's choice to withdraw the subject from the study. This will be considered a screening failure if the Investigator withdraws the subject prior to randomization.
3. Study termination for either administrative or safety reasons.

For participants who withdraw their consent for study participation, the Investigator will complete the "End of Study CRF" and no additional data will be collected after the date of their discontinuation.

Participants will be instructed to notify the study team in the event that they become pregnant during the course of the study. Participants will be withdrawn if they become pregnant and recorded using the 'End of Study CRF'.

##### 5.9 Treatment Protocol

The Vielight RX Plus is self-administered for 20 minutes twice a day, separated at least by 6 hours for the first 5 days. Subsequently, it is used once daily for 20 minutes. The banded NIR LED module is positioned over the manubrium of the sternum and intranasal applicator inside the left or right nostril for the rest of the study duration of 30 days.

©2020 Vielight Inc.
All rights reserved

As a note, the reduction of treatment time from twice daily to once daily after 5 days takes into account the need to minimize the risk of plateauing efficacy or inducing a negative effect. This is a characteristic of PBM in the literature (Huang et al, 2011).

### 6.0 Eligibility Criteria and Recruitment

#### 6.1 Recruitment Strategy

Potential subjects are expected to be recruited utilizing a variety of recruitment methods, including advertisements (e.g. brochures and posters at COVID-19 screening centres and medical clinics), a study website, social media and physician referrals.

#### 6.2 Inclusion Criteria

Only subjects with the following characteristics are eligible for study entry:

- Confirmation of SARS-CoV-2 with PCR test or alternative
- WURSS-44 Question 1 score of 4-7
- Between 18 to 65 years of age

#### 6.3 Exclusion Criteria

Subjects with the following characteristics are not eligible for study entry:

- Need for hospitalization at the time of diagnosis
- Current need for supplemental oxygen or positive pressure support and/or has required supplemental oxygen or positive pressure support for >or= 24 hours
- > 10 days from symptom onset
- Pregnant
- Inability to complete daily study forms online (in English)
- Diagnosis of Chronic Obstructive Pulmonary Disease (COPD)
- Positive for Hepatitis C Virus (HCV), Hepatitis B Virus (HBV) or Human Immunodeficiency Virus

### 7.0 Study Endpoints

#### 7.1 Primary Endpoint

Time to overall recovery in days as measured by item 1 of the WURSS-44 “How sick do you feel today”. Recovery is defined as a rating of 0 (not sick) that is confirmed over 3 consecutive days.

#### 7.2 Secondary Endpoints

©2020 Vielight Inc.
All rights reserved

A. Time to elimination of symptoms in days on the following items in the WURSS-44, where elimination of symptoms is defined as a rating of 0 (do not have this symptom or not at all) that is confirmed over 3 consecutive days:
    - Item 2 “Cough”
    - Item 12 “Body aches”
    - Item 18 “Feeling tired”
    - Item 37 “Breathe easily”
    - Item 16 “Feeling feverish”
B. Mean number of days with a rating of 0, 1, 2 or 3 for the following items:
    - Item 1 “How sick do you feel today”
    - Item 2 “Cough”
    - Item 12 “Body aches”
    - Item 18 “Feeling tired”
    - Item 37 “Breathe easily”
    - Item 16 “Feeling feverish”
C. Mean number of days with mild overall respiratory symptoms as measured by a total WURSS-44 score of <=129
D. Time to reduction in symptoms in days as measured by item 1 of the WURSS-44 “How sick do you feel today”. Reduction in symptoms is defined as a rating of 3 or less that is confirmed over X days.
E. Time to elimination of symptoms in days on the following items in the WURSS-44, where elimination of symptoms is defined as a rating of 0 (do not have this symptom or not at all) that is confirmed over 3 consecutive days:
    - Item 16 “Feeling feverish”
    - Item 37 “Breathe easily”

F. Average number of days spent with mild respiratory symptoms for the 7 questions relating to respiratory symptoms defined below (WURSS-44 respiratory score < or = 21) in treatment group compared to SOC.
WURSS-44 rating on past 24hrs for
    - ‘cough’,
    - ‘coughing stuff up’,
    - ‘cough interfering with sleep’,
    - ‘chest congestion’,
    - ‘chest tightness’,
    - ‘heaviness in chest’
    - ‘breathe easily’
G. Oxygen Saturation by Pulse Oximetry [Time Frame: 30 Days]
H. Percentage of subjects reporting ‘yes’ for each disposition category at day 30: 1) Death; 2) Hospitalized, on invasive mechanical ventilation or extracorporeal membrane oxygenation (ECMO); 3) Hospitalized, on non-invasive ventilation or high flow oxygen devices; 4) Hospitalized, requiring supplemental oxygen; 5) Hospitalized, not requiring supplemental oxygen - requiring ongoing medical care (COVID-19 related or otherwise); 6) Hospitalized, not requiring supplemental oxygen - no longer requires ongoing medical care; 7) Not hospitalized, limitation on activities and/or requiring home oxygen; 8) Not hospitalized, no limitations on activities.
I. Mortality rate

©2020 Vielight Inc.
All rights reserved

#### 8.0 Statistical Methods

##### 8.1 Sample Size Justification

The study will enroll 280 patients in a 1:1 randomization and the primary analysis will be performed when all patients have completed treatment, regardless of the number of recovery events observed.

If, as expected, nearly all patients should recover within the 30-day treatment period, then a hazard ratio of 1.40 can be detected with approximately 80% power with a two-sided significance level of 5%.

If a substantial proportion of patients (i.e. approximately 15%) do not record a recovery within the 30-day treatment period, then a hazard ratio of 1.44 can be detected with approximately 80% power with a two-sided significance level of 5%.

##### 8.2 Analysis Populations

Endpoints will be assessed in two study populations, the per protocol (PP) and intention-to-treat (ITT) population. All participants who complete all study procedures will per protocol will be included in the PP analysis. Participants who have protocol deviations (as described in Section 12.7). ITT population will include all enrolled participants. For participants with missing data, the missing timepoint will be populated using the last observation.

##### 8.3 Primary Endpoint: Recovery from 4-7 to 0 in WURSS-44 mean scores: Log-rank Test

Time-to-event is defined as the interval from the date of first use of the device to the start date of the improvement. Three consecutive days of reporting 0 on Question 1 of the WURSS-44 is considered durable improvement. The start date of the improvement will be taken to be the first day where the criteria is met. Subjects who have not met the definition of improvement will be censored as of their last available assessment. Subjects without a baseline assessment or post-baseline assessment will be censored at the date of randomization.

Time-to-improvement will be estimated using Kaplan-Meier methods. The proportion of subjects with an improvement will be reported by treatment group. The overall group comparison will be done using a log-rank test stratified by time since onset of symptoms (< or = 5 days vs. 6-10 days). The hazard ratio and its associated 95% confidence interval will be calculated based on a stratified Cox proportional hazards model.

##### 8.4 Secondary Endpoints

###### 8.4.1 Outcomes based on mean number of days

Outcomes based on mean number of days will be computed in two ways. The first method will utilize all available assessments. The second method will assume a 30-day follow-up and will consider any missing assessment as not having met the criteria.

The mean number of days meeting a certain criterion will be analyze using an ANCOVA model with a fixed effect for treatment group assignment and a covariate of time since onset of symptoms (< or = 5 days vs. 6-10 days).

©2020 Vielight Inc.
All rights reserved

##### 8.4.2 Outcomes based on time-to-event

The following endpoint assesses a time-to-event, and will be assessed as described for the primary endpoint, utilizing a Log-rank test:

> Time to elimination of symptoms in days on the following items in the WURSS-44, where elimination of symptoms is defined as a rating of 0 (do not have this symptom or not at all) that is confirmed over 3 days:
>
> - Item 2 “Cough”
> - Item 12 “Body aches”
> - Item 18 “Feeling tired”
> - Item 37 “Breathe easily”
> - Item 16 “Feeling feverish”

##### 8.4.3 Descriptive Statistics

Secondary endpoints assessing between group proportions will be assessed via chi-square test.

Participant demographic, and baseline measurements will be presented in contingency tables as mean, standard deviation (SD). Baseline values, post-Treatment values, and change from baseline to post-Treatment will be compared between the groups using independent samples t-test, with statistical significance at $p < 0.05$, and 95% confidence intervals reported.

#### 8.5 Assessment of Device Safety: Incidence of device attributable adverse events and usability

The commercial version of the RX Plus has been commercially available over 2 years, with no report of any major side effects, other than the occasional report of a warm feeling from the 810 nm LED targeted for placement over the manubrium. It has the same controller and driver platforms as the Vielight Neuro RX Gamma which has been tested for safety and approved for a clinical trial by Health Canada. Unrelated to safety, the other factor that may arise is a malfunction of the device, in which case the CRO will liaise with the Sponsor who will take corrective action. Notwithstanding the anticipated absence of major issues, a mechanism is in place for the subjects to report an adverse event to the CRO, who will document it and report to the Sponsor for further corrective action.

A device-related event is determined by the “relatedness score” for Adverse Events. A relatedness score of “Possible” or “Probably” or “Definite” indicate the extent that the Adverse Event is related to the use of the device. Adverse Events receiving a relatedness score of “Unrelated” or “Unlikely” are considered unrelated to the device use and are not included. All safety endpoints will be adjudicated by one of the study administrators.

A statistical comparison of the incidence rates will be conducted. Additionally, an evaluation of any events due to the general nature of the device (i.e. headset discomfort, nasal irritation) and not specific to the delivery of the NIR energy will be qualitatively evaluated. All emergent findings from participant reported oxygen saturation will be documented and reviewed by the QI to complete an AE form and update the AE log if required.

©2020 Vielight Inc.
All rights reserved

##### 8.6 Interim Analysis

A single interim analysis will be conducted after 73 participants completed their study. To preserve the overall type I error rate at the nominal level of 0.05, the stopping boundary for efficacy will be set at $p<0.0051$ (O'Brien and Fleming, 1979). The stopping boundary for futility will be set at the conditional power of 50% (Lan et al, 1982).

#### 9.0 CRO and Data Collection Methods

This study is conducted remotely, via an EDC with oversight from a Contract Research Organization (CRO). The PI, QI, sub-investigators and study staff will conduct study procedures and responsibilities remotely. All study procedures will be completed by participants online while they are at home in self-isolation.

##### 9.1 Study Duration

Recruitment rate of subjects is to be determined based on availability. The duration of the study is 30 days for each subject, from first use of the device. The total expected study duration from first subject enrolled to final subject follow up will be approximately 6 months.

#### 10.0 Ethical Considerations

##### 10.1 Consent Process

Consent shall be obtained in accordance with Canada's Medical Device Regulations section 81(k)(ii) and the Tri-Council Policy Statement – Ethical Conduct for Research Involving Humans (TCPS2) and World Medical Association Declaration of Helsinki – Ethical Principles for Medical Research Involving Human Participants, and 21 CFR Part 50 Protection of Human Subjects.. Potential subjects must be informed as to the purpose of the study and the potential risks and benefits known or that can be reasonably predicted or expected as described in the written consent form. The subject shall have sufficient opportunity to consider participation in the study; consent forms shall be written in English. A subject cannot be led to believe that they are waiving their rights as a subject or the liability of the sponsor or investigator. Subjects are then invited to sign and date the consent form, indicating their consent for enrollment. (Investigators may not date the consent form on the subject's behalf.)

##### 10.2 Institutional Review Board (IRB) and Research Ethics Board (REB) Approvals

For this study, IRB and REB approvals are optional under an interim order by the Ministry of Health, "Expedited Review of Health Product Submissions and Applications to address COVID-19". The Sponsor will apply for an approval by an IRB.

##### 10.3 Data Protection

A Subject Identification Log shall link the data collected during the study to source documents such as medical records. The Subject ID Log shall be kept in a secure and confidential manner with access restricted to study staff.

All information collected during the course of this study will be kept confidential, except the Sponsor's representatives and regulatory authorities will have access to this information.

©2020 Vielight Inc.
All rights reserved

The Sponsor's representative and/or regulatory authorities will have access to relevant medical information (i.e. COVID-19 positive test report) for purposes for source data verification, including medical information for monitoring and reporting of safety events.

No information which could identify a subject will be used in reports or publications. Subjects will remain de-identified for data analysis.

The source documents will be made available for future review with authorization from the Sponsor, subject to privacy laws.

The EDC is privacy compliant in Canada (PIPEDA and PHIPA), the U.S. (HIPAA).

##### 10.4 Study Suspension or Early Termination

The study can be discontinued at the discretion of the study Sponsor for reasons including, but not limited to, the following:

- Obtaining new scientific knowledge that shows that the study is no longer valid or necessary
- Insufficient recruitment of subjects
- Unanticipated adverse device effect (UADE) presenting an unreasonable risk to subjects
- Persistent non-compliance with the protocol
- Persistent non-compliance with regulatory requirements

As defined in 21 CFR 813.3, an unanticipated adverse device effect means any serious adverse effect on health or safety or any life-threatening problem or death caused by, or associated with, a device, if that effect, problem, or death was not previously identified in nature, severity, or degree of incidence in the investigational plan or application (including a supplementary plan or application), or any other unanticipated serious problem associated with a device that relates to the rights, safety, or welfare of subjects.

As per 21 CFR 812.150, if Sponsor determines that an unanticipated adverse device effect presents an unreasonable risk to subjects, all investigations or parts of investigations presenting that risk shall be terminated, as soon as possible. Termination shall occur not later than 5 working days after Sponsor makes this determination and not later than 15 working days after the sponsor first received notice of the effect. The study shall not be resumed without FDA and IRB approval.

If the study is discontinued or suspended, the Sponsor shall promptly inform the PI, QI, CRO and participants of the termination or suspension and the reason(s) for this. The IRB will also be informed promptly and provided with the reason(s) for the termination or suspension by the Sponsor or by the clinical investigator/investigation center(s). Regulatory authorities and the personal physicians of the subjects may also need to be informed if deemed necessary.

#### 11.0 Investigational Devices

Device Inventory Log records, the reference/part number, lot number, date received, date used, and PIN of the study subject it was used for, will be maintained by the CRO. Devices will be shipped to the subjects through courier services or driver contractors. Once the study is over (i.e. after 30 days), the subjects will be asked to destroy the device by cutting the wires and sending photos for proof.

A. Labeling: All products will include labeling on the inner pouch and shelf carton. All products will include Instructions for Use included in the shelf carton.

©2020 Vielight Inc.
All rights reserved

B. Storage: All products at the CRO will be stored at room temperature in a secure location.

C. Return: If the Device is associated with a device related adverse event, malfunction or failure, the Device should be returned to Vielight Inc. for appropriate action, or destroyed as instructed by the Sponsor.

#### 12.0 Safety and Adverse Events

##### 12.1 Definition of Adverse Events

An adverse event is any unfavourable and unintended sign, including an abnormal laboratory finding, symptom, or disease temporarily associated with the use of a medical treatment that may or may not be considered related to the medical treatment or procedure.

Adverse events (AEs) may occur after enrolment but prior to the start of treatment or during the treatment period. Adverse events occurring prior to the start of treatment, while not directly associated with the use of the Vielight RX Plus, will be documented in the subject's medical record but will not count as a study device-related event. Participants will be asked to report any adverse events in a daily basis through their daily diary. If the subjects are experiencing a device related adverse event, they will be instructed to contact research staff. Participants will be instructed to follow their local public health guidance on seeking medical attention. It is the participants' responsibility to monitor their own health and to seek medical attention per their local public health guidance and instructions from their local health care provider.

Investigators will record characteristics of each adverse event on an Adverse Event CRF. Each adverse event will be judged by the Investigator as to its relationship and level of relatedness to the investigational device. Relatedness will be scored consistent with CTCAE v4.0 guidelines. CTCAE v4.0 is the National Cancer Institute (USA) Common Terminology Criteria for Adverse Effects version 4.0 (published August 9, 2006) is a descriptive terminology which can be used for Adverse Event (AE) reporting.

- Unrelated – the AE is clearly not related to the investigational agent(s),
- Unlikely – the AE is doubtfully related to the investigational agent(s),
- Possible – the AE may be related to the investigational agent(s),
- Probable – the AE is likely related to the investigational agent(s),
- Definite – the AE is clearly related to the investigational agent(s).

In addition, the Investigator will identify the date of onset, severity and duration. Severity is determined from the grades presented in CTCAE v4.0. The CTCAE v4.0 displays Grades 1 through 5 with unique clinical descriptions of severity for each AE based on this general guideline:

- Grade 1 – Mild AE
- Grade 2 – Moderate AE
- Grade 3 – Severe AE
- Grade 4 – Life-threatening or disabling AE
- Grade 5 – Death related to AE

©2020 Vielight Inc.
All rights reserved

An AE is defined as being serious (SAE) if it results in death or meets the definition of serious injury, as defined in Title 21 of the Code of Federal Regulations, Section 803.3 (21 CFR 803.3), which means an injury or illness that:

i. Is life-threatening,
ii. Results in permanent impairment of a body function or permanent damage to a body structure; or
iii. Necessitates medical or surgical intervention to preclude permanent impairment of a body function or permanent damage to a body structure. Permanent means irreversible impairment or damage to a body structure or function, excluding trivial impairment or damage.

As defined in 21 CFR 803.3, caused or contributed means that the adverse device effect was or may have been attributed to the study device, or may have been a factor in in the event, including events occurring as a result of:

- Failure,
- Malfunction,
- Improper or inadequate design,
- Manufacture,
- Labeling, or
- User error.

All adverse events will be monitored until they are adequately resolved or explained.

##### 12.2 Definition of Device Incident

An *incident* according to Canada's Medical Device Regulations Section 59 (1) and Good Clinical Practice (http://www.fda.gov/RegulatoryInformation/Guidances/ucm122046.htm) is defined as an event that:

(a) is related to a failure of the device or a deterioration in its effectiveness, or any inadequacy in its labeling or in its directions for use; and

(b) has led to the death or a serious deterioration in the state of health of a subject, user or other person, or could do so were it to recur.

##### 12.3 Reporting Adverse Events and Device Incidents

All Serious Adverse Events (SAEs, see Definitions) must be reported to the study Sponsor immediately, not to exceed 24 hours after the investigator first learns of the event. The completed adverse event investigation form shall be faxed or emailed to the Sponsor within 24 hours together with a cover letter describing the event and detailing the medical history, concomitant medication, and an assessment of compliance with therapy.

All adverse events, serious or non-serious adverse events will be reported in the daily diary

All SAEs need to be followed until the event is resolved (with or without sequelae). The PI will decide if more follow up information is needed in case the event is not resolved at study completion. In case of death, all possible information that is available, including the possible relationship to the device, should be provided.

©2020 Vielight Inc.
All rights reserved

The investigator must submit to the Sponsor any unanticipated adverse device incident (see Definitions) within 24 hours after the investigator first learns of the effect. The investigator must also report the unanticipated adverse device effect to the IRB within its pre-specified timeline. As per 21 CFR 812.150, this incident reporting shall not be later than 10 working days after the investigator first learns of the effect. The Investigator will report all of the above to the reviewing IRB (as applicable) according to their reporting requirements. Reports must identify subjects using the study's unique identifier to protect subject's confidentiality.

The Sponsor will be responsible for submitting incident information to Health Canada and FDA within the specified timeframe. As per 21 CFR 812.46, Sponsor shall immediately conduct an evaluation of any unanticipated adverse device effect. Sponsor shall report the results of such evaluation to FDA and to the reviewing IRB and participating investigators within 10 working days after receiving notice of the effect. Thereafter the sponsor shall submit such additional reports concerning the effect as FDA requests.

##### 12.4 Adverse Event Follow-up

The investigator shall continue to monitor any unanticipated, device related adverse event until it is resolved or up to one month following the end of the study.

##### 12.5 Anticipated Adverse Events

There are no adverse events anticipated to be device attributable with use of the Vielight RX Plus. The device predecessor (Vielight X-Plus) has been currently sold as a general wellness device worldwide with no major adverse events having been reported to date. It satisfies the criteria of an unregulated low risk device defined by the FDA policy, "General Wellness: Policy for Low Risk Devices".

##### 12.6 Incidental Findings

Review of the participant's oxygen saturation and other study data may result in incidental findings. All incidental findings captured by the CRO will be reviewed by the QI/PI. The participant may be notified of any incidental findings if they significantly deviate from the normal range. The participant will be instructed to seek medical attention per their local public health guidance, and according to their health care provider. It is at the discretion of the participant to follow up with their health care provider regarding any incidental findings.

##### 12.7 Protocol Deviations

A protocol deviation is defined as any study action taken by the QI/PI to be in conflict with the Study Protocol. Investigators must make every effort to follow the protocol except where necessary to protect the life or physical well-being of a subject in an emergency. All deviations from the protocol will be reported on the appropriate Deviations Log.

Deviations must be reported to the Sponsor within a reasonable time frame. Subject specific deviations will be reported on the Deviations Log. Non-subject specific deviations, (e.g. unauthorized use of an investigational device outside the study, etc.), will be reported to the QI/PI. Investigators will also adhere to procedures for reporting study deviations to the IRB/REB if applicable.

Regulations require that Investigators maintain accurate, complete and current records, including documents showing the dates of and reasons for each deviation from the protocol.

©2020 Vielight Inc.
All rights reserved

For participants who are noncompliant with follow up visits, every effort will be made to collect the data requested by this protocol through the follow up period. All data collected up until the period the subject has withdrawn from the study will be used for this study. A participant will be defined as non-compliant if they do not perform at least one treatment per day in the first 5 days of treatment. Participants will also be defined as non-compliant if they miss 5 consecutive days of treatment prior to reaching the criteria for recovery.

#### 13.0 Study Management

##### 13.1 Sponsor Overall Responsibility

As the study sponsor, Vielight Inc. has the overall responsibility for the conduct of the study according to Canada's Medical Device Regulations Part 3, E6 Good Clinical Practice Consolidated Guidance, ICH, 1997), ISO 14155: Part 1 and 2, the Declaration of Helsinki, Medical Device Directive, Annex X, conditions imposed by the reviewing IRB/REB, Health Canada, FDA 21 CFR Part 812, 21 CFR Part 50, 21 CFR Part 56 and all applicable local regulatory requirements. For this study, Vielight Inc. will have certain direct responsibilities and will delegate other responsibilities to appropriate consultants and contract research organizations (CROs). Together, Vielight Inc., consultants and CROs will ensure that the study is conducted according to all applicable regulations. All personnel to participate in the conduct of this clinical trial will be qualified by training, education and/or experience to perform his or her respective tasks.

NOTE: A complete list of participating investigators will be maintained and will be available upon request.

##### 13.2 Investigator Responsibilities

The Investigator shall be responsible for the day-to-day conduct of the investigation as well as for ensuring that the investigation is conducted according to all signed agreements, applicable elements of ISO 14155, the Clinical Investigational Plan, applicable Health Canada regulations, applicable FDA regulations, the principles that have their origin in the Declaration of Helsinki, and any conditions of approval imposed by Health Canada. The investigator is also responsible for protecting the rights, safety and welfare of subject's under the investigator's care and for obtaining informed consent in accordance with Canada's Medical Device Regulations Part 3(81)(k) and FDA's regulations 21 CFR Part 50 Protection of Human Subjects and 21 CFR Part 812 Investigational Device Exemptions. Each Investigator must sign the Investigator Agreement and Financial Disclosure prior to subject enrolment. No investigator will be added to the investigation until a signed Investigator Agreement is provided.

Responsibilities of the Investigator include, but are not limited to:

1. Submit proposed amendments to the protocol and informed consent to the IRB and await approval, unless the change reduces the risk to subjects.
2. Ensuring that all personnel assisting with the clinical trial are adequately informed and understand their trial-related duties and functions
3. Obtain informed consents of subjects.
4. Permit monitor to inspect facilities and records.
5. Maintain medical histories of subjects.
6. Enroll subjects, execute the study, transcribe data from source documents to case report forms, and conduct study in accordance with protocol.

©2020 Vielight Inc.
All rights reserved

7. Submit annual progress reports, final reports, and Adverse Event reports to the IRB and to Sponsor.

8. Notify the local or national authority and the manufacturer (Vielight Inc.) within 72 hours of any incident that meets the criteria as defined in Subsection 59(1) of the Medical Devices Regulations. [Subsection Sub-section 59(1) defines two types of device related incidents. The first is related to a failure or deterioration in the effectiveness of the device or any inadequacy in its labelling or in the directions for use accompanying it. The second involves an incident that has led to the death or a serious deterioration in the health of a subject, user or other person or, where it is reasonable to believe that such an incident were it to recur, could lead to the death or a serious deterioration of the state of health of a subject, user or other person.

9. Disclose to Vielight Inc. sufficient accurate financial information to allow them to submit completed and accurate certification and disclosure statements under US 21 CFR54. The investigator shall promptly update this information if any relevant changes occur during the course of the investigation and for 1 year following completion of the study

   The Investigator will allow direct access to source data/documents for trial related monitoring, audit, IRB review and regulatory inspection. Also, the investigator will allow auditing of their clinical investigational procedure(s).

##### 13.3 Required Documents from the Investigators

At a minimum, the following documents will be provided by the Investigators to the study sponsor:

- Signed Investigator Agreement
- Current Curriculum Vitae
- Completed and accurate certification and disclosure statements under US 21 CFR54
- Any other relevant documents requested by the study sponsor, IRB or other regulatory authority

The study procedures will not be initiated until all of the above listed documents have been provided to the study sponsor.

##### 13.4 Investigator Records

The Investigator is responsible for maintaining medical and study records for every subject participating in the clinical study (including information maintained electronically such as digital imaging). The Investigator will also maintain original source documents from which study-related data are derived, which include, but are not limited to:

- all correspondence including required reports with another investigator, an IRB, the sponsor, Health Canada, FDA or any other competent authority.
- records of each subject's case history and exposure to the device which must include,
- signed and dated consent forms
- condition of each subject upon entering the study
- relevant previous medical history
- observations of relevant adverse device effects (anticipated or unanticipated)
- adverse events reporting and follow-up of the adverse events
- medical records (physician and nurse progress notes, hospital charts, etc.)
- results of all laboratory tests

©2020 Vielight Inc.
All rights reserved

- notations on abnormal lab results
- case report forms
- subjects' condition upon completion of or withdrawal from the study
- any other supporting data
- the protocol and documentation (date and reason) for each deviation from the protocol.
- any other records that Health Canada, FDA or other regulatory body requires to be maintained by regulation or by specific requirement for a category of investigation or a particular investigation.
- The Investigator must ensure that all subject records are stored for at least 2 years after the end of the clinical study or the records are no longer required to support a regulatory approval, whichever date is later, per 21 CFR 812.140(d).. To avoid error, the CRO should contact Vielight Inc. prior to the destruction of study records to ensure that they no longer need to be retained. In addition, Vielight Inc. should be contacted if the Investigator plans to leave the study so that arrangements can be made for the handling or transfer of study records. Records must be retained in designated administrative files at each Investigational Site and/or retained by the study sponsor
    - Trial protocol and all amendments
    - Ethics Committee Approval Letter(s) and approved informed consent(s) (including any revisions)
    - Device Instructions for Use
    - CVs of all investigators
    - Site Subject Log
    - Monitoring reports
    - Site Authorized Personnel Signature List
    - Reports (includes Adverse Event reports and final reports from investigator and sponsor)

##### 13.5 Specific Sponsor Responsibilities

Vielight Inc. is the Sponsor of this study. Vielight Inc.'s responsibilities include but are not limited to:

1. Selecting investigators that are qualified by training and education (qualifications will be documented by curriculum vitae)
2. Providing investigators with the information necessary to conduct the investigation properly
3. Providing appropriate training to each the CRO and all study personnel (monitors), as necessary
4. Documenting training where appropriate
5. Selecting monitors qualified by training and experience to monitor the investigational study in accordance with FDA and Health Canada regulations or train monitors if necessary.
6. Ensuring that the IRB approval is obtained
7. Ensuring that any reviewing IRB or regulatory authority are informed of significant new information
8. Providing the devices to qualified investigators
9. Report and investigate unanticipated, device related Adverse Events

©2020 Vielight Inc.
All rights reserved

10. Obtaining signed Investigator Agreement for each investigator prior to their participation in the study
11. Obtaining sufficient and accurate financial disclosure information
12. Reporting per local regulations
13. Retain records for at least 2 years following completion of this study.

##### 13.6 Sponsor Records

The Sponsor shall maintain the following accurate, complete and current records relating to the investigation:

- All correspondence including required reports
- Signed investigator's agreements including financial disclosure information
- Records concerning adverse device effects and complaints

##### 13.7 Study Monitoring Plan

The study will be monitored to ensure that applicable regulations are followed. Written procedures will be established in a monitor plan to ensure the quality of the study and to ensure that each person involved in the monitoring process carries out the required duties. The sponsors shall designate or assign monitors to this clinical study.

Source-documents will be reviewed to verify CRFs and database information for completion and accuracy. CRF findings of non-compliance or required modifications shall be reviewed with the Investigator(s). The monitor will report to the sponsor any non-compliance with the signed Investigator's Agreement, conditions imposed by the IRB or regulatory authorities. The sponsor shall then either secure compliance, or terminate the investigator's participation in the investigation (Section 85(2)(b) of the Medical Device Regulations).

An initial review of study progress and compliance will be conducted after the first 5 participants have completed baseline procedures. At this time the monitor will review study progress and compliance with the protocol, including but not limited to, review of rate of recruitment, number of screen failures, EDC queries and time to resolution, time between screening, training, and baseline procedures, record of participant queries and study documents.

Interim monitoring assessments will be conducted after each recruitment quartile (i.e. at 35 participants in each group, followed by 70/group, and 105/group), and once all participants have completed the study procedures for study-close out. Monitoring visits may occur at other timepoints at the Sponsor's discretion. At each monitoring visit the CRO will provide the Sponsor designated monitor with the database lock for that time-point, along with any other requested study material for review.

##### 13.8 Medical Monitoring

An independent Medical Monitor will review safety reports biweekly. The reports will contain the number and type of adverse events, and also all the serious adverse events classified as probably or definitely related to enrolment in the trial. The Medical monitor will review all SAEs within 48 hours of their report. The Medical Monitor will have the ability to request additional safety analyses and make recommendations about the safe conduct of the trial. Within 7 business days of the review date, the medical monitor will communicate any findings or recommendations directly with Principal Investigator**.** The Medical Monitor can recommend stopping the investigation due to safety reasons.

©2020 Vielight Inc.
All rights reserved

Participants will be instructed to seek medical attention per local public health guidance and according to their healthcare practitioner. Incidental findings from oxygen saturation monitoring and WURSS self-reports will be documented and reviewed by the investigator, and participants may be notified if these values are out of normal range, however, it is up to the participant's discretion whether to seek medical care and follow-up with their health care provider.

#### 13.9 Ethical Considerations

The rights, safety and well-being of clinical investigation subjects shall be protected consistent with the ethical principles laid down in the Declaration of Helsinki. This shall be understood, observed and applied at every step in this clinical investigation.

It is expected that all parties will share in the responsibility for ethical conduct in accordance with their respective roles in the investigation. The Sponsor and the Investigator(s) shall avoid improper influence or inducement of the subject, monitor, the clinical investigator(s) or other parties participating in or contributing to the clinical investigation.

##### 13.10 Protection of Subject Confidentiality

At all times throughout the clinical investigation, confidentiality will be observed by all parties involved. All data shall be secured against unauthorized access. Privacy and confidentiality of information about each subject shall be preserved in study reports and in any publication. Each subject participating in this study will be assigned a unique identifier. All CRFs will be tracked, evaluated, and stored using only this unique identifier.

The EDC will be overseen by the CRO, and will maintain a confidential subject list identifying all enrolled subjects (Enrollment Log). This list will contain the assigned subject's unique identifier and name. The EDC is Health Insurance Portability and Accountability Act (HIPAA) compliant.

Monitors and auditors will have access to the subject list and other personally identifying information of subjects to ensure that data reported in the CRF corresponds to the person who signed the ICF and the information contained in the original source documents. Such personal identifying information may include, but is not limited to date of birth, sex, race and COVID-19 test report.

Any source documents copied for monitoring purposes by the Sponsor will be identified by using the assigned subject's unique identifier in an effort to protect subject confidentiality.

##### 13.11 Quality Assurance and Supervision by Authorities and Privacy

All documents and data shall be produced and maintained in such a way to assure control of documents and data to protect the subject's privacy as far as reasonably practicable. The EDC technology platform is privacy compliant in Canada (PIPEDA and PHIPA), the U.S. (HIPAA) and will undergo regular monitoring as detailed in 13.7. The Sponsor and representatives of the regulatory authorities are permitted to inspect the study documents (e.g., study protocol, CRFs, and original study-relevant medical records/files) as needed. All attempts will be made to preserve subject confidentiality.

##### 13.12 Final Report

A final report will be completed, even if the study is prematurely terminated. At the conclusion of the trial, an abstract reporting the results will be prepared and will be presented at a major meeting(s). A manuscript will also be prepared for publication in a reputable scientific journal.

©2020 Vielight Inc.
All rights reserved

##### 13.13 Information Confidentiality

All information not previously published concerning the test device and research, including patent applications, manufacturing processes, basic scientific data, etc., is considered confidential and should remain the sole property of Vielight Inc. All information and data generated in association with this study will be held in strict confidence and remain the sole property of Vielight Inc. The Investigator agrees to use this information for the sole purpose of completing this study and for no other purpose without written consent from Vielight Inc.

##### 13.14 Trial Registration

The study will be registered in a publicly accessible trial database (clinicaltrials.gov) prior to study initiation.

#### 14.0 Study Close-out

##### 14.1 Timeline of Close-out

Study close-out will be conducted by the Sponsor's monitor within 30 days of receipt of the last case report form.

##### 14.2 Record Storage and Retention

The Sponsor shall retain copies of all study data and documentation for a minimum of 2 years after the last approval of a marketing application in an ICH region and until there are no pending or contemplated marketing applications in an ICH region or at least 2 years have elapsed since the formal discontinuation of clinical development.

##### 14.3 Discontinuation of Study

Vielight Inc. reserves the right to discontinue any study for business or ethical reasons at any time, such as but not limited to, a decision to discontinue further clinical investigations with the test article, improper conduct of the study by the investigator, inability to obtain the number of subjects required by the protocol, etc. Reimbursement for reasonable expenses will be made if such action is necessary.

#### 15.0 Definitions and Acronyms

**Adverse Event (AE)** - any untoward medical occurrence in a subject (ISO 14155). Note: This definition does not imply that there is a relationship between the adverse event and the device under investigation

An adverse event is any unfavourable and unintended sign (including an abnormal laboratory finding, symptom, or disease temporarily associated with the use of a medical treatment or procedure that may or may not be considered related to the medical treatment or procedure (CTCAE v 4.0).

**Serious Adverse Event (SAE)** - an adverse event that (ISO 14155):

- led to a death,
- led to a serious deterioration in the health of the subject,
- resulted in a life-threatening illness or injury,
- resulted in a permanent impairment of a body structure or a body function,

©2020 Vielight Inc.
All rights reserved

- required hospitalization or prolongation of existing hospitalization,
- resulted in medical or surgical intervention to prevent permanent impairment to body structure or function,
- led to fetal distress, fetal death, a congenital abnormality, or birth defect.

**Adverse Device Effect (ADE)** - any untoward and unintended response to a medical device, including that resulting from user error (ISO 14155)

**Serious Adverse Device Effect (SADE)** - an adverse device effect that has resulted in any of the consequences characteristic of a serious adverse event or that might have led to any of these consequences if suitable action had not been taken or intervention had not been made or if circumstances had been less opportune (ISO 14155).

**Unanticipated Adverse Device Effect (UADE)** - any serious adverse effect on health or safety or any life-threatening problem or death caused by, or associated with, a device, if that effect, problem, or death was not previously identified in nature, severity, or degree of incidence in the investigational plan or application (including a supplementary plan or application), or any other unanticipated serious problem associated with a device that relates to the rights, safety, or welfare of subjects (ISO 14155).

**Applicable Regulatory Requirement(s)-** Any law(s) and regulation(s) addressing the conduct of clinical trials of investigational products of the jurisdiction where trial is conducted (E6, GCP Guidance)

**Case Report Form (CRF)** - A printed document designed to record all of the protocol-required information to be reported to the sponsor on each subject (E6, GCP Guidance)

**Contract Research Organization (CRO)** - A person or an organization (commercial, academic or other) contracted by the sponsor to perform one or more of a sponsor's trial-related duties and functions (E6, GCP Guidance).

**COVID-19** - Coronavirus Disease 2019

**Documentation** - All records, in any form (including, but not limited to, written electronic, magnetic, and optical records; and scans. X-rays, and electrocardiograms) that describe or record the methods, conduct, and/or results of a trial, the factors affecting a trial and the actions taken (E6, GCP Guidance).

**Electronic Data Capture (EDC)** - is a computerized system designed for the collection of clinical data in electronic format for use mainly in human clinical trials.

**Electroencephalography (EEG)** - is an electrophysiological monitoring method to record electrical activity of the brain.

**Fitzpatrick scale -** is a numerical classification schema for human skin color

**Functional Magnetic Resonance Imaging (fMRI)** – A technique that measures brain activity by detecting changes associated with blood flow.

**Good Clinical Practice (GCP)**- A standard for the design, conduct, performance, monitoring, auditing, recording, analyses, and reporting of clinical trials that provides assurance that the data and reported results are credible and accurate, and that the rights, integrity, and confidentiality of trial subjects are protected (E6, GCP Guidance)

**HBV** - Hepatitis B Virus

**HCV** - Hepatitis C Virus

**HIV** - Human Immunodeficiency Virus

©2020 Vielight Inc.
All rights reserved

**Informed Consent** - The process by which the subject voluntarily confirms his or her willingness to participate in a particular trial, after having been informed of all aspects of the trial that are relevant to the subject's decision to participate. Informed consent is documented by means of a written, signed, and dated informed consent form.

**Informed Consent Form (ICF)** - a document that describes:

a. the risks and anticipated benefits to his or her health arising from participation in the clinical trial; and
b. All other aspects of the clinical trial that are necessary for that person to make the decision to participate in the clinical trial

**Institutional Review Board (IRB)**
IRB) - An independent body (a review board or a committee, institutional, regional, national or supranational), constituted of medical/scientific professionals and nonmedical/non-scientific members, whose responsibility is to ensure the protection of the rights, safety, and well-being of human subjects involved in a trial and to provide public assurance of that protection, by, among other things, reviewing and approving/providing favourable opinion on the trial protocol, the suitability of the investigator(s), facilities, and the methods and materials to be used in obtaining and documenting informed consent of the trial subjects (E6, GCP Guidance).

**Instructions for Use (IFU)** - Step by step directions for using the product.

**Investigator /Principal Investigator (PI)** - A person responsible for the conduct of the clinical trial. If a trial is conducted by a team of individuals, the investigator is the responsible leader of the team and may be called the principal investigator (E6, GCP Guidance).

**Monitoring** - Monitor when used as a verb means to oversee an investigation (ISO 14155). Monitoring is the act of overseeing the progress of a clinical trial, and of ensuring that it is conducted, recorded, and reported in accordance with the protocol, standard operating procedures, GCP and the applicable regulatory requirements (E6, GCP Guidance).

**NIR** - Near Infrared

**PBM** – Photobiomodulation

**Qualified Investigator -** A person who is a member in good standing of a professional association of persons entitled under the laws of a province to provide health care in the province and who is designated, by the ethics committee of the health care facility at which investigational testing is to be conducted, as the person to conduct the testing.

**SARS-CoV-2** - Severe Acute Respiratory Symptom Coronavirus 2.

**SOC** - Standard of care

**Source Data** - All information in original and identified records and certified copies of original records of clinical findings, observations, or other activities in a clinical investigation, necessary for the reconstruction and evaluation of the clinical investigation. Source data are contained in source documents (E6 GCP Guidance, ISO 14155).

**Source Documents** - Original documents, data and records (ISO 14155).

*Note: This may be, for example, hospital records, laboratory notes, pharmacy dispensing records, copies or transcriptions certified after verification as being accurate copies, photographic negatives, radiographs, and records kept at the pharmacy, at the laboratories, and at medico-technical departments involved in the clinical investigation.*

©2020 Vielight Inc.
All rights reserved

**Standard Operating Procedure (SOP)**

**Termination** -Termination means a discontinuation, by sponsor or by withdrawal of IRB or Health Canada approval, of an investigation before completion.

**Treatment** - Vielight RX Plus and Standard of Care (SOC).

**WURSS-44** - Wisconsin Upper Respiratory Syndrome Survey, 44 questions

©2020 Vielight Inc.
All rights reserved

©2020 Vielight Inc.
All rights reserved

#### Appendix 1 – Wisconsin Upper Respiratory Symptom Survey (WURSS-44)

##### Wisconsin Upper Respiratory Symptom Survey

***Daily Symptom Report***

| *Date:* | *Time:* | *ID*: |
|---|---|---|

Please fill in one circle for each of the following items.

| | **Not sick** | Very mildly | | Mildly | | Moderately | | Severely |
|---|---|---|---|---|---|---|---|---|
| | **0** | **1** | **2** | **3** | **4** | **5** | **6** | **7** |
| How sick do you feel **today**? | ○ | ○ | ○ | ○ | ○ | ○ | ○ | ○ |

Please rate the **average severity of your cold symptoms over the last 24 hours**.

| | **Do not have this symptom** | Very mild | | Mild | | Moderate | | Severe |
|---|---|---|---|---|---|---|---|---|
| | **0** | **1** | **2** | **3** | **4** | **5** | **6** | **7** |
| Cough | ○ | ○ | ○ | ○ | ○ | ○ | ○ | ○ |
| Coughing stuff up | ○ | ○ | ○ | ○ | ○ | ○ | ○ | ○ |
| Cough interfering with sleep | ○ | ○ | ○ | ○ | ○ | ○ | ○ | ○ |
| Sore throat | ○ | ○ | ○ | ○ | ○ | ○ | ○ | ○ |
| Scratchy throat | ○ | ○ | ○ | ○ | ○ | ○ | ○ | ○ |
| Hoarseness | ○ | ○ | ○ | ○ | ○ | ○ | ○ | ○ |
| Runny nose | ○ | ○ | ○ | ○ | ○ | ○ | ○ | ○ |
| Plugged nose | ○ | ○ | ○ | ○ | ○ | ○ | ○ | ○ |
| Sneezing | ○ | ○ | ○ | ○ | ○ | ○ | ○ | ○ |
| Headache | ○ | ○ | ○ | ○ | ○ | ○ | ○ | ○ |
| Body aches | ○ | ○ | ○ | ○ | ○ | ○ | ○ | ○ |
| Feeling "run down" | ○ | ○ | ○ | ○ | ○ | ○ | ○ | ○ |
| Sweats | ○ | ○ | ○ | ○ | ○ | ○ | ○ | ○ |
| Chills | ○ | ○ | ○ | ○ | ○ | ○ | ○ | ○ |
| Feeling feverish | ○ | ○ | ○ | ○ | ○ | ○ | ○ | ○ |
| Feeling dizzy | ○ | ○ | ○ | ○ | ○ | ○ | ○ | ○ |
| Feeling tired | ○ | ○ | ○ | ○ | ○ | ○ | ○ | ○ |
| Irritability | ○ | ○ | ○ | ○ | ○ | ○ | ○ | ○ |
| Sinus pain | ○ | ○ | ○ | ○ | ○ | ○ | ○ | ○ |
| Sinus pressure | ○ | ○ | ○ | ○ | ○ | ○ | ○ | ○ |
| Sinus drainage | ○ | ○ | ○ | ○ | ○ | ○ | ○ | ○ |

*More symptoms…* →

WURSS© 2001 Created by Bruce Barrett MD PhD et al, UW Department of Family Medicine, 777 S. Mills St. Madison, WI 53715

| | Do not have this symptom 0 | Very Mild 1 | 2 | Mild 3 | 4 | Moderate 5 | 6 | Severe 7 |
|---|---|---|---|---|---|---|---|---|
| Swollen glands | ○ | ○ | ○ | ○ | ○ | ○ | ○ | ○ |
| Plugged ears | ○ | ○ | ○ | ○ | ○ | ○ | ○ | ○ |
| Ear discomfort | ○ | ○ | ○ | ○ | ○ | ○ | ○ | ○ |
| Watery eyes | ○ | ○ | ○ | ○ | ○ | ○ | ○ | ○ |
| Eye discomfort | ○ | ○ | ○ | ○ | ○ | ○ | ○ | ○ |
| Head congestion | ○ | ○ | ○ | ○ | ○ | ○ | ○ | ○ |
| Chest congestion | ○ | ○ | ○ | ○ | ○ | ○ | ○ | ○ |
| Chest tightness | ○ | ○ | ○ | ○ | ○ | ○ | ○ | ○ |
| Heaviness in chest | ○ | ○ | ○ | ○ | ○ | ○ | ○ | ○ |
| Lack of energy | ○ | ○ | ○ | ○ | ○ | ○ | ○ | ○ |
| Loss of appetite | ○ | ○ | ○ | ○ | ○ | ○ | ○ | ○ |

**Over the last 24 hours,** how much has your cold interfered with your ability to:

| | Not at all 0 | Very mildly 1 | 2 | Mildly 3 | 4 | Moderately 5 | 6 | Severely 7 |
|---|---|---|---|---|---|---|---|---|
| Think clearly | ○ | ○ | ○ | ○ | ○ | ○ | ○ | ○ |
| Speak clearly | ○ | ○ | ○ | ○ | ○ | ○ | ○ | ○ |
| Sleep well | ○ | ○ | ○ | ○ | ○ | ○ | ○ | ○ |
| Breathe easily | ○ | ○ | ○ | ○ | ○ | ○ | ○ | ○ |
| Walk, climb stairs, exercise | ○ | ○ | ○ | ○ | ○ | ○ | ○ | ○ |
| Accomplish daily activities | ○ | ○ | ○ | ○ | ○ | ○ | ○ | ○ |
| Work outside the home | ○ | ○ | ○ | ○ | ○ | ○ | ○ | ○ |
| Work inside the home | ○ | ○ | ○ | ○ | ○ | ○ | ○ | ○ |
| Interact with others | ○ | ○ | ○ | ○ | ○ | ○ | ○ | ○ |
| Live your personal life | ○ | ○ | ○ | ○ | ○ | ○ | ○ | ○ |

**Compared to yesterday**, I feel that my cold is...

| Very much better | Somewhat better | A little better | The same | A little worse | Somewhat worse | Very much worse |
|---|---|---|---|---|---|---|
| ○ | ○ | ○ | ○ | ○ | ○ | ○ |

WURSS© 2001 Created by Bruce Barrett MD PhD et al, UW Department of Family Medicine, 777 S. Mills St. Madison, WI 53715

©2020 Vielight Inc.
All rights reserved

**Appendix 2 – White Paper**

### Can the Vielight X-Plus be a Therapeutic Intervention for COVID-19 Infection?

Lew Lim, *PhD* [1] and Michael R Hamblin, *PhD* [2]

1. Vielight Inc., Toronto, ON M4Y 2G6, Canada.
2. Wellman Center for Photomedicine, Massachusetts General Hospital, Boston MA 02114, USA,

   Department of Dermatology, Harvard Medical School, Boston, MA, 02115, USA, Harvard-MIT Division of Health Sciences and Technology, Cambridge, MA, 02139, USA.

April 7, 2020.

**Abstract**

**This paper proposes the use of Vielight X-Plus as a therapeutic intervention to address COVID-19. The device is based on the science of photobiomodulation (PBM), utilizing red and near infrared (NIR) light to modulate body functions. Fundamentally, PBM involves light of specific wavelengths acting on the respiratory chain of the mitochondria which then triggers higher-level, homeostasis restorative functions. In the process, PBM also improves the immune system to potentially attack viruses and help respiratory diseases. Light from ultraviolet to infrared may destabilize the envelope structures of coronaviruses, increasing the effectiveness of PBM. During PBM, nitric oxide is also released, which inhibits the replication of coronaviruses. PBM also reduces the risks of cytokine storm and sepsis associated with the uncontrolled activity of the immune system as the body fights the disease. The Vielight X-Plus was designed to take advantage of these mechanisms that could make it effective to address COVID-19. It has the added advantage of being portable, suitable for home use, and affordable, with the potential for prophylaxis. It now warrants a randomized controlled study to validate its benefits.**

**Introduction**

COVID-19 is a novel respiratory disease and hence its pathology is largely unknown at this time – recently published on February 28, 2020 are considered to be in early phase[1], although newer data are expected due to intensive global research. Currently there are no vaccines, monoclonal antibodies, or drugs available for the virus, SARS-CoV-2.[2] However, due to the novelty of the virus , the existence of any viable treatments remains an unanswered question although drug discovery efforts have dominated the search for a cure.[3] Medical devices have remained largely unexplored, except for mechanical ventilation and extracorporeal membrane oxygenation.

This paper is prepared in the context of the current COVID-19 pandemic and proposes a randomized controlled study to test the "Vielight X-Plus" as a treatment for COVID-19. It is currently commercially available as a general wellness device and can be mass-manufactured with a lead time of 2 months. The

©2020 Vielight Inc.
All rights reserved

technology has the potential for acute response with further potential as a prophylaxis. The X-Plus is based on the science of "photobiomodulation" (PBM), which involves the delivery of red and near-infrared light to the cells of the body from light emitting diodes (LEDs). The field of PBM has a substantial volume of research literature dating back to 1967, about 55 years ago. The device has passed independent electrical safety tests for public use.

**Fundamental cellular mechanisms**

The fundamental mechanisms of PBM are based on absorption of photons by the mitochondria to modulate cellular functions. The X-Plus delivers light of specific wavelength, power and duration to the body to achieve this. The process involves numerous interacting mechanisms that modulate bodily functions.[4] One result is the regulation of the immune system.[5] PBM elevates a weakened immune system, and in a healthy cases, could be prophylactic. The central mechanisms of PBM revolve around stimulating the mitochondrial respiratory chain where a transient release of non-cytotoxic levels of reactive oxygen species (ROS) leads to positive modulation of the immune response. PBM has a regulatory role via crosstalk with nuclear factor kappa-light-chain-enhancer of activated B cells (NF-κB) for the management of various conditions, including immune-related ones.[8] In an immuno-compromised system, the chain of activity leads to increased production of appropriate levels of white blood cells, while managing inflammation. In short, the appropriate dose of PBM directed to the mitochondria could positively modulate the immune system to address viral infections.

Figure 1: Vielight X-Plus

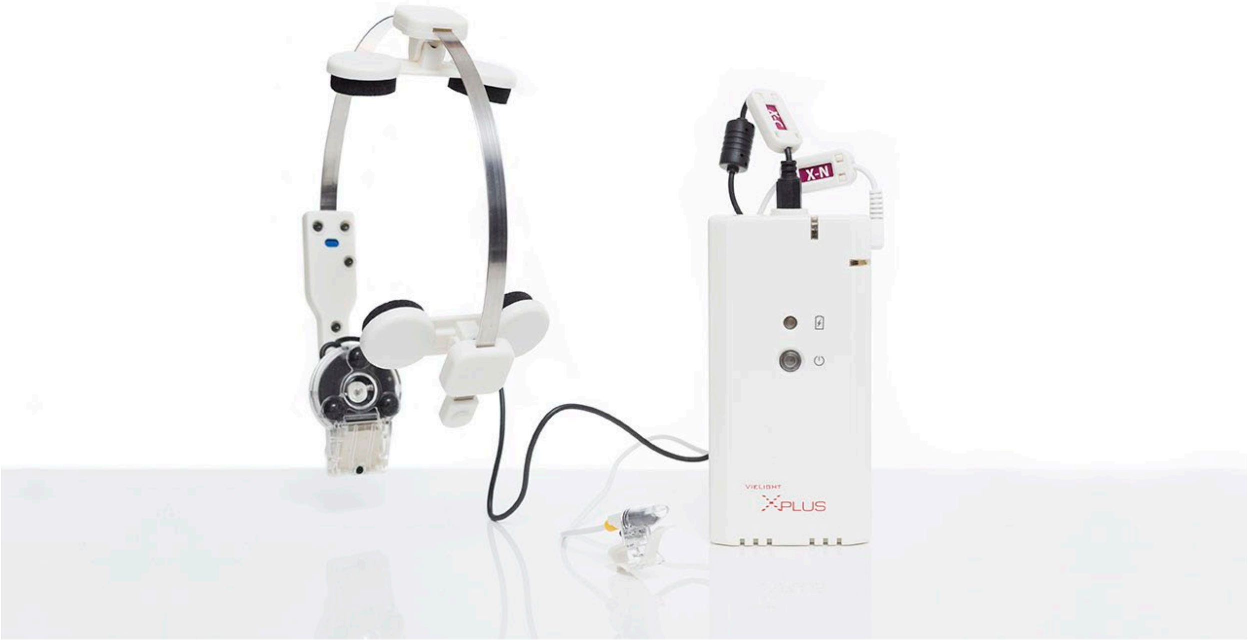

The coronavirus membrane protein in its envelope has a higher propensity to absorb light from ultraviolet to infrared. The process could destabilize the virus and makes it particularly amenable to further action of PBM.[5]

©2020 Vielight Inc.
All rights reserved

A molecule released during PBM is nitric oxide (NO). It is commonly identified with vasodilatation,[4] and improved blood circulation. However, in the context of COVID-19, its value to potentially to attack viruses is much more consequential.[31]

**The Key Success Factor is the Effect on the Immune System**

Many different viruses exist, and because they have a high propensity to mutate, this makes them perpetual moving targets for drug and vaccine treatments. Even if a treatment for COVID-19 is found today, we can assume that variants will emerge, which we hope will not be as transmissible or have as high a mortality rate. The coronavirus, SARS-CoV-2 differs substantially from related viruses that cause the common cold including four other types of coronaviruses (OC43, HKU1, NL63, and 229E) and the various influenza viruses.

The human body has repeatedly been shown to have the ability to adapt to the ever-morphing microbes and viruses. The ability to combat the moving targets rests heavily on the state of the immune system. It is therefore sensible to test ways to support and enhance the natural capacity to do this. The PBM modality can support the body's naturally ability to restore functional homeostasis, and in the process enhance the immune response against COVID-19 and future variants. Based on its specifications, the Vielight X-Plus may be a suitable choice for a PBM device.

*In vitro* studies irradiating blood serum samples with light have demonstrated significant proliferation of lymphocytes. A study using red light at 660 nm[6] increased reactive oxygen species (ROS), a free radical. This suggests that the ROS could have been produced by activated free-floating mitochondria[21], which then act as signals for the lymphocyte proliferation as a response. It may also suggest that there are naïve common lymphoid progenitor cells present in the blood [7] that could be activated by PBM for transformation into mature lymphocytes. There is growing evidence that stem cells are highly responsive to PBM resulting in changes to initiate cell proliferation and induce signaling cascades.[8]

In a seemingly confounding manner, yellow/orange (589 nm) light irradiation of blood serum showed greater lymphocyte proliferation than red (which is used in the X-Plus). In this *in vitro* experiment directly irradiating blood samples in tubes, there was a significant increase in CD45 lymphocytes and natural killer (NK) (CD16, CD56) cells. However, there was no significant change in CD3 T lymphocytes, T-suppressor (CD3, CD8) cells, T-helper (CD3, CD4) cells, and CD19 B lymphocytes.[9] The yellow/orange light produced more pronounced changes than blue or near infrared.

These *in vitro* experiments are not necessarily representative of *in vivo* body responses. In this regard, another study produced evidence-based arguments that the *in vitro* effects of light for exerting an

antiviral effect is not based on wavelength but on the ability to generate oxygen species such as ROS, which is produced in the PBM process.[26] We should therefore recognize tissue barriers to light and give more credence to: 1) action spectrum for PBM, and 2) wavelengths required for tissue penetration. The dose provided by the X-Plus through 633 nm (red) and 810 nm (NIR) light respects these requirements.

**Photobiomodulation with the Vielight X-Plus as a Potential Treatment for COVID-19**

Health Canada and the FDA have cleared a number of PBM devices for various medical indications. These include a variety of aesthetic applications, tissue healing and pain management. Vielight is

©2020 Vielight Inc.
All rights reserved

currently engaged in a regulatory-reviewed pilot and pivotal clinical trials to investigate the Vielight Neuro RX Gamma for Alzheimer's Disease. The cellular mechanisms at cellular levels allow for these benefits to apply across different organ systems, in higher-level restoration of tissue functions.

The literature suggests that PBM can be applied to many more medical conditions.[4] Those that involve improving the immune system and the management of excessive inflammation are relevant to treating COVID-19.[10]

Below, we discuss specific areas that relate to the immune system, and how the design of the Vielight X-Plus that could address the COVID-19 infection.

**Targeting the Thymus Gland, Sternal Bone Marrow and Lungs**

Figure 2 below shows how the LED of the X-Plus is positioned to treat the thymus, sternal bone marrow and lungs.

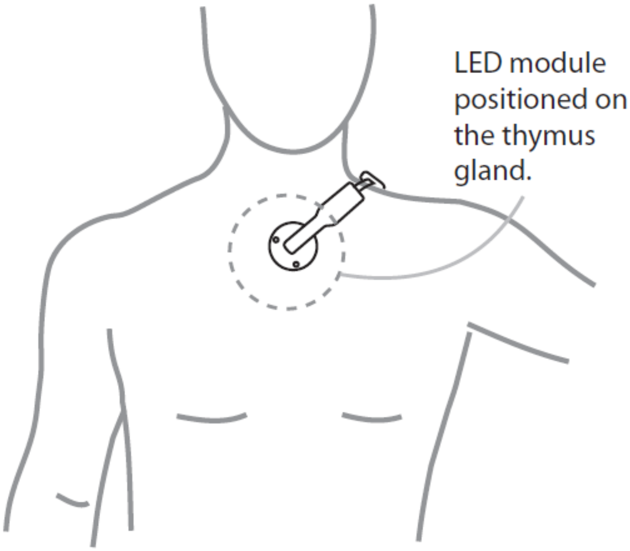

Figure 2: LED module of the X-Plus positioned over the thymus gland

The X-Plus has a NIR LED module placed on the sternum, and from that position, concurrently irradiates the thymus gland, sternal bone marrow and the lungs for PBM effects.

PBM activates stem cell mobilization that leads to tissue repair and production of white blood cells to support the immune system.[11] In the case of T-lymphocytes, these are produced in the bone marrow and mature in the thymus gland. Therefore, the application of a controlled dose of PBM to the thymus gland is expected to improve its function and hence improve maturation. Although, the thymus gland loses density as we age, it is rational to believe that it remains active throughout the adult life.[12 13] Hence the opportunity for thymic stimulation is never lost. PBM can activate the thymus gland and mitigate its reduction throughout its lifespan,[14] as well as activate the surrounding bone marrow to contribute to

©2020 Vielight Inc.
All rights reserved

mesenchymal stem cell genesis.[15] The overall effect helps to boost the immune system to resist a viral infection. An animal study grafting new tissue into aged animals, even showed that reviving the thymus may extend the lifespan – reflecting its importance beyond T-cell support.[16]

As evidence pertaining to T-lymphocytes, it was demonstrated in an animal study that PBM irradiation with short treatments at low-level showed elevated IL-2 cytokines, which are products of activated CD4+ T cells. In addition, NO was also produced. However, it should be noted that prolonged use reduced the positive effects, showing that there is a window for optimum results in PBM therapy.[17]

**Targeting the Respiratory Tract**

Viral respiratory infections, such as COVID-19 affects both the upper respiratory tract (nasal cavity, pharynx, larynx) and the lower respiratory tract (trachea, primary bronchi, lungs). The potency of COVID-19 is its ability to migrate to the lung and access the host type II alveolar cells, the most abundant type of alveoli where gas exchanges take place. Its entry is facilitated through the enzyme ACE2 recognized by its "corona" spikes.[18] As the alveolar diseases progresses, respiratory failure ensues, and death could follow.[19]

The Vielight X-Plus could have a role in intervening the passage of the coronavirus in two ways, by application of:

1. An intranasal LED device in the nasal cavity,
2. A LED module positioned on the on the thymus gland

Both components of the X-Plus may also contribute an additional beneficial systemic effect that is characteristic of PBM.

**Intranasal PBM on Respiratory Infections**

Discussion on the effect of intranasal PBM here involves two broad pathways: 1. The systemic effect through irradiating the free-floating mitochondria, and 2. Direct inhibition of the virus through the production of nitric oxide (NO). The intranasal applicator of the X-Plus is used as shown in Figure 3 below. Its LED delivers red light at 633 nm at a safe power density of 6.5 $mW/cm^2$.

Figure 3: Intranasal applicator in use

**The Systemic Effect of Irradiating the Free-floating Mitochondria**

The nasal cavity has been chosen to position a LED for access to the dense local blood capillary networks which are shielded by a very thin light-permeable membrane in the nasal mucosa. This makes it relatively easy for light from the X-Plus LED to reach the blood circulatory system and the requisite tissue. PBM has a body-wide systemic effect mediated by "circulating factors" described in early PBM literature[20], but recently identified as free-floating mitochondria.[21]

Therefore, the effect of therapeutic light can be delivered throughout the body by simply irradiating the blood capillaries in the vicinity of the nasal cavity. This effect is circulated and spread throughout the body through the major vessels where blood traverses 50 to 100 cycles an hour.

©2020 Vielight Inc.
All rights reserved

An early study using a red laser optic fiber to treat vasomotor rhinitis showed a significant increase of T-lymphocytes.[22] So the complex and cascading mechanisms that start from the activity of the free-floating mitochondria in blood in the nasal cavity are the likely factors behind boosting T-lymphocyte presence.

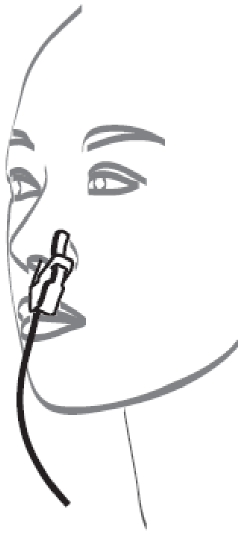

**Directly Inhibiting Coronavirus through Nitric Oxide Release**

Nitric oxide (NO) produced in the nasal passages is possibly part of the defense system against bacterial and viral infections.[23] In viral infections, NO effects are complex and can be protective or deleterious.[24] The presence of NO has been found to inhibit the replication of coronavirus.[31] A PBM applicator, as used by the X-Plus to release of NO this way, would therefore present an opportunity to stop the infection before its migration to the lower respiratory tract where it causes the main damage.

There are suggestions that ultraviolet (UV) C may help to eliminate viruses with direct irradiation[25] but prolonged exposure has carcinogenic risks[26]. Hence recommendation for this wavelength for use in the nasal cavity should be subject to further investigations for safety.

Blue light, which has been found to be anti-microbial is based on disintegrating cellular membrane in bacteria. There is no evidence that it is effective against virus unless an activator such as a singlet oxygen enhancer is introduced[27], in which case, toxicity risks emerges when applied internally.

In another application of light, there is research suggesting the use of visible (mainly blue or green) ultrashort pulsed laser (USPL) supported by compelling results for viral inactivation. It is based on resonance action of impulsive stimulated Raman scattering (ISRS) on the viral capsid.[28] This is not feasible for practical application for several reasons: 1) The equipment is experimental, large, relatively costly and requires training, 2) The necessity for ultra-short pulsing for viral inactivation has been legitimately challenged, [29] 3) It is only effective *in vitro*, 4) SARS-CoV-2 has a layer of envelope in addition to the capsid, which deemed USPL ineffective for this viral strain.

In summary, application of PBM in the nasal cavity with red LED of 633 nm wavelength inhibits coronavirus replication and activates the body's immune response system through whole-body signaling

©2020 Vielight Inc.
All rights reserved

cascades. The wavelength is also in an action spectrum for PBM effects.[30] This effect of the X-Plus is supported by scientific literature.[4]

**Direct Photobiomodulation of the Lower Respiratory Tract**

It is necessary to consider the possibility of direct irradiation of the lower respiratory tract, particularly the lungs where most of the COVID-19 consequential pathology takes place in a symptomatic patient. When positioned on the sternum, the NIR light from the LED module of the X-Plus may reach the lungs See Figure 3.

The earlier discussion on NO having the beneficial effect of inhibiting the replication cycle of coronavirus is also relevant here.[31]

In addition, it has also been found that direct PBM therapy of viral-infected cells has an inhibitory effect when compared to uninfected cells.[32] This could have been facilitated by the receptivity of the virus to NIR wavelength.[5] In another type of viral infection involving herpes simplex virus (HSV), irradiation with PBM also significantly lowered the incidence of local pathology recurrence.[33] Several studies showed that PBM can lead to increased count of various components of the white blood cell complex, that include macrophages and T-lymphocytes.[34 35] This points to the benefits of irradiating the lungs directly.

The lungs can become acutely weakened as the COVID-19 progresses. A parallel can be drawn in chronic obstructive pulmonary disease (COPD). In a double-blind study involving COPD patients, PBM therapy of the respiratory muscles was effective in improving acute functional capacity in the patients.[36]

In summary, positioning the X-Plus module on the sternum allows the benefit of activating the activities of the bone marrow, thymus gland and protecting the lungs. The LED module of the X-Plus for this area emits light of 810 nm. This wavelength was chosen because it has widely been recognized for its penetration depth into mammalian tissues.[37]

**Calming the Cytokine Storm**

Accumulating evidence suggests that a subgroup of patients with severe COVID-19 might have a cytokine storm syndrome.[38] A “cytokine storm” is an overproduction of immune cells and their secreted cytokines, which is often associated with a surge of activated immune cells into the lungs. The resulting lung inflammation and fluid buildup can lead to respiratory distress and can be contaminated by a secondary bacterial pneumonia – raising the mortality risk of patients.[39]

In studies, PBM has been shown to promote inflammation through NF-κB proteins in normal unstimulated cells. However, in the presence of excessive inflammatory markers, PBM has been shown to behave differently. It has been shown to be anti-inflammatory.[40] The anti-inflammatory characteristic of PBM is expected to calm a potential cytokine storm.

**Application in Rehabilitation**

Damage to lungs could lead to the risk of sepsis, caused by a weakened immune system continuing on overdrive. Statistics show that half of the survivors of lung sepsis suffer from other infections, kidney failure or cardiovascular problems, i.e. cardiovascular diseases, about three months after the incidence.

©2020 Vielight Inc.
All rights reserved

Furthermore, many sepsis patients suffer severe, long-term functional, cognitive or psychological consequences such as paralysis, depression or anxiety disorders. In 2017, the global burden of sepsis account for 49 million cases and 11 million deaths.[41] In an animal study, the findings suggested that LED application is an inexpensive and non-invasive treatment for lung sepsis that could be effective.[42]

**Application as Prophylaxis**

As discussed, PBM has been shown to modulate the body's own adaptive immune response, both locally and systemically. The literature suggests that PBM may be a viable component in immunotherapy even for cancer.[43] Of relevance to this discussion, the fact that PBM strengthens the innate immune system makes it a credible prophylaxis against diseases, which include viral infections.

Viruses have the propensity to mutate, and we could be potentially facing a "COVID-20 (and beyond)" after this outbreak. Hence the need to continuously support the innate immune system, which in turn also helps with improved adaptive immune response in readiness for the next viral infection. PBM's support of the body's innate immunity, inherently provides a simple solution. The X-Plus offers a safe instrument to deliver this.

The usefulness of the intranasal PBM treatment is underlined by new evidence that the loss of smell, or anosmia, and taste are characteristic of about 20 to 59 percent of cases of early COVID-19 infection - making this amongst the strongest predictors of the disease.[44] [45] [46] As discussed, the X-Plus intranasal PBM applicator through NO release, may be able to slow down or stop a viral invasion by inhibiting its replication at the nasal cavity before progressing further into the respiratory tracts.[31]

In summary, the role of intranasal and trans-sternal PBM in prophylaxis for the disease is its potential to improve the body's innate and adaptive immunity, and to halt viral replication.

**Conclusion**

In conclusion, several factors support the proposal that the Vielight X-Plus is a viable consideration to address COVID-19.

PBM is supported by a growing amount of research and the evidence that is deep and credible, going back about 55 years.

In relation to the X-Plus:

- It is readily available, with no development time required
- It is commercially available as a general wellness device with a 2-year track record
- Positive anecdotal reports of improved well-being have been received with no safety issues
- The development of the device is based on photobiomodulation (PBM), specifically related to improving the immune constitution
- It is low-cost, portable, does not require training to use and is suitable as a home-use device
- Its design is based on established scientific principles, supported by related empirical evidence.

©2020 Vielight Inc.
All rights reserved

However, there are certain qualifications:

- Its efficacy needs to be validated in a clinical study
- Although PBM is rapidly gaining scientific recognition, many authorities remains skeptical, which

  can be corrected with education

- Volume production would require a lead time of two months.

The device warrants a clinical trial. Notwithstanding, the X-Plus present a promising non-invasive solution to attack the COVID-19 virus. This is supported by a body of related literature, history in the market and safety.

[46] Loss of smell and taste is strongest predictor of early COVID-19 infection. Neuroscience News. April 1, 2020.

https://neurosciencenews.com/covid-19-smell-taste-16056/. Accessed April 1, 2020.

©2020 Vielight Inc.
All rights reserved

#### Table 1.3: Table of Amendments to Protocol

| Description of Change | Brief Rationale | Section(s) |
|---|---|---|
| A. Time to elimination of symptoms in days on the following items in the WURSS-44, where elimination of symptoms is defined as a rating of 0 (do not have this symptom or not at all) that is confirmed over 3 consecutive days:<br>o Item 2 “Cough”<br>o Item 12 “Body aches”<br>o Item 18 “Feeling tired”<br>o Item 37 “Breathe easily”<br>o Item 16 “Feeling feverish”<br><br>B. Mean number of days with a rating of 0, 1, 2 or 3 for the following items:<br>o Item 1 “How sick do you feel today”<br>o Item 2 “Cough”<br>o Item 12 “Body aches”<br>o Item 18 “Feeling tired”<br>o Item 37 “Breathe easily”<br>o Item 16 “Feeling feverish”<br><br>C. Mean number of days with mild overall respiratory symptoms as measured by a total WURSS-44 score of <=129<br><br>D. Time to reduction in symptoms in days as measured by item 1 of the WURSS-44 “How sick do you feel today”. Reduction in symptoms is defined as a rating of 3 or less that is confirmed over 3 days.<br><br>E. Time to elimination of symptoms in days on the following items in the WURSS-44, where elimination of symptoms is defined as a rating of 0 (do not have this symptom or not at all) that is confirmed over 3 consecutive days:<br>o Item 16 “Feeling feverish”<br>o Item 37 “Breathe easily”<br><br>F. Average number of days spent with mild respiratory symptoms for the 7 questions relating to respiratory symptoms defined below (WURSS-44 respiratory score < or = 21) in treatment group compared to SOC. WURSS-44 rating on past 24hrs for<br>o ‘cough’,<br>o ‘coughing stuff up’,<br>o ‘cough interfering with sleep’, | It is more useful to cover all the available WURSS-44 items, #2-23, as secondary endpoints. | 1.0 Secondary endpoints<br><br>and<br>7.2 Secondary endpoints (repeated) |

| | | |
|---|---|---|
| o ‘chest congestion’,<br>o ‘chest tightness’,<br>o ‘heaviness in chest’<br>o ‘breathe easily’<br><br>**Amended to:**<br><br>“A. Time to elimination of symptoms in days for items #2-43 on WURSS-44, where elimination of symptoms is defined as a rating of 0 (do not have this symptom or not at all) that is confirmed over 3 consecutive days.<br><br>B. Mean number of days with a rating of 0, 1, 2 or 3 for items #2-43 on WURSS-44.<br><br>C. Mean number of days with mild overall respiratory symptoms as measured by a total WURSS-44 score of <=129<br><br>D. Time to reduction in symptoms in days as measured by item 1 of the WURSS-44 “How sick do you feel today”. Reduction in symptoms is defined as a rating of 3 or less that is confirmed over 3 days.” | | |
| The following endpoint assesses a time-to-event, and will be assessed as described for the primary endpoint, utilizing a Log-rank test:<br>Time to elimination of symptoms in days on the following items in the WURSS-44, where elimination of symptoms is defined as a rating of 0 (do not have this symptom or not at all) that is confirmed over 3 days:<br>o Item 2 “Cough”<br>o Item 12 “Body aches”<br>o Item 18 “Feeling tired”<br>o Item 37 “Breathe easily”<br>o Item 16 “Feeling feverish”<br><br>**Amended to:**<br><br>“Time to elimination of symptoms in days for items #2-43 on WURSS-44, where elimination of symptoms is defined as a rating of 0 (do not have this symptom or not at all) that is confirmed over 3 consecutive days.” | It is more useful to cover all the available WURSS-44 items, #2-23, as secondary endpoints. | 8.4.2 |
