## Supplementary material for "Home-use Photobiomodulation Device Treatment Outcomes for COVID-19": Treatment Device Specifications

### Supplement 2

#### Treatment Device, Vielight RX Plus Specifications

**Concept of RX Plus design:**

1. A movable 810 nm LED module can be positioned on the thymus gland and sternum at the same time.
2. An intranasal diode of visible red 633 nm wavelength added to introduce light into the nasal cavity.

**Movable LED module applicator:**

Wavelength: 810 nm, non-laser

Waveform: continuous wave

Beam spot size upon contact ≈ 1 cm2

Treatment time: 20 minutes

Power output density on the skin surface = 50 mW/cm

Energy dose density on skin surface = 50*20*60/1000 = 60 J/cm2

**Nasal applicator LED:**

Wavelength: 633 nm

Waveform: continuous wave

Beam spot size upon contact ≈ 1 cm2

Treatment time: 20 minutes

Power output density to mucosa = 6.5 mW/cm2 .

Energy dose density to mucosa = 8*20*60/1000 = 9.6 J/cm2

**Total energy dose to surface tissues: 60+9.6 = 69.6 J/cm2**

Dated: June 1, 2020
