## Supplementary material for "Home-use Photobiomodulation Device Treatment Outcomes for COVID-19": Statistical Analysis Plan and Methods

### Supplement 3

### Statistical Analysis Plan and Statistical Methods

**Table of Contents**

3.1 Notes to the Statistical Analysis Plan

3.2 Statistical Analysis Plan

3.3 Table of Amendments to Statistical Analysis Plan

3.4 Statistical Methods Used

#### Supplement 3.1

#### Notes to the Statistical Analysis Plan

The Statistical Analysis Plan (SAP) reflects the amendments made to the Protocol (Supplements 1.1-1.3).

The SAP (Supplement 3.2) should be read in conjunction with the Amendments (Supplement 3.3). The main amendments relate to expanding analyses beyond the original few named items of WURSS-44 items:

- Item 2 “Cough”
- Item 12 “Body aches”
- Item 18 “Feeling tired”
- Item 37 “Breathe easily”
- Item 16 “Feeling feverish”

This would provide more useful information based on all available items as secondary endpoints.

Adverse events reporting (Section 9 of Supplement 3.2) has been amended from basing on event-frequency to patient-frequency, also giving emphasis to the usefulness of the information.

The primary endpoint would remain unaffected.

Supplement 3.2

### Statistical Analysis Plan

*A RANDOMIZED STUDY EVALUATING THE EFFICACY OF THE VIELIGHT RX PLUS IN THE TREATMENT OF COVID-19 RESPIRATORY SYMPTOMS*

| | |
|---|---|
| **Investigational Device Name:** | Vielight RX Plus |
| **SAP Version:** | 2.1 |
| **SAP Version Date:** | 2020-12-09 |
| **Trial Statistician:** | Conrad Kabali |
| **Protocol Version:** | 2.0 |
| **Trial Sponsor:** | Vielight Inc. |

#### Signatures

I give my approval for the attached statistical analysis plan (SAP) entitled "*A Randomized Study Evaluating the Efficacy of the Vielight Rx Plus in the Treatment of Covid-19 Respiratory Symptoms*" dated 2020-12-09.

**Statistician (Author)**

Name: Conrad Kabali

Signature: CKabali

Date: 2020-12-09

**Trial Sponsor**

Name: Nazanin Hosseinkhah, PhD

Signature: Nazanin. H

Date: Dec 10, 2020

#### Table of Contents


1 Introduction 2
1.1 Scope of the analyses 2
2 Study Objectives and Endpoints 2
2.1 Study Objective 2
2.2 Endpoints 2
3 Study Methods 3
3.1 General Study Design and Plan 3
3.2 Inclusion-Exclusion Criteria and General Study Population 4
3.3 Randomization 4
4 Sample Size 5
4.1 Final Analysis 5
4.2 Analysis Populations 5
4.3 Covariates and Subgroups 5
4.4 Missing Data 5
4.5 Interim Analysis 6
5 Summary of Study Data 6
5.1 Protocol Deviations 7
5.2 Demographic and Baseline Variables 7
6 Primary Efficacy Analysis 8
7 Secondary Efficacy Analyses 8
7.1 Outcomes based on mean number of days 8
7.2 Outcomes based on time-to-event 8
8 Descriptive Efficacy Analyses 9
9 Safety Analyses 9
10 Reporting Conventions 10
11 References 10

### 1 Introduction

The Vielight RX Plus is a home-use photobiomodulation device which is hypothesized to inhibit nasal coronavirus replication, elevate immune cell function, control inflammation, and thus act to accelerate recovery and reduce viral infection morbidity. It involves the application of red (600-700 nm) or near-infrared (760-1200 nm) light to modulate body functions.

This unblinded randomized controlled study will investigate the efficacy and safety of the Vielight RX Plus in treating Covid-19 symptoms. The study is expected to take a duration of 6 months, covering the period from the date of first subject enrollment to completion of final participant. The maximum follow-up period per subject will be 30 days since treatment initiation. Respiratory symptoms will be self-reported using WURSS-44 questionnaire.

#### 1.1 Scope of the Analysis

Analysis will assess the efficacy and safety of Vielight RX Plus + standard of care (SOC) versus SOC alone for the treatment of Covid-19 symptoms. It will cover: evaluation of baseline distribution characteristics to characterize the study population and assess imbalance in baseline covariates, evaluation of efficacy of Vielight RX Plus + SOC on the primary endpoint using survival analyses, comparisons of  means or medians of various secondary endpoints between treatment arms,  and comparisons of rates of adverse evets between the two treatments. An interim analysis will be conducted after recruiting the first 73 participants. The aim of the interim analysis will be to determine if the trial should be stopped early due to any of the following reasons: treatment is clearly efficacious, treatment is clearly harmful, or treatment is obviously futile.

### 2 Study Objectives and Endpoints

#### 2.1 Study Objective

The objective of this study is to investigate the efficacy and safety of the Vielight RX Plus as an adjunct to SOC compared with SOC alone in decreasing time to recovery of symptoms in subjects with COVID-19.

#### 2.2 Endpoints

The primary endpoint will be

- Time to overall recovery in days as measured by item 1 of the WURSS-44 “How sick do you feel today”. Recovery is defined as a rating of 0 (not sick) that is confirmed over 3 consecutive days.

The secondary endpoints will include

- Time to elimination of symptoms in days on the following items in the WURSS-44, where elimination of symptoms is defined as a rating of 0 (do not have this symptom or not at all) that is confirmed over 3 consecutive days:
    - Item 2 “Cough”

- Item 12 “Body aches”
- Item 18 “Feeling tired”
- Item 37 “Breathe easily”
- Item 16 “Feeling feverish”

- Mean number of days with a rating of 0, 1, 2 or 3 for the following items:
  - Item 1 “How sick do you feel today”
  - Item 2 “Cough”
  - Item 12 “Body aches”
  - Item 18 “Feeling tired”
  - Item 37 “Breathe easily”
  - Item 16 “Feeling feverish”
- Mean number of days with mild overall respiratory symptoms as measured by a total WURSS-44 score of <=129
- Time to reduction in symptoms in days as measured by item 1 of the WURSS-44 “How sick do you feel today”. Reduction in symptoms is defined as a rating of 3 or less that is confirmed over 3 days
- Time to elimination of symptoms in days on the following items in the WURSS-44, where elimination of symptoms is defined as a rating of 0 (do not have this symptom or not at all) that is confirmed over 3 consecutive days:
  - Item 16 “Feeling feverish”
  - Item 37 “Breathe easily”
- Average number of days spent with mild respiratory symptoms for the 7 questions relating to respiratory symptoms defined below (WURSS-44 respiratory score ≤21) in treatment group compared to SOC. WURSS-44 rating on past 24hrs for
  - ‘cough’,
  - ‘coughing stuff up’,
  - ‘cough interfering with sleep’,
  - ‘chest congestion’,
  - ‘chest tightness’,
  - ‘heaviness in chest’
  - ‘breathe easily’
- Oxygen Saturation by Pulse Oximetry [Time Frame: 30 Days]
- Mortality rate in treatment group compared to SOC
- Hospitalization rate in Tteatment group compared to SOC
- Proportion of subjects with serious adverse events in treatment group compared to SOC

### 3 Study Methods

#### 3.1 General Study Design and Plan

This will be a randomized, unblinded, parallel-group, superiority study. Subjects will be randomized to receive treatment which is comprised of Vielight RX Plus + SOC or control which consists of SOC alone. The minimization method will be used to randomize participants. The randomization schedule will be generated by the Contract Research Organization. A diagram of study flow is shown in Figure 1.

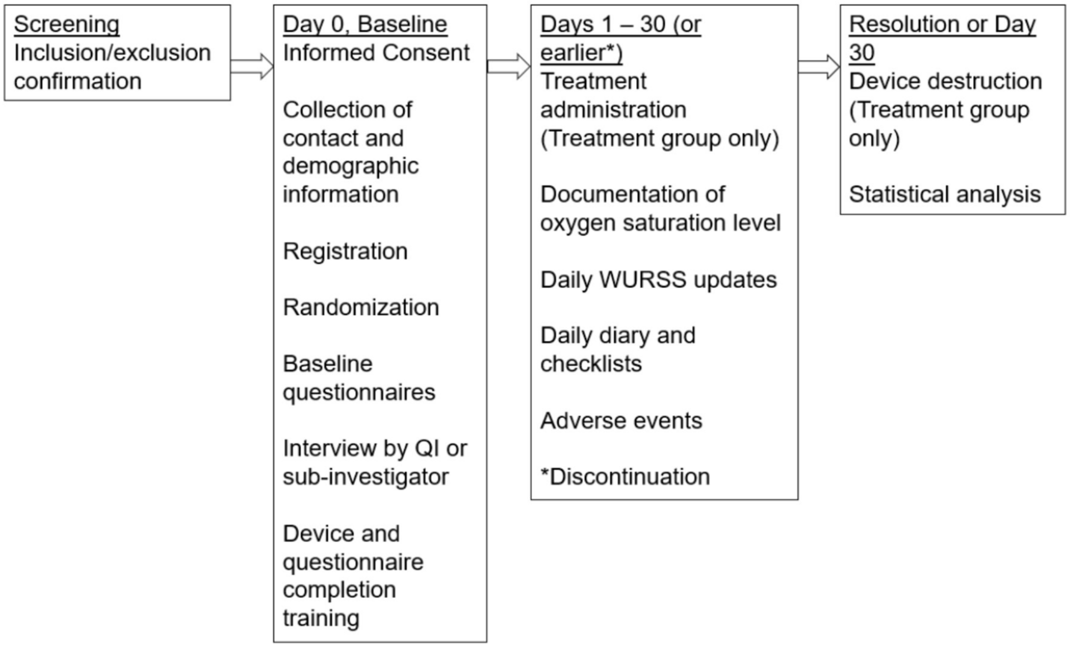


Figure 1. Study flow chart

#### 3.2 Inclusion-Exclusion Criteria and General Study Population

Inclusion criteria will be
- Confirmation of COVID-19 infection
- Score of 4-7 for WURSS-44 Question 1 (moderate to severe sickness)
- Between 18-65 years of age
- No other household member already recruited

Exclusion criteria will be
- Need for hospitalization at the time of diagnosis
- Current need for supplemental oxygen or positive pressure support and/or has required supplemental oxygen or positive pressure support for >or= 24 hours
- Greater than 10 days since symptom onset
- Diagnosis of Chronic Obstructive Pulmonary Disease (COPD)
- Pregnant
- Positive for Hepatitis C Virus (HCV), Hepatitis B Virus (HBV) or Human Immunodeficiency Virus
- Inability to electronically complete WURSS-44 daily (in English)

#### 3.3 Randomization

Study staff will complete a randomization form using OxMAR minimization software. Participants will be randomly allocated to the treatment group or SOC group (1:1) based on the minimization algorithm which aims to reduce the differences in the two groups based on baseline factors (i.e.

duration of days since symptom onset (≤ 5 or 6-10 days) and WURSS Q1 score (4-5 or 6-7). Once baseline information has been entered into the OxMAR minimization software, study staff will be notified which arm the participant has been allocated to via email.

Participants will receive a notification from the electronic data capture system regarding group assignment and details of the study requirements. Each participant will also be provided with a link to a secure private portal unique to them. All questionnaires, diaries and checklists will be available in the portal for baseline and daily access.

### 4 Sample Size

The study will enroll 280 patients in a 1:1 randomization. If, as expected, nearly all patients should recover within the 30-day treatment period, then a hazard ratio of 1.40 for the primary endpoint can be detected with approximately 80% power with a two-sided significance level of 5%. If a substantial proportion of patients (i.e. approximately 15%) do not record a recovery within the 30-day treatment period, then a hazard ratio of 1.44 can be detected with approximately 80% power with a two-sided significance level of 5%.

#### 4.1 Final Analysis

The final analysis will be performed when all patients have completed treatment, regardless of the number of recovery events observed.

#### 4.2 Analysis Populations

Endpoints will be assessed in two study populations, the per protocol (PP) and intention-to-treat (ITT) population. All participants who complete all study procedures per protocol will be included in the PP analysis. Participants who have protocol deviations will be reported on the appropriate Deviations Log. ITT population will include all enrolled participants.

#### 4.3 Covariates and Subgroups

If a notable imbalance in treatment allocation is observed for the following covariates: Duration of Days Since Symptom Onset and WURSS Q1 score, a stratified Cox proportional hazards model will be adjusted for these covariates. Other baseline covariates such as pre-existing health conditions, medication use, race/ ethnicity, and sex will also be considered for inclusion in the Cox model, if their inclusion will not negatively impact the precision of efficacy estimates.

#### 4.4 Missing Data

For transient missing data, investigators will follow-up with participants to gather the actual values for the empty fields. The missing values due to dropouts will be handled at the analysis stage. The type of analysis for handling dropouts will depend on the assumptions underlying the missingness mechanism. If the dropouts are assumed to be missing completely at random (i.e. they are a random sample of subjects who are still in the study), the expected-maximization algorithm[1] will be used to impute missing values. Conversely, if the dropouts are assumed to be missing at random (i.e. they are a random sample of subjects who are still in the study, conditional on the observed values of these subjects), the multiple imputations by chain equations will be applied.[2] However, if the missingness mechanism is not at random (i.e. if the reason for dropping out can only be explained by dropouts themselves), no imputation will be done, instead the missing data problem will be discussed under study limitations. The extent of missing data will be summarized in a table.

#### 4.5 Interim Analysis

A single interim analysis will be conducted after recruiting the first 73 participants. To preserve the overall type I error rate at the nominal level of 0.05, the stopping boundary for efficacy will be set at $p<0.0051$.[3] The stopping boundary for futility will be set at the conditional power[4] of 50%.

### 5 Summary of Study Data

All continuous variables will be summarized using the following descriptive statistics: n (non-missing sample size), mean and standard deviation (for normally distributed measurements), median and interquartile range (for skewed measurements), maximum, and minimum. The frequency and percentages (based on the non-missing sample size) of observed levels will be reported for all categorical measures. In general, all data will be listed, sorted by site, treatment, and subject, and by visit number within subject. All summary tables will be structured with a column for each treatment in the order (Vielight Rx Plus, Standard of Care) and will be annotated with the total population size relevant to that table/treatment, including any missing observations.

The CONSORT flow diagram (Figure 2) will be used to provide a summary of the filtering process for analytic data.

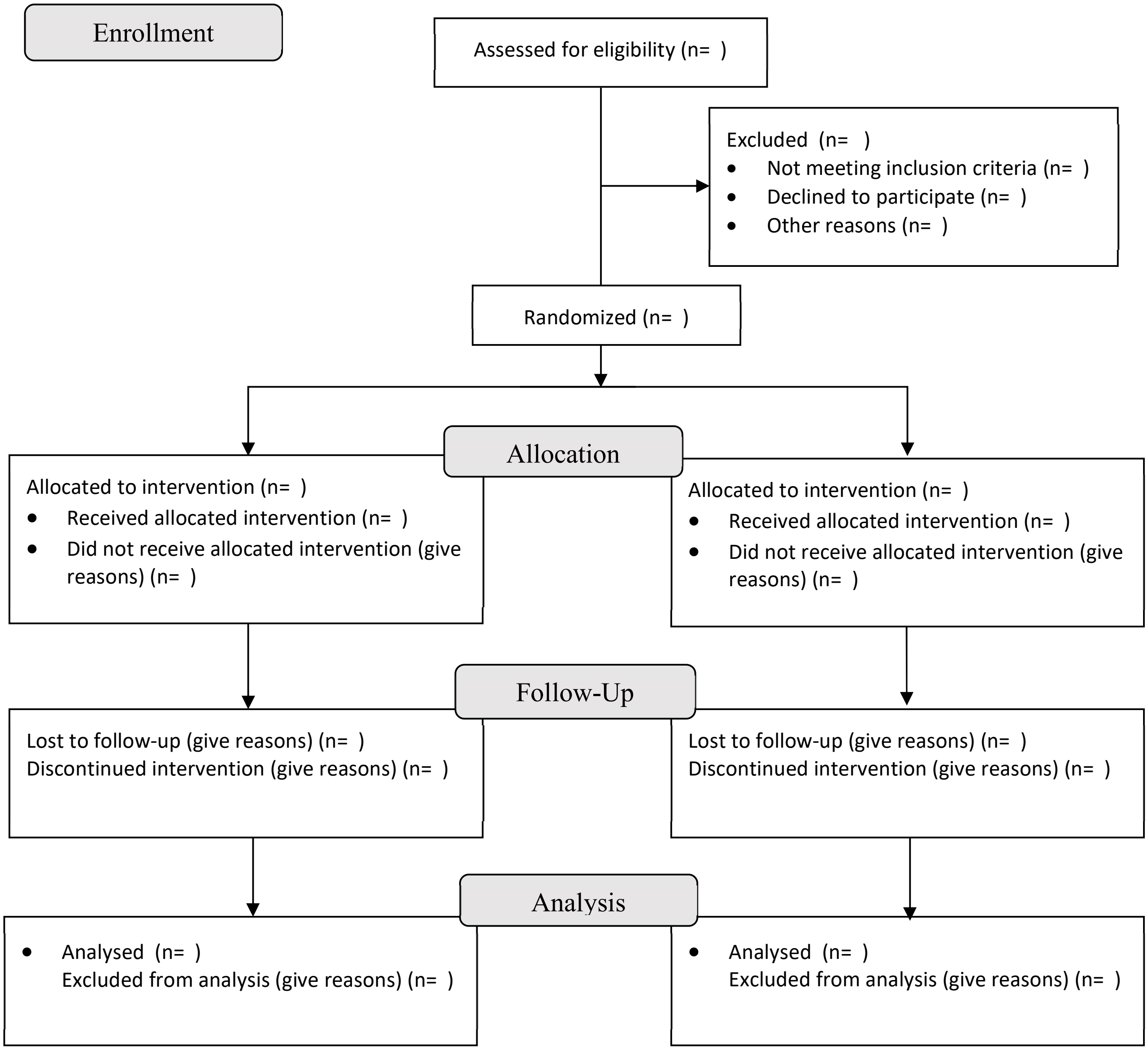


Figure 2. CONSORT flow diagram

#### 5.1 Protocol Deviations

Since this trial is not blinded, it is possible for participants living in the same social circle but randomized to different arms to switch treatments, thereby contaminating the results. To ameliorate this problem, only one person per household will be recruited.

#### 5.2 Demographic and Baseline Variables

The distribution of the following baseline and demographic variables will be reported: pre-existing health conditions, medication use, WURSS-44, Fitzpatrick scale, age, race/ ethnicity, and sex.

### 6 Primary Efficacy Analysis

Time-to-event is defined as the interval from the date of first use of the device to the start date of the improvement. Three consecutive days of reporting 0 on Question 1 of the WURSS-44 is considered durable improvement. The start date of the improvement will be taken to be the first day where the criteria is met. Subjects who have not met the definition of improvement will be censored as of their last available assessment. Subjects without a baseline assessment or post-baseline assessment will be censored at the date of randomization.

Time-to-improvement will be estimated using Kaplan-Meier methods. The proportion of subjects with an improvement will be reported by treatment group. The overall group comparison will be done using a log-rank test stratified by time since onset of symptoms (≤ 5 days vs. 6-10 days). The hazard ratio and its associated 95% confidence interval will be calculated based on a stratified Cox proportional hazards model.

### 7 Secondary Efficacy Analyses

#### 7.1 Outcomes based on mean number of days

Outcomes based on mean number of days will be computed in two ways. The first method will utilize all available assessments. The second method will assume a 30-day follow-up and will consider any missing assessment as not having met the criteria.

The mean number of days meeting a certain criterion will be analyzed using an ANCOVA model with a fixed effect for treatment group assignment and a covariate of time since onset of symptoms (≤ 5 days vs. 6-10 days).

- Item 2 “Cough”
- Item 12 “Body aches”
- Item 18 “Feeling tired”
- Item 37 “Breathe easily”
- Item 16 “Feeling feverish”

#### 7.2 Outcomes based on time-to-event

The following endpoint assesses a time-to-event, and will be assessed as described for the primary endpoint, utilizing a Log-rank test: time to elimination of symptoms in days on the following items in the WURSS-44, where elimination of symptoms is defined as a rating of 0 (do not have this symptom or not at all) that is confirmed over 3 days:

- Item 2 “Cough”
- Item 12 “Body aches”
- Item 18 “Feeling tired”
- Item 37 “Breathe easily”
- Item 16 “Feeling feverish”

#### 8 Descriptive Efficacy Analyses

Secondary endpoints assessing between group proportions will be assessed via a chi-square test. Participant demographic, and baseline measurements will be presented in contingency tables as mean, standard deviation. Baseline values, post-treatment values, and change from baseline to post-treatment will be compared between the groups using independent samples t-test, with statistical significance at $p < 0.05$, and 95% confidence intervals reported.

#### 9 Safety Analyses

The commercial version of the RX Plus has been commercially available over 2 years, with no report of any major side effects, other than the occasional report of a warm feeling from the 810 nm LED targeted for placement over the manubrium. It has the same controller and driver platforms as the Vielight Neuro RX Gamma which has been tested for safety and approved for a clinical trial by Health Canada. Unrelated to safety, the other factor that may arise is a malfunction of the device, in which case the CRO will liaise with the Sponsor who will take corrective action. Notwithstanding the anticipated absence of major issues, a mechanism is in place for the subjects to report an adverse event to the CRO, who will document it and report to the Sponsor for further corrective action.

A device-related event is determined by the "relatedness score" for Adverse Events. A relatedness score of "Possible" or "Probably" or "Definite" indicate the extent that the Adverse Event is related to the use of the device. Adverse Events receiving a relatedness score of "Unrelated" or "Unlikely" are considered unrelated to the device use and are not included. All safety endpoints will be adjudicated by one of the study administrators.

A statistical comparison of the incidence rates will be conducted. Additionally, an evaluation of any events due to the general nature of the device (i.e. headset discomfort, nasal irritation) and not specific to the delivery of the NIR energy will be qualitatively evaluated. All emergent findings from participant reported oxygen saturation will be documented and reviewed by the QI to complete an AE form and update the AE log if required.

Three tables summarizing adverse events will be reported:

- All-Cause Mortality: A table of all anticipated and unanticipated deaths due to any cause, with number and frequency of such events in each arm will be reported.

- Serious Adverse Events (events that are serious enough to require hospitalization): A table of all anticipated and unanticipated serious adverse events, grouped by organ system, with number and frequency of such events in each arm will be reported.

- Other (Not Including Serious) Adverse Events: A table of anticipated and unanticipated events (not included in the serious adverse event table) that exceed a frequency threshold of 5 % within any arm, grouped by organ system, with number and frequency of such events in each arm will be reported.

#### 10 Reporting Conventions

The p-values ≥0.001 will be reported to 3 decimal places; p-values less than 0.001 will be reported as "<0.001". The mean, standard deviation, and any other statistics other than quantiles, will be reported to one decimal place greater than the original data. Quantiles, such as median, or minimum and maximum will use the same number of decimal places as the original data. Estimated parameters, not on the same scale as raw observations (e.g. regression coefficients) will be reported to 3 significant figures.

#### Supplement 3.3

#### Table of Amendments to Statistical Analysis Plan

| Description of Change | Brief Rationale | Section(s) |
|---|---|---|
| "Time to elimination of symptoms in days on the following items in the WURSS-44, where elimination of symptoms is defined as a rating of 0 (do not have this symptom or not at all) that is confirmed over 3 consecutive days:<br>o Item 2 "Cough"<br>o Item 12 "Body aches"<br>o Item 18 "Feeling tired"<br>o Item 37 "Breathe easily"<br>o Item 16 "Feeling feverish"<br>Amended to:<br>"Time to elimination of symptoms in days for items #2-43 on WURSS-44, where elimination of symptoms is defined as a rating of 0 (do not have this symptom or not at all) that is confirmed over 3 consecutive days." | It is more useful to cover all the available WURSS-44 items, #2-23, as secondary endpoints. | 2.2 |
| "Mean number of days with a rating of 0, 1, 2 or 3 for the following items:<br>o Item 1 "How sick do you feel today"<br>o Item 2 "Cough"<br>o Item 12 "Body aches"<br>o Item 18 "Feeling tired"<br>o Item 37 "Breathe easily"<br>o Item 16 "Feeling feverish""<br><br>Amended to:<br>"Mean number of days with a rating of 0, 1, 2 or 3 for items #2-43 on WURSS-44." | It is more useful to cover all the available WURSS-44 items, #2-23, as secondary endpoints. | 2.2 |
| "Time to elimination of symptoms in days on the following items in the WURSS-44, where elimination of symptoms is defined as a rating of 0 (do not have this symptom or not at all) that is confirmed over 3 consecutive days:<br>o Item 16 "Feeling feverish"<br>o Item 37 "Breathe easily""<br><br>Amendment:<br>Delete this section | This is already covered above under "Time to elimination…" | 2.2 |
| "Average number of days spent with mild respiratory symptoms for the 7 questions relating | This is already covered above under "Mean number of days…." | 2.2 |

| | | |
|---|---|---|
| to respiratory symptoms defined below (WURSS-44 respiratory score ≤21) in treatment group compared to SOC. WURSS-44 rating on past 24hrs for<br>o ‘cough’,<br>o ‘coughing stuff up’,<br>o ‘cough interfering with sleep’,<br>o ‘chest congestion’,<br>o ‘chest tightness’,<br>o ‘heaviness in chest’<br>o ‘breathe easily’”<br><br>Amended to:<br>Delete this section. | | |
| Added to Missing Data section:<br>“In the use of the Kaplan-Meier Method, missing data need not be imputed.” | Kaplan-Meier estimates account for variable follow-up time under the assumption of non-informative censoring. | 4.4 |
| The mean number of days meeting a certain criterion will be analyzed using an ANCOVA model with a fixed effect for treatment group assignment and a covariate of time since onset of symptoms (≤ 5 days vs. 6-10 days).<br>• Item 2 “Cough”<br>• Item 12 “Body aches”<br>• Item 18 “Feeling tired”<br>• Item 37 “Breathe easily”<br>• Item 16 “Feeling feverish”<br><br>Amended to:<br>“The mean number of days with mild symptoms for WURSS-44 items #1-43, will be analyzed using an ANCOVA model with a fixed effect for treatment group assignment and a covariate of time since onset of symptoms (≤ 5 days vs. 6-10 days).” | This refers to the number of days with mild symptoms.<br>It is more useful to cover all the 43 available WURSS-44 symptom items. | 7.1 |
| “The following endpoint assesses a time-to-event, and will be assessed as described for the primary endpoint, utilizing a Log-rank test: time to elimination of symptoms in days on the following items in the WURSS-44, where elimination of symptoms is defined as a rating of 0 (do not have this symptom or not at all) that is confirmed over 3 days:<br>• Item 2 “Cough” | It is more useful to cover all the 43 available WURSS-44 symptom items. | 7.2 |

| | | |
|---|---|---|
| • Item 12 “Body aches”<br>• Item 18 “Feeling tired”<br>• Item 37 “Breathe easily”<br>• Item 16 “Feeling feverish””<br><br>Amended to:<br>“In assessing the time-to-event endpoints for all WURSS-44 items, the Log-rank test will be used: time to elimination is defined as a rating of 0 (do not have this symptom or not at all) that is confirmed over 3 days” | | |
| Three tables summarizing adverse events will be reported:<br>● All-Cause Mortality: A table of all anticipated and unanticipated deaths due to any cause, with number and frequency of such events in each arm will be reported.<br>● Serious Adverse Events (events that are serious enough to require hospitalization): A table of all anticipated and unanticipated serious adverse events, grouped by organ system, with number and frequency of such events in each arm will be reported.<br>● Other (Not Including Serious) Adverse Events: A table of anticipated and unanticipated events (not included in the serious adverse event table) that exceed a frequency threshold of 5 % within any arm, grouped by organ system, with number and frequency of such events in each arm will be reported.<br><br>Amended to:<br>Three tables summarizing adverse events will be reported for Treatment/Control arms (monitored from enrollment after follow-up but regardless of WURSS-44 severity scores at Baseline):<br>• All-Cause Mortality: A table of all anticipated and unanticipated deaths, listing the diagnoses on expiration.<br>• Serious Adverse Events requiring hospitalization (excluding deaths): A table of all anticipated and unanticipated listing of patients, citing the main symptoms.<br>• Non-serious adverse events: A table of anticipated and unanticipated listing of patients that exceed 5% of total of monitored patients. | For usefulness, the adverse events are reported by the number of patients that experience each symptom, belonging to Treatment or Control.<br>Frequency of events are not as useful because of the risk of overrepresentation of events that affect few people. | 9 |

#### Supplement 3.4

#### Statistical Methods Used

The aim of the methods deployed was to support the primary and secondary endpoints stated in the clinical trial protocol (Supplement 1) as well as other supplemental statistical analyses.

The primary and secondary endpoints were based on questions 1 to 43 of the Wisconsin Upper Respiratory Severity Survey (WURSS) -44 set of questions. Question 44, "Compared to yesterday, I feel my cold is…" was not used as it was deemed not applicable.

Summaries of salient findings are stated in the main text.

Further details of the statistical methods used in the study are consolidated in the discussion here. They can be broadly sectioned as follows:

**Meeting the primary endpoint of time-to-recovery in answer to the WURSS question of "How sick do you feel today?" as the Primary Outcome**

- Kaplan-Meier Method[1]
- Cox Proportional Hazards Model[2]

**Meeting the secondary endpoints which involve time-to-recovery and days with symptoms listed by the WURSS-44**

- Kaplan Meier Method for time-to-recovery[1]
- Cox Proportional Hazards Model for time-to-recovery[2]
- Analysis of Variance for days with mild symptoms

**Oxygen Saturation**

- Mixed Model Repeated Measures Analysis of Covariance Treatment[3], comparing Oxygen Saturation (%) in the Change from Baseline

**Adverse Events**

- Poisson Regression Model[4]

**Comments on the statistical methods used**

<u>Kaplan-Meier Method</u>

The time-to-recovery estimates for the primary efficacy outcome were assessed with the Kaplan-Meier (KM) method[1], and comparisons between Treatment and Control groups were assessed with the log-rank test, stratified by symptom duration days (0-5, 6-10). An unstratified log-rank test was utilized for statistical analysis within symptom duration strata. This method was used for both the primary and secondary outcomes for time-to-event assessments.

"Time-to-recovery" is defined as the interval (days) from the date of first use of the device to the start date of a '0' (no symptoms) WURSS-44 score for 3 consecutive days, in subjects with baseline WURSS-44 score of 4-7. Subjects who did not meet the definition of recovery were censored as of their last available assessment on day 30. Censoring also included subjects who did not complete 30 days and those with less than 3 consecutive days of no symptoms at the end of the 30-day assessment.

The Treatment group data collection started on the day of "first use" of the treatment device ("Day 1"), which had had mean 1.9 (median 2.0) in Treatment and 1.8 (median 2.0) in control. The delay for first-use of the device was largely due to shipping. After adjusting for a median 2-day delay from shipping to device use, symptoms duration category 0-5 days is interpreted as 0-7 days, and symptom duration 6-10 day category is interpreted as 8-12 days from the start of symptoms.

A stratified and unstratified Log-rank test was applied to test the significance of the difference in recovery (median days at 95% confidence interval) between the Treatment group and the Control group.

The full table is presented in Supplement 6.3.

Cox Proportional Hazards Model

The Cox Proportional Hazards Model[2] used here estimated the Treatment:Control hazard ratios and tested for significance between Treatment and Control over the 30-day assessment period, where hazard ratios (HR) greater than 1 would be in favor of Treatment.

The model was used to analyze WURSS-44 Q1 as the primary outcome as well as the 42 other questions listed (Supplement 7.1). Proportional hazards models were used to test outcomes between Treatment and Control by subgroups of Demographics and Characteristics (Supplement 7.2).

Analysis of Variance

The least-squares mean for the number of days spent with mild symptoms (0-3 on the WURSS-44 severity scores) was estimated. Model terms include treatment and symptom duration strata (0-5 days, 6-10 days on enrollment). For analysis by symptom duration strata, a one-way ANOVA was used. See Supplement 8 for the full table.

Mixed Model for Repeated Measures

To analyze the percentage changes in oxygen saturation, we used the Mixed Model Repeated Measures (MMRM) Analysis of Covariance[3]. Model terms include treatment, day at weekly intervals, treatment-by-day interaction, symptom duration strata (0-5 days, 6-10 days on enrollment), and

Baseline/Screening covariate. This was carried out for all reported data at Screening and Baseline. See Supplement 9.1 for the results.

Poisson Regression Model

Based on the frequency distributions of adverse events presented, a Poisson regression model[4] was used to compare the mean number of episodes of the adverse event (AE) and patients with AEs between Treatment and Control. See Supplements 10.1 and 10.2.
