## Supplementary material for "Home-use Photobiomodulation Device Treatment Outcomes for COVID-19": Patient Demographics, Information

### Supplement 4

### Patient Information

**Table of Contents**

4.1 Chronology, Recruitment Numbers and Baseline Discussion

4.2 Full Table of Patient Demographics and Baseline Characteristics

4.3 Demographics and Baseline Characteristics of Patients with Baseline WURSS-44 Q1 Score of 4-7 and Summary Statistics for Days between Enrollment and Baseline

### Supplement 4.1

#### Chronology, Recruitment and Baseline Discussion

**Trial Chronology and Registrations**

Following early reports of the rapid spread of the COVID-19 pandemic in February 2020, we set out to develop a protocol for an RCT to test whether a home-use PBM device could be effective as a treatment for COVID-19. The device, the Vielight RX Plus, embodied as a general wellness device was already available at the time.

The trial protocol was submitted to the Medical Device Directorate of Health Canada on May 12, 2020, for Investigational Testing clearance, and was duly authorized to proceed on September 18, 2020 (Application No. 315768). It also received approval from Advarra, an institutional review board on May 22, 2020. Information was posted on NIH National Library of Medicine website on June 2, 2020 (ClinicalTrials.gov Identifier: NCT04418505).

We started the clinical trial recruitment on September 20, 2020, with the target of recruiting 280 patients, giving a hazard ratio of 1.4 in a successful trial. After collecting 73 datasets, we conducted an interim data analysis on January 15, 2021, to test the trial for superiority or futility. The trial continued as stopping criteria were not met.

As we approached the later part of the trial, in the summer of 2021, the dynamics of the standard of care (SOC) were quickly evolving that would adversely impact the baseline consistency. Recruitment of qualified patients was also becoming untenable. A large part of the reasons was the rapidly growing use of monoclonal antibodies for effective treatment of the dominant variant at the time, Delta. The monoclonal antibody treatments for COVID-19 patients, bamlanivimab[1] and casivimab plus imdevimab (REGEN-COV)[2] had been available since November 2020 in the United States. By the summer of 2021, the use of monoclonal antibodies was accelerating significantly in the treatment of newly diagnosed COVID-19 outpatients, changing the definition of the "standard of care". It was therefore disrupting consistent baseline data for fair comparisons between the earlier and later datasets. Furthermore, at around this time, CDC was sharing data that fully vaccinated people were experiencing milder symptoms, reducing potential candidates that would meet our inclusion criteria. And vaccinations were accelerating at that time. The homogeneity of the baselines of earlier and later participants was further affected. Given these factors, and to maintain the integrity of the trial, we decided this was a suitable time to stop recruitment and took the step to lock the 294 qualified datasets collected at that point, which was July 5, 2021. The number was deemed viable for credible statistical power. After the 30-day assessment period, we proceeded with the process of unblinding and data analyses on August 6, 2021.

**Recruitment and Baseline Adjustment**

We had screened 701 adults who were confirmed to have COVID-19 through RT-PCR testing. Out of these, 407 were excluded. 406 participants failed to meet the inclusion criterion of a score between 4 to 7 for Question 1 of the WURSS-44 scale, "How sick do you feel today" which would have indicated a moderate to severe level of sickness; or the participants were younger or older than the qualifying ages of between 18 to 65. 1 was unable to fill the forms in English, which was one of the exclusion criteria. That left 294 participants to agree to informed consent, briefing and training for clinical trial. Out of this, 142 received Treatment with the Vielight RX Plus device, and 152 received the standard of care (Control).

During follow-up from Enrollment and Allocation, a total of 27 patients were lost to withdrawal of consent, failure to provide or upload data, or they were disqualified by investigators for medical risks (see Figure). This left a total of 267 patients being monitored and assessed for safety with details reported in Supplements 9 & 10).

The Baseline of Treatment group patients was the first-use day ("Day 1") of the treatment device for the Treatment group. This required a 4-7 score for WURSS Q1 on "Day 1" of the treatment. The adjustment had a median (range 0 to 6) of 2 days due to shipping of the devices plus training. After further dropouts, there were 100 subjects in the Treatment group and 99 in the Control group. Statistical analysis of Demographics and Characteristics of Patients with Baseline WURSS-44 Q1 Score of 4-7, and Estimation of the Mean number of Days between Enrollment and Baseline are presented in Supplement 4.3. This adjustment did not affect the number of patients of 267 monitored and assessed for safety as this did not involve the time-to-event analyses.

The CONSORT flow chart for patient enrollment and treatment assignment is shown as the Figure of the main manuscript.

**Supplement 4.2**

#### Full Table of Patient Demographics and Baseline Characteristics

| | | | Treatment Group | | | |
|---|---|---|---|---|---|---|
| Variable | Statistic | Category | Treatment (N=100) | Control (N= 99) | Total (N=199) | P-Value |
| Sex | N (%) | Female | 31 (31.0) | 32 (32.3) | 63 (31.7) | 0.603 |
| | | Male | 68 (68.0) | 67 (67.7) | 135 (67.8) | |
| | | Unknown | 1 ( 1.0) | 0 | 1 ( 0.5) | |
| Age (years) | N | | 99 | 99 | 198 | 0.151 |
| | Mean (SD) | | 37.9 (13.16) | 35.3 (11.82) | 36.6 (12.54) | |
| | Median(IQR) | | 37 ( 22.4) | 34 ( 18.7) | 36 ( 20.7) | |
| | Min, Max | | ( 18, 66) | ( 19, 64) | ( 18, 66) | |
| Race | N (%) | AmerIndian/Alaskan | 2 ( 2.0) | 0 | 2 ( 1.0) | 0.434 |
| | | Black | 2 ( 2.0) | 5 ( 5.1) | 7 ( 3.5) | |
| | | Hawaiian/Islander | 4 ( 4.0) | 4 ( 4.0) | 8 ( 4.0) | |
| | | Unknown | 9 ( 9.1) | 12 (12.1) | 21 (10.6) | |
| | | White | 82 (82.8) | 78 (78.8) | 160 (80.8) | |
| Former Smoker | N (%) | No | 64 (64.0) | 61 (61.6) | 125 (62.8) | 0.612 |
| | | Unknown | 16 (16.0) | 21 (21.2) | 37 (18.6) | |
| | | Yes | 20 (20.0) | 17 (17.2) | 37 (18.6) | |
| Height (ins) | N | | 99 | 99 | 198 | 0.943 |
| | Mean (SD) | | 66.7 ( 3.63) | 66.6 ( 4.23) | 66.7 ( 3.93) | |
| | Median(IQR) | | 66.0 ( 5.0) | 66.0 ( 7.0) | 66.0 ( 5.0) | |
| | Min, Max | | (61.0, 75.0) | (58.0, 79.0) | (58.0, 79.0) | |
| Weight (lbs) | N | | 99 | 99 | 198 | 0.231 |
| | Mean (SD) | | 181.4 (54.24) | 173.0 (43.71) | 177.2 (49.31) | |
| | Median (IQR) | | 172.0 ( 69.0) | 165.0 ( 55.0) | 170.0 ( 60.0) | |
| | Min, Max | | ( 109, 411.0) | ( 110, 290.0) | ( 109, 411.0) | |

| | | | | | | |
|---|---|---|---|---|---|---|
| BMI (kg/m^2) | N | | 99 | 99 | 198 | 0.208 |
| | Mean (SD) | | 28.7 ( 8.26) | 27.3 ( 6.37) | 28.0 ( 7.39) | |
| | Median(IQR) | | 27.0 ( 9.2) | 26.2 ( 8.2) | 26.6 ( 8.6) | |
| | Min, Max | | (17.9, 65.7) | (19.1, 51.9) | (17.9, 65.7) | |
| Symptom Days Duration | N (%) | 0 to 7 | 68 (68.0) | 68 (68.7) | 136 (68.3) | 0.917 |
| | | 8 to 12 | 32 (32.0) | 31 (31.3) | 63 (31.7) | |
| WURSS Q1 Score at Baseline | N (%) | 4 | 32 (32.0) | 31 (31.3) | 63 (31.7) | 0.935 |
| | | 5 | 51 (51.0) | 48 (48.5) | 99 (49.7) | |
| | | 6 | 15 (15.0) | 17 (17.2) | 32 (16.1) | |
| | | 7 | 2 ( 2.0) | 3 ( 3.0) | 5 ( 2.5) | |
| Residency | N (%) | Canada | 8 ( 8.0) | 5 ( 5.1) | 13 ( 6.5) | 0.401 |
| | | United States | 92 (92.0) | 94 (94.9) | 186 (93.5) | |
| | N | | 100 | 99 | 199 | 0.696 |
| | Mean (SD) | | 1.9 ( 0.92) | 1.8 ( 0.94) | 1.9 ( 0.93) | |
| | Median(IQR) | | 2.0 ( 1.0) | 2.0 ( 1.0) | 2.0 ( 1.0) | |
| | Min, Max | | ( 0.0, 5.0) | ( 0.0, 6.0) | ( 0.0, 6.0) | |

*p<0.05 by two-sample t-test (continuous) and chi-square test (categorical). 17MAR22, T01_demog_mitt

#### Supplement 4.3

##### Demographics and Baseline Characteristics
##### In Patients with Baseline WURSS-44 Q1 Score of 4-7
##### and Summary Statistics for Days between Enrollment and Baseline

| | | | Treatment Group | | | |
|---|---|---|---|---|---|---|
| **Variable** | **Statistic** | **Category** | **Treatment (N=100)** | **Control (N= 99)** | **Total (N=199)** | **P-value** |
| Sex | N (%) | Female | 31 (31.0) | 32 (32.3) | 63 (31.7) | 0.603 |
| | | Male | 68 (68.0) | 67 (67.7) | 135 (67.8) | |
| | | Unknown | 1 ( 1.0) | 0 | 1 ( 0.5) | |
| Age (years) | N | | 99 | 99 | 198 | 0.151 |
| | Mean (SD) | | 37.9 (13.16) | 35.3 (11.82) | 36.6 (12.54) | |
| | Median(IQR) | | 37 ( 22.4) | 34 ( 18.7) | 36 ( 20.7) | |
| | Min, Max | | ( 18, 66) | ( 19, 64) | ( 18, 66) | |
| Race | N (%) | AmerIndian/Alask | 2 ( 2.0) | 0 | 2 ( 1.0) | 0.434 |
| | | Black | 2 ( 2.0) | 5 ( 5.1) | 7 ( 3.5) | |
| | | Hawaiian/Islande | 4 ( 4.0) | 4 ( 4.0) | 8 ( 4.0) | |
| | | Unknown | 9 ( 9.1) | 12 (12.1) | 21 (10.6) | |
| | | White | 82 (82.8) | 78 (78.8) | 160 (80.8) | |
| Former Smoker | N (%) | No | 64 (64.0) | 61 (61.6) | 125 (62.8) | 0.612 |
| | | Unknown | 16 (16.0) | 21 (21.2) | 37 (18.6) | |
| | | Yes | 20 (20.0) | 17 (17.2) | 37 (18.6) | |
| Height (ins) | N | | 99 | 99 | 198 | 0.943 |
| | Mean (SD) | | 66.7 ( 3.63) | 66.6 ( 4.23) | 66.7 ( 3.93) | |
| | Median(IQR) | | 66.0 ( 5.0) | 66.0 ( 7.0) | 66.0 ( 5.0) | |
| | Min, Max | | (61.0, 75.0) | (58.0, 79.0) | (58.0, 79.0) | |
| Weight (lbs) | N | | 99 | 99 | 198 | 0.231 |
| | Mean (SD) | | 181.4 (54.24) | 173.0 (43.71) | 177.2 (49.31) | |
| | Median(IQR) | | 172.0 ( 69.0) | 165.0 ( 55.0) | 170.0 ( 60.0) | |
| | Min, Max | | ( 109, 411.0) | ( 110, 290.0) | ( 109, 411.0) | |
| BMI (kg/m^2) | N | | 99 | 99 | 198 | 0.208 |
| | Mean (SD) | | 28.7 ( 8.26) | 27.3 ( 6.37) | 28.0 ( 7.39) | |
| | Median(IQR) | | 27.0 ( 9.2) | 26.2 ( 8.2) | 26.6 ( 8.6) | |
| | Min, Max | | (17.9, 65.7) | (19.1, 51.9) | (17.9, 65.7) | |
| Symptom Days | N (%) | 0-5 | 68 (68.0) | 68 (68.7) | 136 (68.3) | 0.917 |

| | | | | | | |
|---|---|---|---|---|---|---|
| | | 6-10 | 32 (32.0) | 31 (31.3) | 63 (31.7) | |
| WURSS Q1 Screen | N (%) | 4 | 19 (19.0) | 21 (21.2) | 40 (20.1) | 0.963 |
| | | 5 | 51 (51.0) | 48 (48.5) | 99 (49.7) | |
| | | 6 | 23 (23.0) | 24 (24.2) | 47 (23.6) | |
| | | 7 | 7 ( 7.0) | 6 ( 6.1) | 13 ( 6.5) | |
| WURSS Q1 Baseline | N (%) | 4 | 32 (32.0) | 31 (31.3) | 63 (31.7) | 0.935 |
| | | 5 | 51 (51.0) | 48 (48.5) | 99 (49.7) | |
| | | 6 | 15 (15.0) | 17 (17.2) | 32 (16.1) | |
| | | 7 | 2 ( 2.0) | 3 ( 3.0) | 5 ( 2.5) | |
| Residency | N (%) | Canada | 8 ( 8.0) | 5 ( 5.1) | 13 ( 6.5) | 0.401 |
| | | United States | 92 (92.0) | 94 (94.9) | 186 (93.5) | |
| | **Suumary Statistics for Days between Enrollment and Baseline** | | | | | |
| Days Enrollment to Baseline | N | | 100 | 99 | 199 | 0.696 |
| | Mean (SD) | | 1.9 ( 0.92) | 1.8 ( 0.94) | 1.9 ( 0.93) | |
| | Median(IQR) | | 2.0 ( 1.0) | 2.0 ( 1.0) | 2.0 ( 1.0) | |
| | Min, Max | | ( 0.0, 5.0) | ( 0.0, 6.0) | ( 0.0, 6.0) | |
| Days (Categorized) | N (%) | 0 | 2 ( 2.0) | 6 ( 6.1) | 8 ( 4.0) | 0.352 |
| | | 1 | 35 (35.0) | 27 (27.3) | 62 (31.2) | |
| | | 2 | 43 (43.0) | 49 (49.5) | 92 (46.2) | |
| | | 3 | 14 (14.0) | 14 (14.1) | 28 (14.1) | |
| | | 4 | 5 ( 5.0) | 2 ( 2.0) | 7 ( 3.5) | |
| | | 5 | 1 ( 1.0) | 0 | 1 ( 0.5) | |
| | | 6 | 0 | 1 ( 1.0) | 1 ( 0.5) | |

*p<0.05 by two-sample t-test (continuous) and chi-square test (categorical).
