## Supplementary material for "Home-use Photobiomodulation Device Treatment Outcomes for COVID-19": Interim Analysis

### Supplement 5

### Interim Analysis

**Table of Contents**

5.1 Interim Analysis Report

5.2 Interim Efficacy Analysis Methods

Supplement 5.1: Interim Analysis Report

Interim Efficacy Analysis: Sealed Results (page 1 of 2)

| | |
|---|---|
| Device: | Vielight Rx Plus |
| Protocol version, date: | 2.0, 19May20 |
| Protocol title: | A Randomized Study Evaluating the Efficacy of the Vielight Rx Plus in the Treatment of Covid-19 Respiratory Symptoms. |
| SAP date: | 12Sep20 |
| Interim analysis by: | Mark Van Buskirk, Data Reduction LLC, 472 State Route 24, Chester, NJ 07930 |
| Interim analysis date: | 25Jan21 |

Sealed Results

Demography

Table 1 presents summary statistics and frequencies of demographic characteristics of Active and Control subjects. At baseline, 54.8% of patients were randomized to Active treatment and 45.2% to Control treatment. Mean age was 35.4 years (standard deviation (SD) 13.09, minimum 18, maximum 65 years). 64.4% of patients had symptom duration of 0-5 days and 35.6% of patients had symptom duration of 6-10 days. Mean response to question 1 at baseline (mean 4.6, SD 1.08) tended (p=0.063) to be greater in Active (mean 4.8, SD 0.80) than Control (mean 4.3, SD 1.31). Females were represented in 71.2% of patients and represented a statistically significantly difference (p=0.008) in Active (85.0%) than Control (54.5%).

Time to Event

Table 2 provides Kaplan-Meier estimates at the interim based on 73 completed subjects. In a model stratified by symptom duration, the median time to complete resolution of WURSS question 1 symptoms (primary efficacy endpoint) was 15.5 days (95% confidence interval (CI) (10.0, 20.0) in Active and 18.0 days (95% CI 11.0, n/a) in Control, and the p-value by a stratified log-rank test was 0.104. Subgroup analysis by symptom duration categories indicated that the greatest difference in response between treatment groups was in those with symptoms 0-5 days with median time to complete resolution of symptoms 11.0 days for Active and 14.0 days for Control, which was marginally significant (p=0.075). In patients with symptoms 6-10 days, median time to event was 21 days in Active and 22 days in Control (p=0.783).

Table 3 presents results Cox proportional hazards models, stratified by duration of symptoms (0-5 days, 6-10 days). Baseline age, gender and race did not contribute to the regression model, and the primary statistical model for the interim efficacy analysis is stratified by symptom duration with factors for treatment and baseline Q1 covariate. Results indicated that the hazard ratio was 1.72 with 95% CI (0.967, 3.060) and p-value 0.065.

A data listing of demography and derived time-to-event is provided.

Interim Efficacy Analysis: Sealed Results (page 2 of 2)

| | |
|---|---|
| Device: | Vielight Rx Plus |
| Protocol version, date: | 2.0, 19May20 |
| Protocol title: | A Randomized Study Evaluating the Efficacy of the Vielight Rx Plus in the Treatment of Covid-19 Respiratory Symptoms. |
| SAP date: | 12Sep20 |
| Interim analysis by: | Mark Van Buskirk, Data Reduction LLC, 472 State Route 24, Chester, NJ 07930 |
| Interim analysis date: | 25Jan21 |

Interim Efficacy Analysis

At the interim efficacy analysis, the trial decision to stop the trial early for superiority was not met as the p-value for superiority by a symptom-duration stratified Cox model with baseline Q1 score covariate was p=0.065; and did not meet the established O'Brien Fleming[1] threshold for superiority of p<0.0051. The trial decision for futility was not met as the lower bound of the 95% CI for the hazard ratio (0.967) meets the criteria of conditional power >50% from Lan[2] equation 5.

Sample Size Re-Estimation

Sample size re-estimation by Cox regression utilizes the overall event rate (0.74) to update statistical power. Provided the event rate, protocol-stated hazard ratio (1.40) and sample size (280), the statistical power to detect a treatment difference is 68%. Power of 80% is achieved with 375 subjects. Under a fixed sample size condition of n=280 subjects, a hazard ratio of 1.58 is required to achieves 80% power to detect a treatment difference, given an overall event rate of 0.74.

[1] O/Brien PC and Fleming TR (1979). A multiple testing procedure for clinical trials. *Biometrics*. 35:549-556.
[2] Lan KKG and Simon R (1982). Stochastically curtailed tests in long-term clinical trials. *Communications in Statistics* Series C. 1:207-219.

Table 1
Interim Efficacy Analysis
Baseline Characteristics

| Variable | Statistic | Category | Treatment Group: Active (N= 40) | Treatment Group: Control (N= 33) | Treatment Group: Total (N= 73) | P-Value^ |
|---|---|---|---|---|---|---|
| Age (years) | N | | 40 | 33 | 73 | 0.792 |
| | Mean (SD) | | 35.0 (13.77) | 35.8 (12.40) | 35.4 (13.09) | |
| | Median(IQR) | | 33.0 ( 23.5) | 36.0 ( 19.0) | 34.0 ( 23.0) | |
| | Min, Max | | (18.0, 65.0) | (19.0, 63.0) | (18.0, 65.0) | |
| Sex | N (%) | Female | 34 (85.0) | 18 (54.5) | 52 (71.2) | 0.008* |
| | | Male | 6 (15.0) | 15 (45.5) | 21 (28.8) | |
| Caucasian | N (%) | Caucasian | 34 (85.0) | 28 (84.8) | 62 (84.9) | 1.000 |
| | | Non-Caucasian | 6 (15.0) | 5 (15.2) | 11 (15.1) | |
| Sympton Days | N (%) | 0-5 days | 24 (60.0) | 23 (69.7) | 47 (64.4) | 0.465 |
| | | 6-10 days | 16 (40.0) | 10 (30.3) | 26 (35.6) | |
| Question 1 at Screening | N | | 40 | 33 | 73 | 0.512 |
| | Mean (SD) | | 5.0 ( 0.68) | 4.9 ( 0.89) | 4.9 ( 0.78) | |
| | Median(IQR) | | 5.0 ( 0.0) | 5.0 ( 1.0) | 5.0 ( 1.0) | |
| | Min, Max | | ( 4.0, 6.0) | ( 4.0, 7.0) | ( 4.0, 7.0) | |
| Question 1 at Baseline | N | | 40 | 33 | 73 | 0.063 |
| | Mean (SD) | | 4.8 ( 0.80) | 4.3 ( 1.31) | 4.6 ( 1.08) | |
| | Median(IQR) | | 5.0 ( 1.0) | 4.0 ( 1.0) | 5.0 ( 1.0) | |
| | Min, Max | | ( 3.0, 6.0) | ( 1.0, 7.0) | ( 1.0, 7.0) | |
| Randomization to 1st Trtmt Days | N | | 40 | 33 | 73 | 0.803 |
| | Mean (SD) | | 0.5 ( 0.75) | 0.5 ( 0.79) | 0.5 ( 0.77) | |
| | Median(IQR) | | 0.0 ( 1.0) | 0.0 ( 1.0) | 0.0 ( 1.0) | |
| | Min, Max | | ( 0.0, 3.0) | ( 0.0, 2.0) | ( 0.0, 3.0) | |

^ P-value for continuous variables by t-test and 2x2 categorical by Fisher's Exact test.

25JAN21, T01_demog

VieLight Rx Plus.
Protocol: Treatment of Covid-19 Respiratory Symptoms

Table 2
Interim Efficacy Analysis
Overall Time to Complete Resolution of WURSS Question 1 by Kaplan Meier Estimates

| Model | Treatment | N | | | Median Days to Event | | LogRank P-Value | |
|---|---|---|---|---|---|---|---|---|
| | | Total | Failed | Censored | Months | 95% c.i. | Unstratified | Stratified |
| Overall | Active | 40 | 33 | 7 | 15.5 | (10.0,20.0) | 0.175 | 0.104 |
| | Control | 33 | 21 | 12 | 18.0 | (11.0, ) | | |
| Sympton Days 0-5 | Active | 24 | 22 | 2 | 11.0 | ( 8.0,18.0) | 0.075 | |
| | Control | 23 | 15 | 8 | 14.0 | (10.0, ) | | |
| Sympton Days 6-10 | Active | 16 | 11 | 5 | 21.0 | (14.0, ) | 0.783 | |
| | Control | 10 | 6 | 4 | 22.0 | ( 5.0, ) | | |

Overall time (days) to resolution of respiratory symptoms (WURSS Q1) is from treatment start until the event or last follow-up if no event.
Median and 95% confidence interval (c.i.) from Kaplan-Meier estimation in model stratified by symptom-days (0-6,6-10).25JAN21, T02_km_interim

Table 3
Interim Efficacy Analysis
Overall Time to Complete Resolution of WURSS Question 1 by Cox Proportional Hazards Model

| Model | Parameter | Hazard Ratio | ---95% Confidence Interval---- Lower | Upper | P-Value |
|---|---|---|---|---|---|
| 1) Treatment Model | Treatment | 1.5817 | 0.9055 | 2.7631 | 0.107 |
| 2) Treatment and Q1 Baseline Covariate | Treatment | 1.7202 | 0.9670 | 3.0601 | 0.065 |
| | Q1 Baseline | 0.8084 | 0.6091 | 1.0729 | 0.141 |

Overall time (days) to resolution of respiratory symptoms (WURSS Q1) is from treatment start until the event or last follow-up if no event
P-value by symptom duration stratified Cox proportional hazards model. Model 2 adjusted for baseline Q1. 25JAN21, T03_cox_interim

#### Supplement 5.2

#### Interim Efficacy Analysis Methods

Interim Efficacy Analysis

| | |
|---|---|
| Device: | Vielight Rx Plus |
| Protocol version, date: | 2.0, 19May20 |
| Protocol title: | A Randomized Study Evaluating the Efficacy of the Vielight Rx Plus in the Treatment of Covid-19 Respiratory Symptoms. |
| SAP date: | 12Sep20 |
| Interim analysis by: | Mark Van Buskirk, Data Reduction LLC, 472 State Route 24, Chester, NJ 07930 |
| Interim analysis date: | 26Sep20 |

Statistical Methods

The primary efficacy endpoint is defined as the time in days from date of first use of the device to the start date of improvement, defined as three consecutive days of reporting 0 on WURSS-44 question 1. The time to endpoint (days) is met (success) on the first of three consecutive days.

A single interim efficacy analysis was conducted on the primary efficacy endpoint in the first 73 subjects. The stopping boundary for superiority was defined as $p<0.0051$ by O'Brien and Fleming[1] group-sequential testing design. The stopping boundary for futility is set at conditional power[2]

Results from the interim efficacy analysis are reported as 'unsealed' and 'sealed' results.

Unsealed results inform:

(1) Stop/Continue study decision:
Results communicated to the sponsor include the decision to either (a) stop the study for superiority, (b) stop the study for inferiority, or (c) continue the study until planned completion.

(2) Sample size re-estimation:
The statistical power of the study is addressed at the interim efficacy analysis. Statistical power analysis by Cox Proportional Hazards[3] is performed using PASS software (NCSS Statistical Software, version 11.0). Sample size re-estimation uses the protocol specified hazard ratio of

---

[1] O'Brien PC and Fleming TR (1979). A multiple testing procedure for clinical trials. Biometrics. 35: 549-556.

[2] Lan KKG and Simon R (1982). Stochastically curtailed tests in long-term clinical trials. Communications in Statistics Series C. 1: 207-219.

[3] Schoenfeld, David A. 1983. 'Sample-size formula for the proportional-hazards regression model', Biometrics, Volume 39, pages 499-503.

1.40 and sample size of 280 subjects and the observed event rate of 74% at the interim efficacy analysis.

Sealed results consist of treatment group comparisons of baseline characteristics, Kaplan-Meier time-to-event estimates, and hazard ratios and confidence intervals by Cox proportional hazards models. Statistical analysis of the primary efficacy endpoint is by Kaplan-Meier estimation, stratified by time since onset of symptoms (<=5 days, 6-10 days). Median days to improvement and 95% confidence



intervals are reported by treatment group. Separately by treatment group, the total number of subjects with the event (success of primary endpoint), without the event (censored), and total number of subjects are presented. For Kaplan-Meier estimation, the stratified test and univariate model by time of onset of symptoms are presented. The Cox proportional hazards model stratified by time since onset of symptoms was applied, and the hazard ratio and 95% confidence interval reported. Additionally, a stratified Cox model including covariate of baseline response to question 1 is reported.
