## Supplementary material for "Home-use Photobiomodulation Device Treatment Outcomes for COVID-19": Kaplan-Meier Method Results

### Supplement 6

### Kaplan-Meier Method

**Table of Contents**

6.1 Introduction to the Kaplan-Meier Method Used

6.2 Outcomes Affecting >5% of Patients for Time-to-recovery and Hazard Ratio (P<0.050)

6.3 Full Table for Kaplan-Meier Estimates in Patients with a WURSS-44 Q1 Score of 4-7 at Baseline

6.4 Kaplan-Meier Curves of Primary Outcome

6.5 Kaplan-Meier Curves for Secondary Symptoms with Duration 0-5 days and 6-10 days at Screening (P≤0.050)

#### Supplement 6.1

##### Introduction to the Kaplan-Meier Method Used

The Kaplan-Meier (KM) Method is utilized to estimate the time-to-recovery (days) applied to the 43 items/symptoms listed in WURSS-44 for primary and secondary efficacy outcomes. “Recovery” is defined as having 3 consecutive days of no symptom, or a score of ‘0’ on the WURSS-44 scale. The first day of the 3-day recovery is used to compute days-to-recovery.

The time-to-recovery values in the primary endpoint were assessed with the KM method, and the difference between the Treatment and Control groups were assessed with the log-rank test, stratified by symptom duration days. In calculating the time-to-recovery for the secondary symptoms, the KM Method and log-rank tests were applied in the same way as for the primary outcome.

‘Day 1’ is the Baseline, defined as the day of first-use of the treatment device. Knowing that there is a roughly 2 days delay in receiving treatment. Symptom duration days can be interpreted as having added a median of 2 days to the 0-5 days strata duration, the equivalent of 0-7 days. Similarly, the 6-10 day symptom duration stratum is equivalent to 8-12 days at baseline.

Patients who did not achieve recovery at the end of the 30-day assessment period were censored. This also included patients whose 3-day results were incomplete.

In this Supplement section, we include a table of significant outcomes for WURSS-44 secondary endpoints (Supplement 6.2, also presented as Table 3), the full table for all the WURSS-44 items (Supplement 6.3) as well as the KM curves for the Primary (Supplement 6.4) and Secondary outcomes (Supplement 6.5) with significance ($P \leq 0.05$).

**Supplement 6.2**

**Outcomes Affecting >5% of Patients for Time-to-recovery and Hazard Ratio (P<0.050)**

| Symptom | Days with Symptoms at Screening | Treatment | Kaplan-Meier N Total | N Recovered | N Censored | Median Days to Recovery | 95% CI | KM Log-rank P-value | Cox Proportional Hazards Hazard Ratio | 95% CI | Cox P-value |
|---|---|---|---|---|---|---|---|---|---|---|---|
| **Patients with 0-10 Days Symptom Duration (all at Baseline)** | | | | | | | | | | | |
| Sinus pain | 0 to 10 | Treatment | 39 | 35 | 4 | 10 | (8, 13) | 0.005 | 2.001 | (1.203, 3.328) | 0.008 |
| | | Control | 42 | 30 | 12 | 17 | (9, 25) | | | | |
| | | Total | 81 | 65 | 16 | 7 | | | | | |
| Chest congestion | 0 to 10 | Treatment | 43 | 38 | 5 | 15 | (9, 18) | 0.017 | 1.878 | (1.105, 3.193) | 0.02 |
| | | Control | 35 | 22 | 13 | 21 | (17, 30) | | | | |
| | | Total | 78 | 60 | 18 | 6 | | | | | |
| Body aches | 0 to 10 | Treatment | 60 | 52 | 8 | 12 | (9,15) | 0.019 | 1.652 | (1.084, 2.519) | 0.02 |
| | | Control | 59 | 39 | 20 | 15 | (12, 20) | | | | |
| | | Total | 119 | 119 | 28 | 3 | | | | | |
| Think clearly | 0 to 10 | Treatment | 44 | 34 | 10 | 11 | (9, 15) | 0.020 | 1.893 | (1.013, 3.248) | 0.021 |
| | | Control | 41 | 23 | 18 | 21 | (12, 30) | | | | |
| | | Total | 85 | 57 | 28 | 10 | | | | | |
| Ear discomfort | 0 to 10 | Treatment | 24 | 21 | 3 | 12.5 | (8, 17) | 0.023 | 2.325 | (1.104, 4.893) | 0.026 |
| | | Control | 21 | 11 | 10 | 24.0 | (12, 36) | | | | |
| | | Total | 45 | 32 | 13 | 13.5 | | | | | |
| Sinus drainage | 0 to 10 | Treatment | 28 | 22 | 6 | 12 | (10, 17) | 0.026 | 2.14 | (1.088, 4.203) | 0.028 |
| | | Control | 27 | 15 | 12 | 23 | (10, 36) | | | | |
| | | Total | 55 | 37 | 18 | 11 | | | | | |
| Headache | 0 to 10 | Treatment | 67 | 53 | 14 | 14 | (11, 20) | 0.031 | 1.586 | (1.036, 2.429) | 0.034 |
| | | Control | 61 | 36 | 25 | 21 | (14, 28) | | | | |
| | | Total | 128 | 89 | 39 | 7 | | | | | |
| Coughing up stuff | 0 to 10 | Treatment | 27 | 25 | 2 | 13 | (10, 20) | 0.037 | 1.817 | (1.013, 3.261) | 0.045 |
| | | Control | 32 | 21 | 11 | 21 | (14, 27) | | | | |
| | | Total | 59 | 46 | 13 | 8 | | | | | |
| Sneezing | 0 to 10 | Treatment | 33 | 29 | 4 | 13 | (9, 18) | 0.049 | 1.92 | (1.064, 3.467) | 0.030 |
| | | Control | 22 | 14 | 8 | 17 | (10, 24) | | | | |
| | | Total | 55 | 43 | 12 | 4 | | | | | |
| **Patients with 0-5 Days Symptom Duration** | | | | | | | | | | | |
| Headache | 0 to 5 | Treatment | 50 | 43 | 7 | 13 | (10, 16) | 0.006 | 2.027 | (1.216, 3.378) | 0.007 |
| | | Control | 40 | 23 | 17 | 19 | (14, 24) | | | | |
| | | Total | 90 | 66 | 24 | 6 | | | | | |
| Sinus pain | 0 to 5 | Treatment | 28 | 24 | 4 | 9 | (7, 13) | 0.022 | 1.926 | (1.074, 3.452) | 0.028 |
| | | Control | 32 | 22 | 10 | 15 | (9, 25) | | | | |
| | | Total | 60 | 46 | 14 | 6 | | | | | |
| Think clearly | 0 to 5 | Treatment | 29 | 22 | 7 | 10 | (9, 15) | 0.024 | 2.067 | (1.093, 3.909) | 0.025 |
| | | Control | 30 | 17 | 13 | 21 | (12, 30) | | | | |
| | | Total | 59 | 39 | 20 | 11 | | | | | |
| Swollen glands | 0 to 5 | Treatment | 20 | 19 | 1 | 8 | (6, 10) | 0.029 | 2.436 | (1.046, 5.676 | 0.039 |
| | | Control | 13 | 9 | 4 | 10 | (7, 13) | | | | |
| | | Total | 33 | 28 | 5 | 2 | | | | | |
| Chest congestion | 0 to 5 | Treatment | 32 | 28 | 4 | 15 | (9, 20) | 0.042 | 1.847 | (1.008, 3.384) | 0.047 |
| | | Control | 27 | 17 | 10 | 21 | (10. 32) | | | | |
| | | Total | 59 | 45 | 14 | 6 | | | | | |
| Body aches | 0 to 5 | Treatment | 46 | 39 | 7 | 12 | (9, 17) | 0.050 | 1.641 | (0.996, 2.704) | 0.052 |
| | | Control | 39 | 26 | 13 | 15 | (12, 23) | | | | |
| | | Total | 85 | 65 | 20 | 3 | | | | | |
| **Patients with 6-10 Days Symptom Duration** | | | | | | | | | | | |
| Feeling tired | 6 to 10 | Treatment | 25 | 9 | 16 | 25.5 | (24, 27 ) | 0.037 | 0.432 | (0.192, 0.972) | 0.094 |
| | | Control | 29 | 17 | 12 | 20 | (13, 27 ) | | | | |
| | | Total | 54 | 26 | 28 | 4.5 | | | | | |

| | | | | | | | | | | | |
|---|---|---|---|---|---|---|---|---|---|---|---|
| Lack of energy | 6 to 10 | Treatment | 25 | 10 | 15 | 26.5 | (27, 26) | 0.049 | 0.47 | (0.220, 0.996) | 0.049 |
| | | Control | 27 | 16 | 11 | 20 | (15, 25) | | | | |
| | | Total | 52 | 26 | 26 | 3.5 | | | | | |

### Supplement 6.3

**Full Table for Kaplan-Meier Estimates**
**in Patients with a WURSS-44 Q1 Score of 4-7 at Baseline**

| | | | N | | | Median | | | |
|---|---|---|---|---|---|---|---|---|---|
| WURSS Questionnaire Item | Symptom Days | Treatment | Total | Recovered | Censored | Days | | 95% CI | P-value^ |
| 01: How sick do you feel today? | 0-5d | Treatment | 68 | 53 | 15 | 18 | | (13, 20) | 0.050 |
| | | Control | 68 | 42 | 26 | 21 | | (15, 28) | |
| | | Total | 136 | 95 | 41 | | | | |
| | 6-10d | Treatment | 32 | 19 | 13 | 23 | | (19, 27)^ | 0.507 |
| | | Control | 31 | 19 | 12 | 21 | | (15, 27)^ | |
| | | Total | 63 | 38 | 25 | | | | |
| | 0-10d | Treatment | 100 | 72 | 28 | 19 | | (16, 22) | 0.197 |
| | | Control | 99 | 61 | 38 | 21 | | (19, 25) | |
| | | Total | 199 | 133 | 66 | | | | |
| 02: Cough | 0-5d | Treatment | 44 | 33 | 11 | 21 | | (16, 27) | 0.572 |
| | | Control | 38 | 25 | 13 | 23 | | (18, 28) | |
| | | Total | 82 | 58 | 24 | | | | |
| | 6-10d | Treatment | 18 | 11 | 7 | 25 | | (21, 29)^ | 0.429 |
| | | Control | 18 | 12 | 6 | 18 | | (12, 24)^ | |
| | | Total | 36 | 23 | 13 | | | | |
| | 0-10d | Treatment | 62 | 44 | 18 | 23 | | (20, 27) | 0.953 |
| | | Control | 56 | 37 | 19 | 22 | | (17, 27) | |
| | | Total | 118 | 81 | 37 | | | | |
| 03: Coughing stuff up | 0-5d | Treatment | 17 | 15 | 2 | 13 | | (11, 28) | 0.100 |
| | | Control | 23 | 14 | 9 | 21 | | (13, 29)^ | |
| | | Total | 40 | 29 | 11 | | | | |
| | 6-10d | Treatment | 10 | 10 | 0 | 11 | | ( 6, 20) | 0.197 |
| | | Control | 9 | 7 | 2 | 17 | | ( 8, 26)^ | |
| | | Total | 19 | 17 | 2 | | | | |
| | 0-10d | Treatment | 27 | 25 | 2 | 13 | | (10, 20) | 0.037* |
| | | Control | 32 | 21 | 11 | 21 | | (14, 27) | |
| | | Total | 59 | 46 | 13 | | | | |

* p<0.05 by log-rank test for symptom days 0-5 and 6-10, and symptom-day stratified log-rank test over symptom days. 95% confidence interval (CI).

Success (recovery) criteria defined as first day of 3 consecutive days of a zero score (does not have this symptom, not at all).

Note, when median not reached, the mean (biased) is used (^). When upper confidence interval (CI) not available, 2*mean-CI_lower (^) is used.
17MAR22,T02_km

#### Supplement 6.4

#### Kaplan-Meier Curve of Primary Outcome

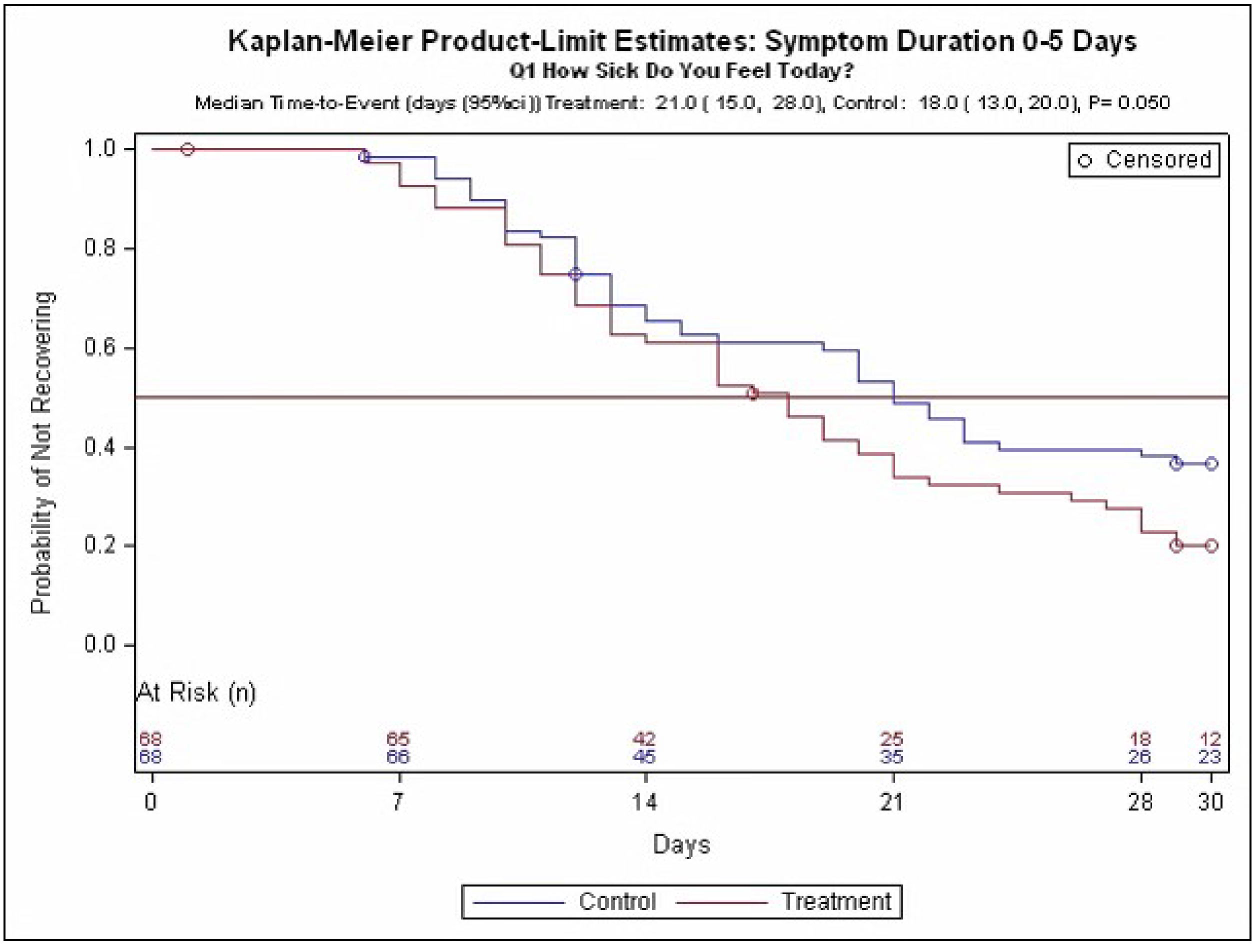


*This figure shows the "survival" probability of the primary outcomes of WURSS-44 Q1: "How sick do you feel today?" for the stratified group with symptom duration of 0-5 days at screening. "Survival" is interpreted as the endpoint not being met yet (did not recover). Subjects that did not have full recovery were censored. The difference in the median time-to-recovery (95% CI) between the two groups were estimated with the log-rank test, and its significance presented in the P-value.*

**Supplement 6.5**

**Kaplan-Meier Curves for Secondary Symptoms with Duration 0-5 days at Screening (P≤0.050)**

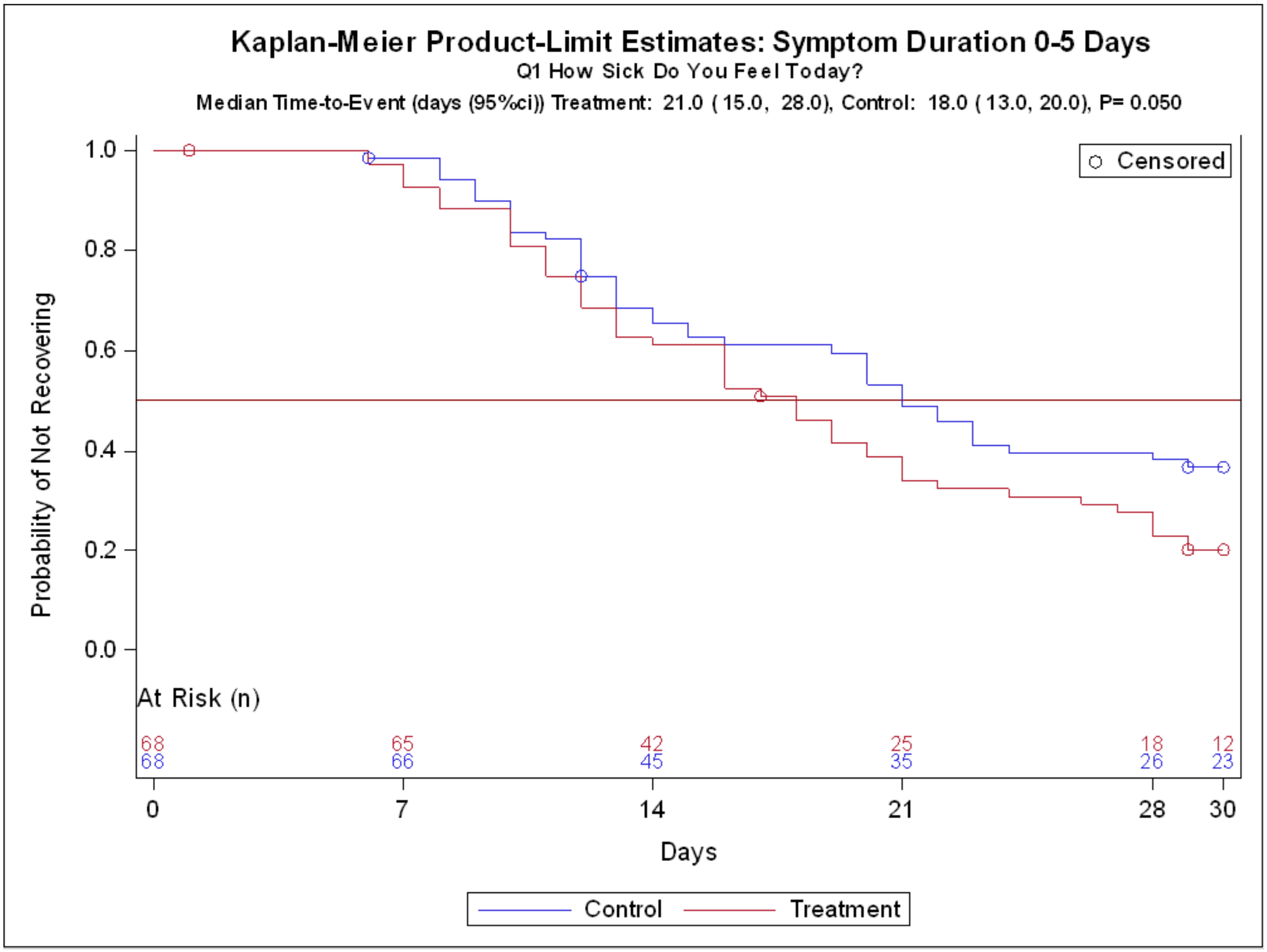

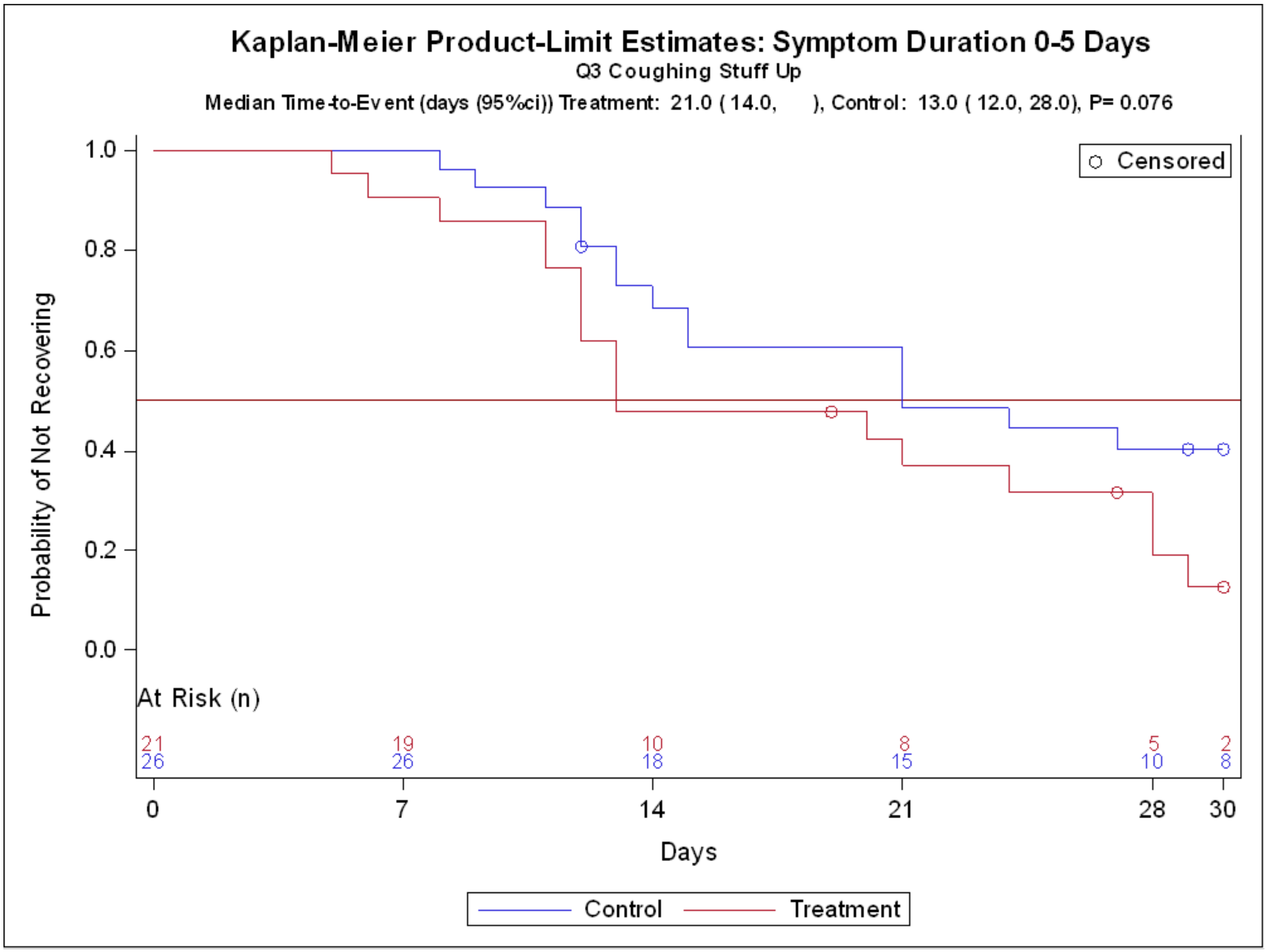

Kaplan-Meier Product-Limit Estimates: Symptom Duration 0-5 Days
Q3 Coughing Stuff Up
Median Time-to-Event (days (95%ci)) Treatment: 21.0 ( 14.0, ), Control: 13.0 ( 12.0, 28.0), P= 0.076
Censored
Probability of Not Recovering
1.0
0.8
0.6
0.4
0.2
0.0
At Risk (n)
21 19 10 8 5 2
26 26 18 15 10 8
0 7 14 21 28 30
Days
Control
Treatment

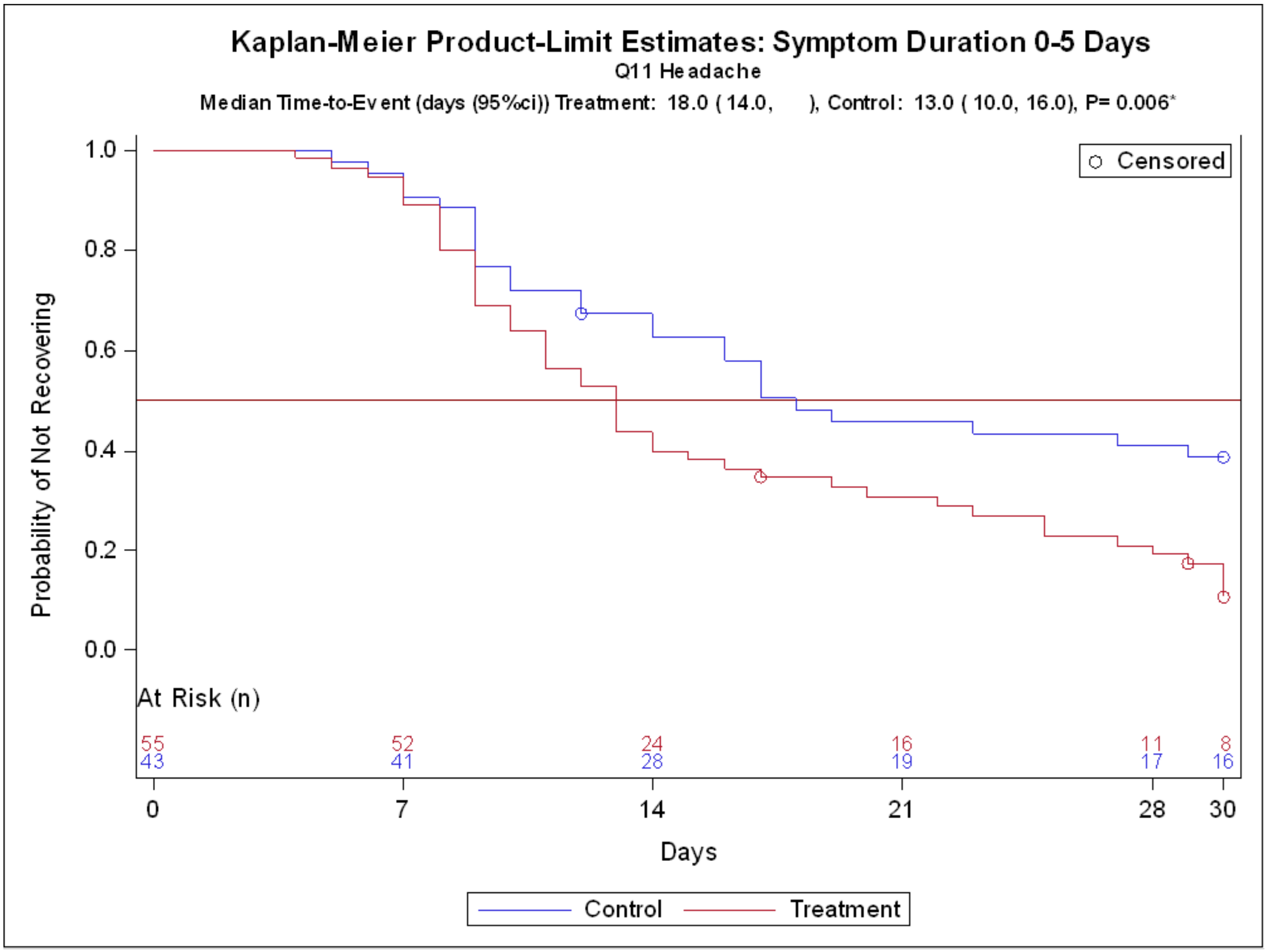

Kaplan-Meier Product-Limit Estimates: Symptom Duration 0-5 Days
Q11 Headache
Median Time-to-Event (days (95%ci)) Treatment: 18.0 ( 14.0, ), Control: 13.0 ( 10.0, 16.0), P= 0.006*
Censored
Probability of Not Recovering
1.0
0.8
0.6
0.4
0.2
0.0
At Risk (n)
55
43
52
41
24
28
16
19
11
17
8
16
0
7
14
21
28
30
Days
Control
Treatment

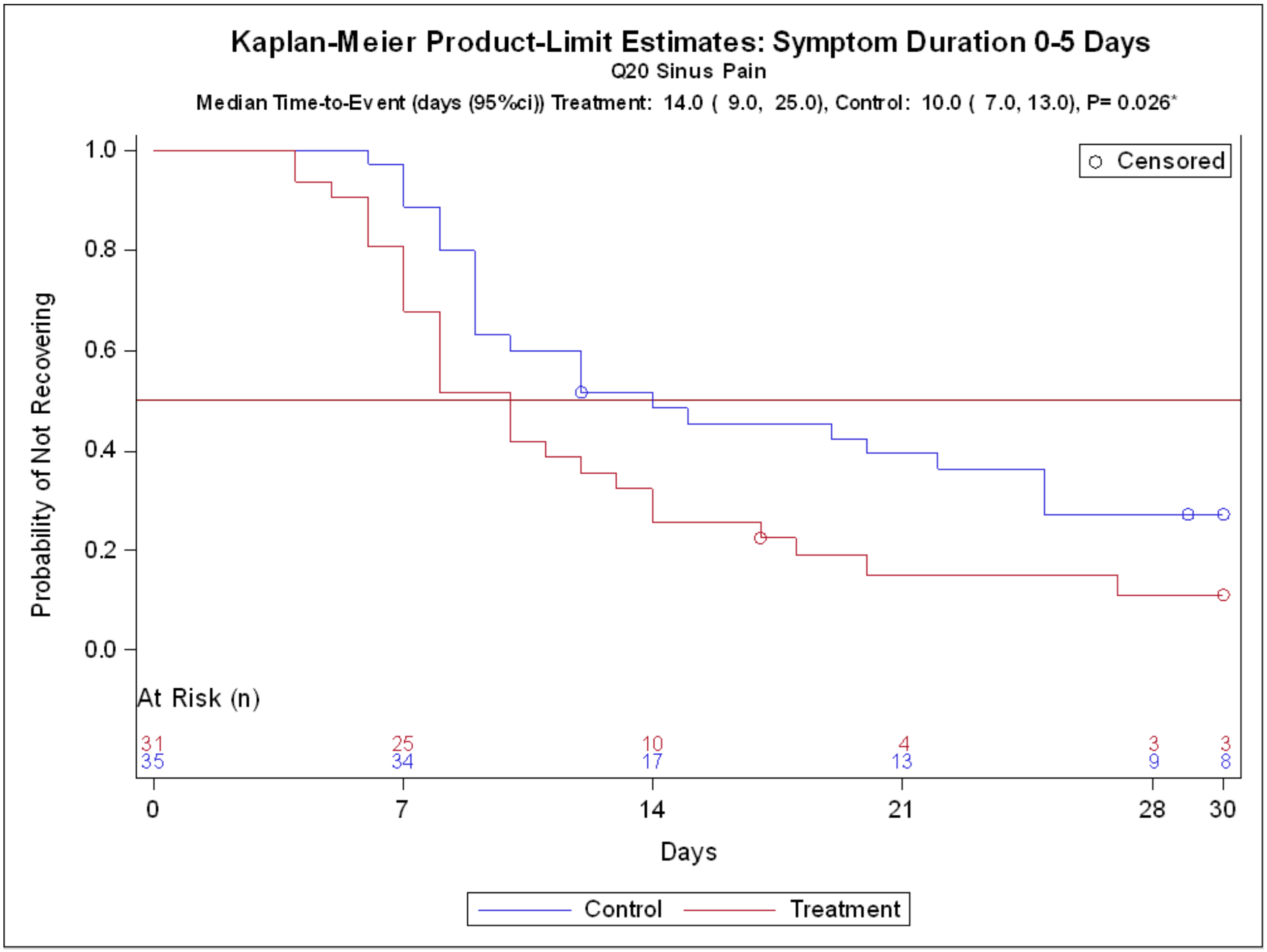

Kaplan-Meier Product-Limit Estimates: Symptom Duration 0-5 Days
Q20 Sinus Pain
Median Time-to-Event (days (95%ci)) Treatment: 14.0 ( 9.0, 25.0), Control: 10.0 ( 7.0, 13.0), P= 0.026*
Probability of Not Recovering
1.0
0.8
0.6
0.4
0.2
0.0
Censored
At Risk (n)
31
25
10
4
3
3
35
34
17
13
9
8
0
7
14
21
28
30
Days
Control
Treatment

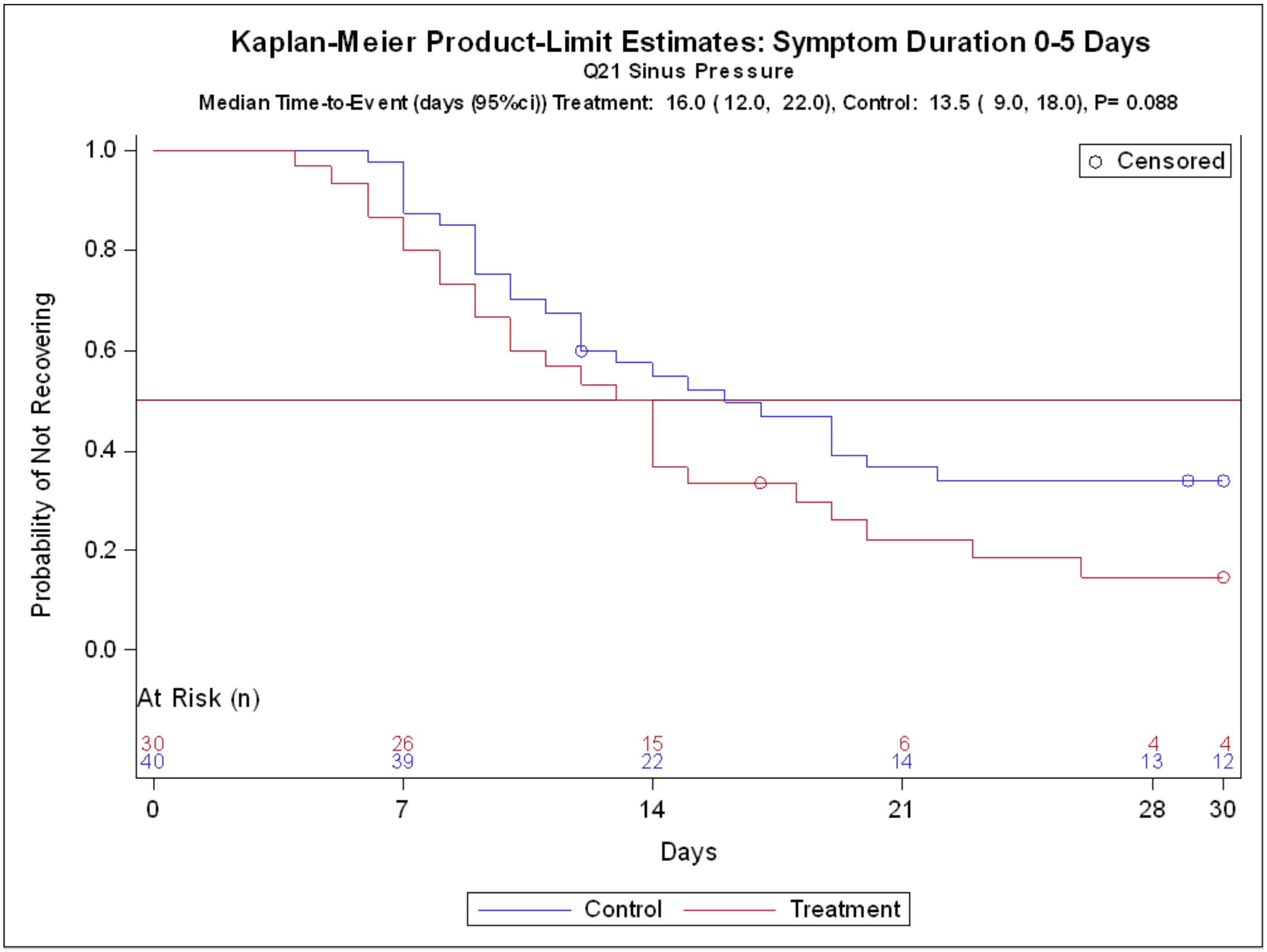
Kaplan-Meier Product-Limit Estimates: Symptom Duration 0-5 Days
Q21 Sinus Pressure
Median Time-to-Event (days (95%ci)) Treatment: 16.0 ( 12.0, 22.0), Control: 13.5 ( 9.0, 18.0), P= 0.088
Probability of Not Recovering
1.0
0.8
0.6
0.4
0.2
0.0
Censored
At Risk (n)
30 26 15 6 4 4
40 39 22 14 13 12
0
7
14
21
28
30
Days
Control
Treatment

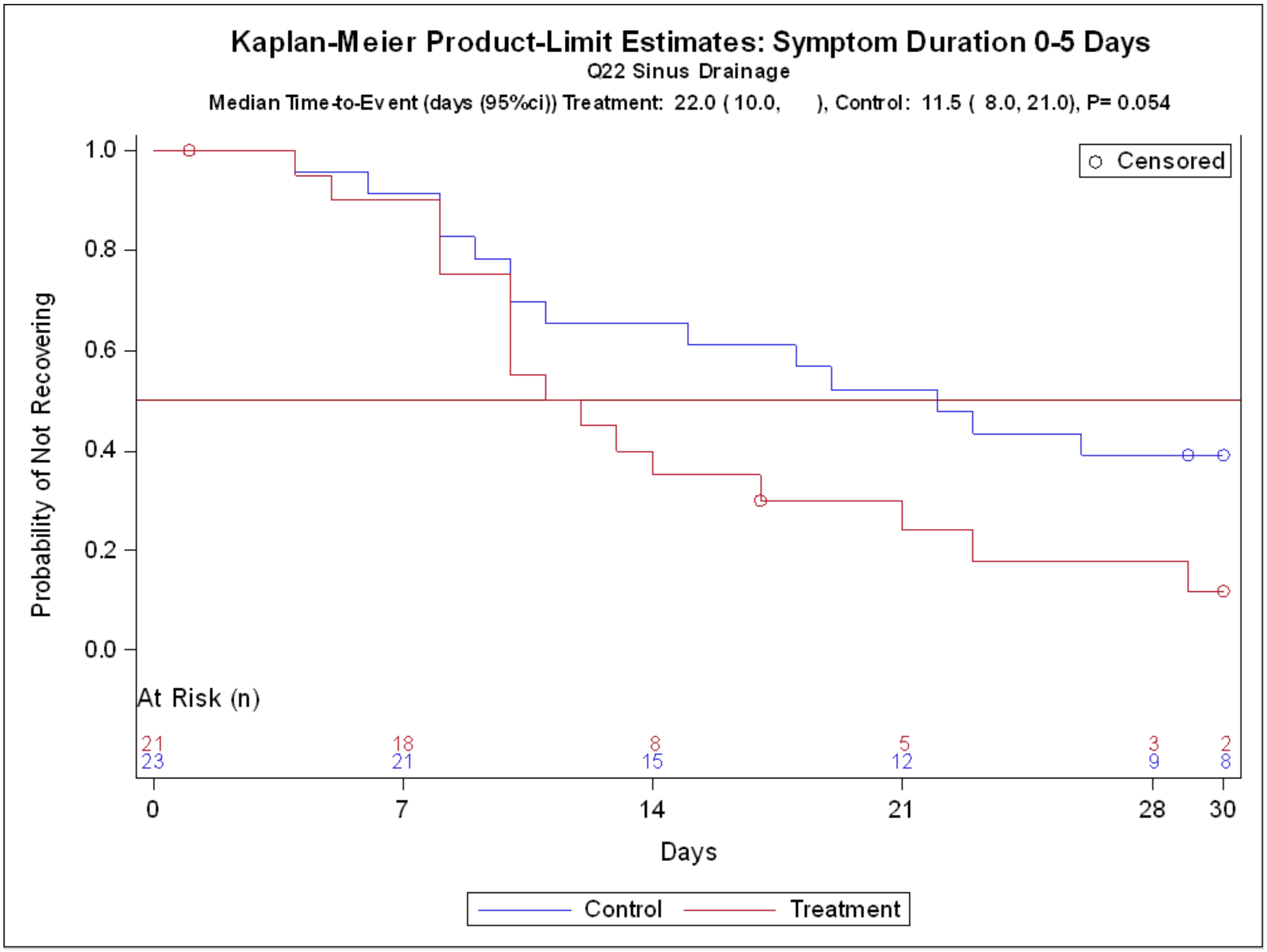

Kaplan-Meier Product-Limit Estimates: Symptom Duration 0-5 Days
Q22 Sinus Drainage
Median Time-to-Event (days (95%ci)) Treatment: 22.0 ( 10.0, ), Control: 11.5 ( 8.0, 21.0), P= 0.054
Censored
Probability of Not Recovering
1.0
0.8
0.6
0.4
0.2
0.0
At Risk (n)
21 18 8 5 3 2
23 21 15 12 9 8
0
7
14
21
28
30
Days
Control
Treatment

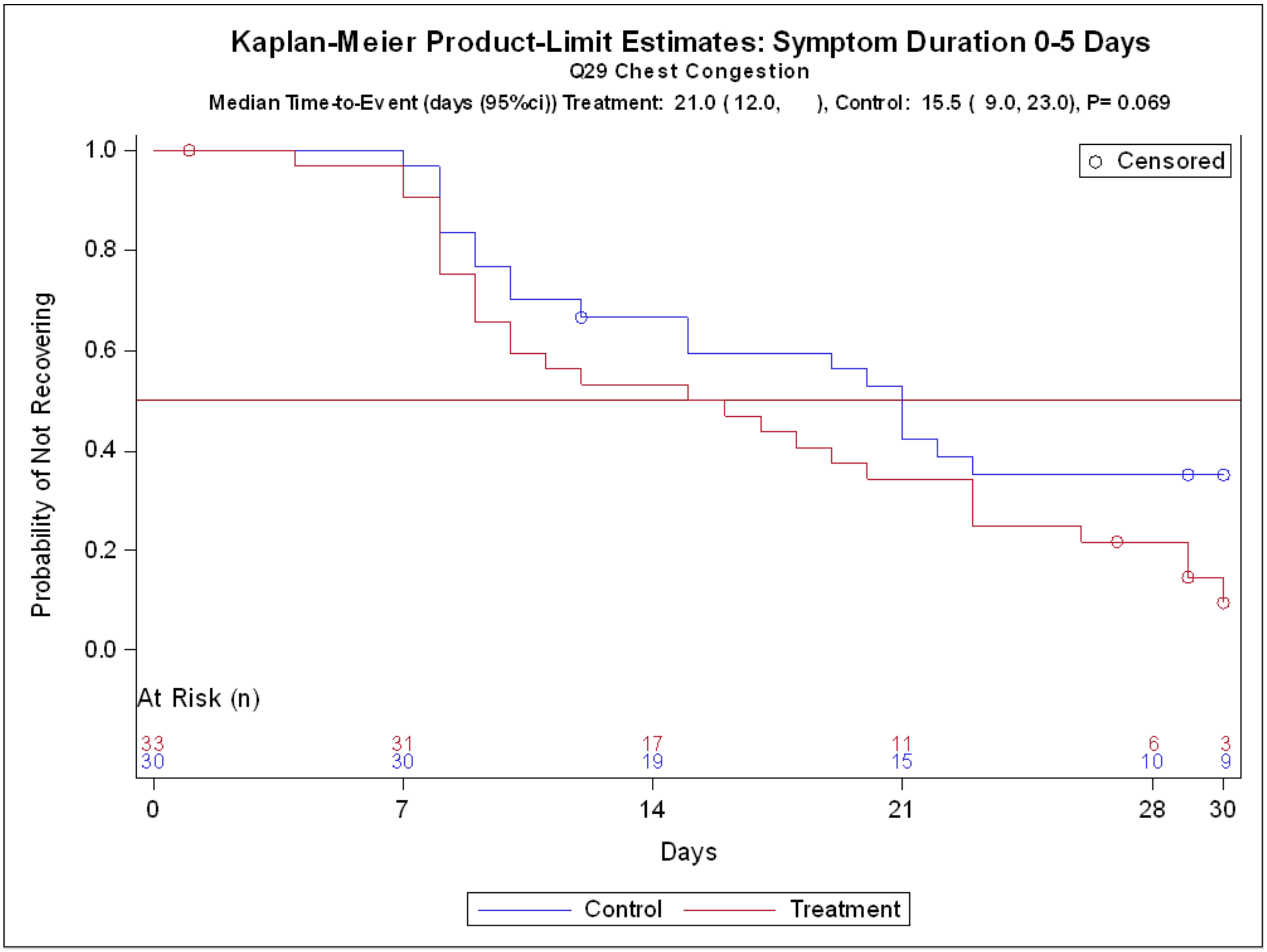
Kaplan-Meier Product-Limit Estimates: Symptom Duration 0-5 Days
Q29 Chest Congestion
Median Time-to-Event (days (95%ci)) Treatment: 21.0 ( 12.0, ), Control: 15.5 ( 9.0, 23.0), P= 0.069
Probability of Not Recovering
1.0
0.8
0.6
0.4
0.2
0.0
Censored
At Risk (n)
33
30
31
30
17
19
11
15
6
10
3
9
0
7
14
21
28
30
Days
Control
Treatment

#### Kaplan-Meier Curves for Secondary Symptom Duration 6-10 days at Screening with P≤0.05

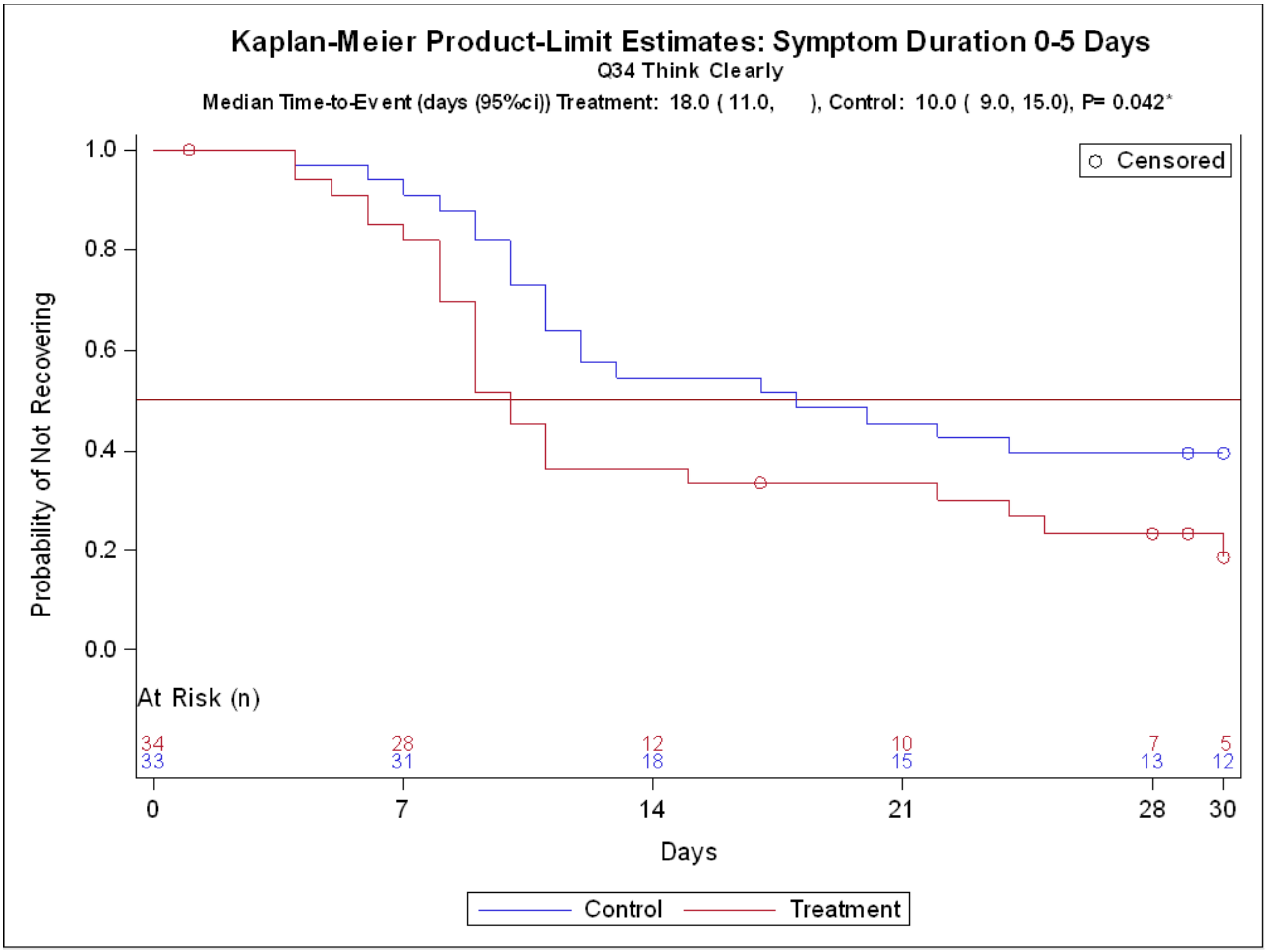

**Supplement 6.5**

**Vielight RX Plus for Covid-19**

**Symptom Duration 6-10 days at Screening (P≤0.050)**

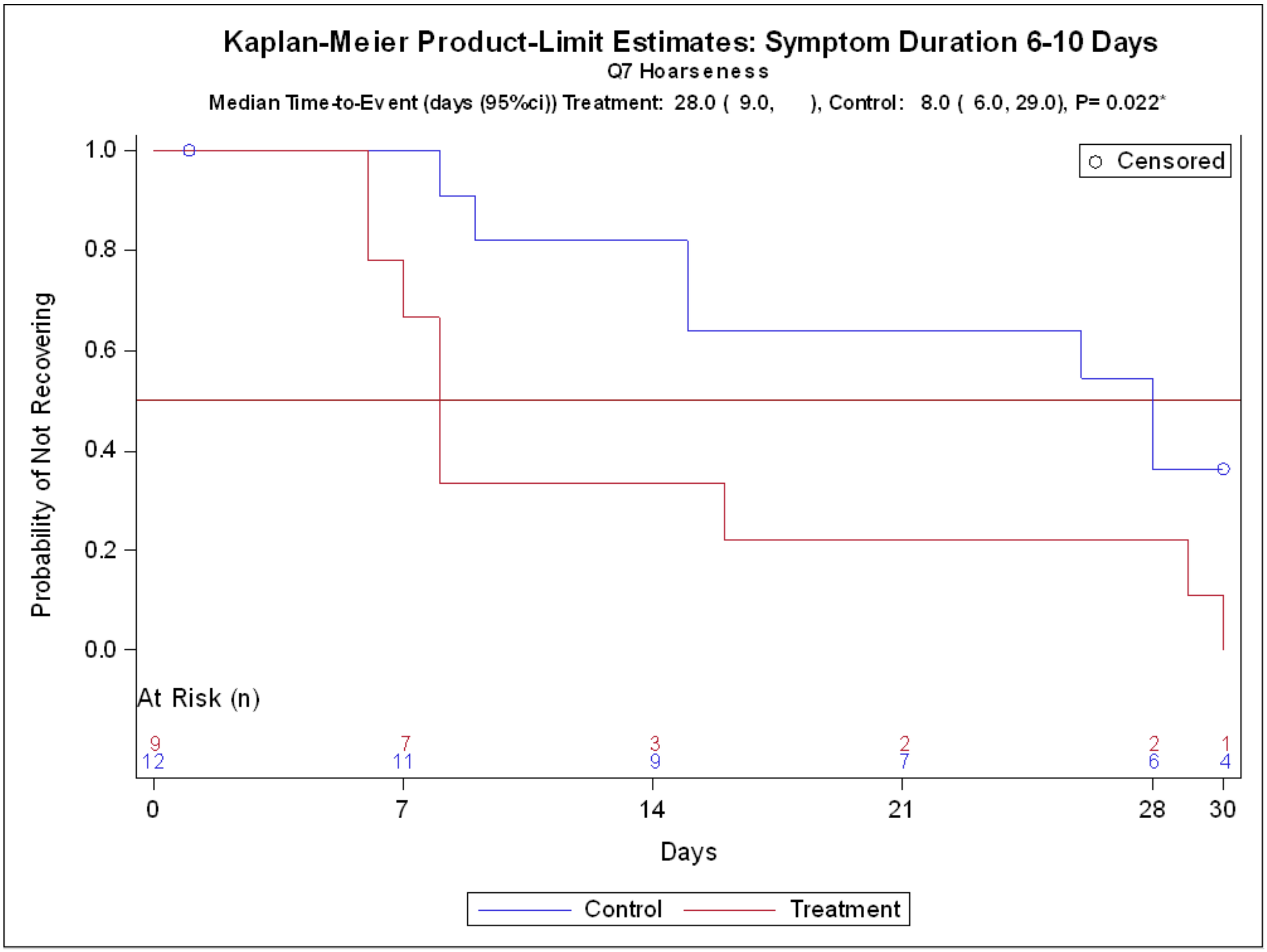

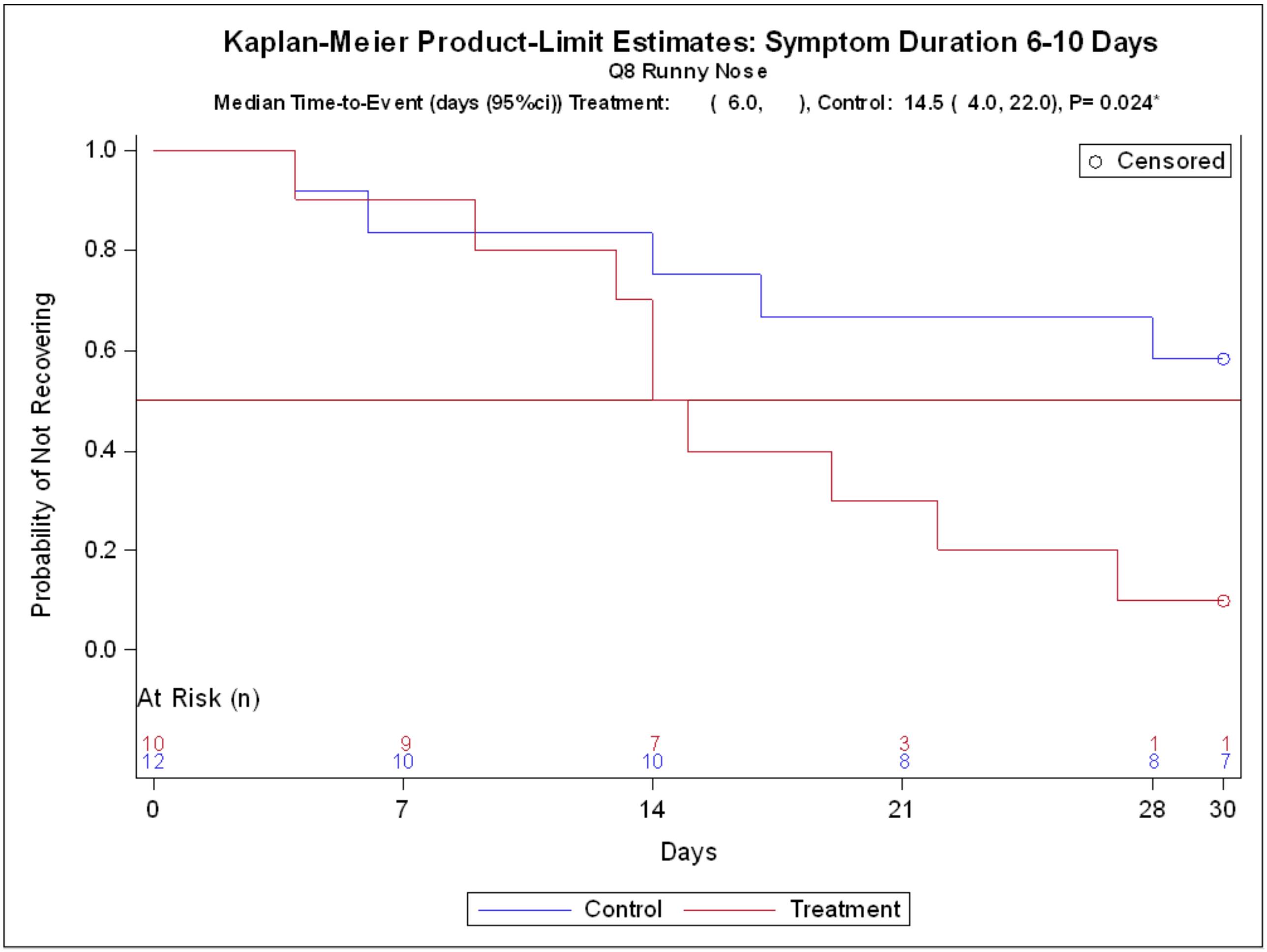

Kaplan-Meier Product-Limit Estimates: Symptom Duration 6-10 Days
Q8 Runny Nose
Median Time-to-Event (days (95%ci)) Treatment: ( 6.0, ), Control: 14.5 ( 4.0, 22.0), P= 0.024*
Censored
Probability of Not Recovering
1.0
0.8
0.6
0.4
0.2
0.0
At Risk (n)
10 9 7 3 1 1
12 10 10 8 8 7
0 7 14 21 28 30
Days
Control
Treatment

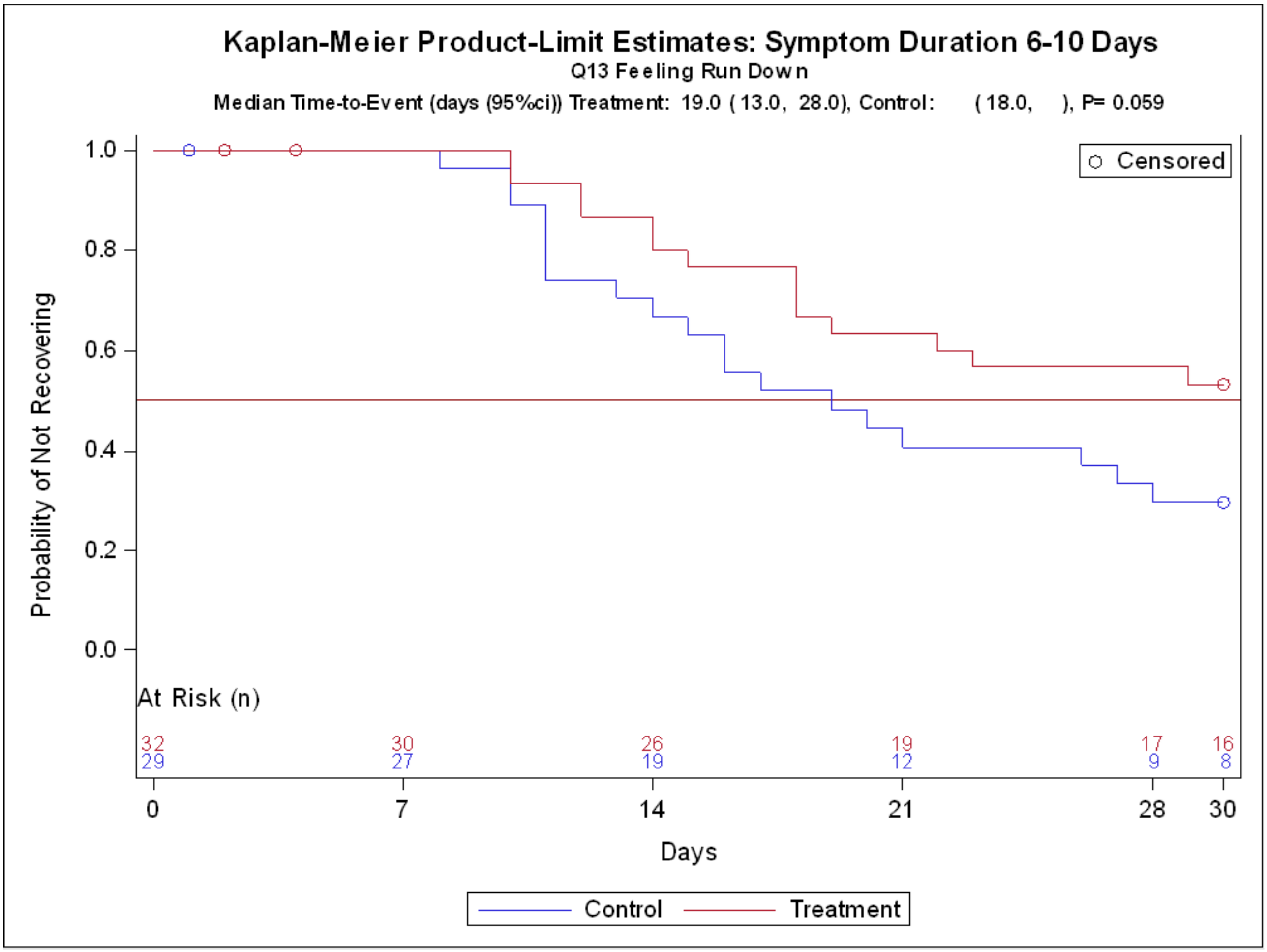

Kaplan-Meier Product-Limit Estimates: Symptom Duration 6-10 Days
Q13 Feeling Run Down
Median Time-to-Event (days (95%ci)) Treatment: 19.0 ( 13.0, 28.0), Control: ( 18.0, ), P= 0.059
Censored
Probability of Not Recovering
1.0
0.8
0.6
0.4
0.2
0.0
At Risk (n)
32 30 26 19 17 16
29 27 19 12 9 8
0 7 14 21 28 30
Days
Control
Treatment

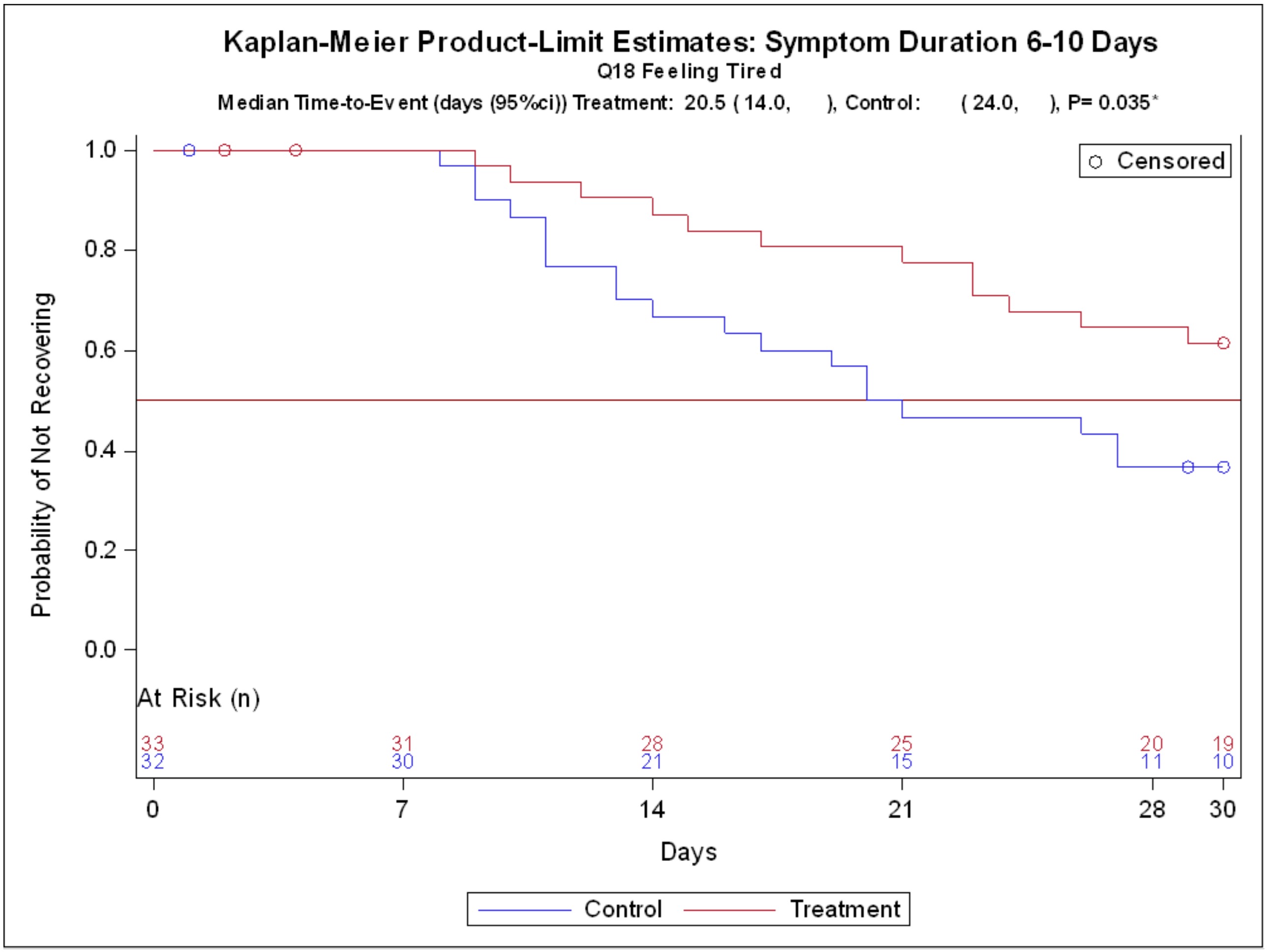
Kaplan-Meier Product-Limit Estimates: Symptom Duration 6-10 Days
Q18 Feeling Tired
Median Time-to-Event (days (95%ci)) Treatment: 20.5 ( 14.0, ), Control: ( 24.0, ), P= 0.035*
Censored
Probability of Not Recovering
1.0
0.8
0.6
0.4
0.2
0.0
At Risk (n)
33 31 28 25 20 19
32 30 21 15 11 10
0 7 14 21 28 30
Days
Control
Treatment

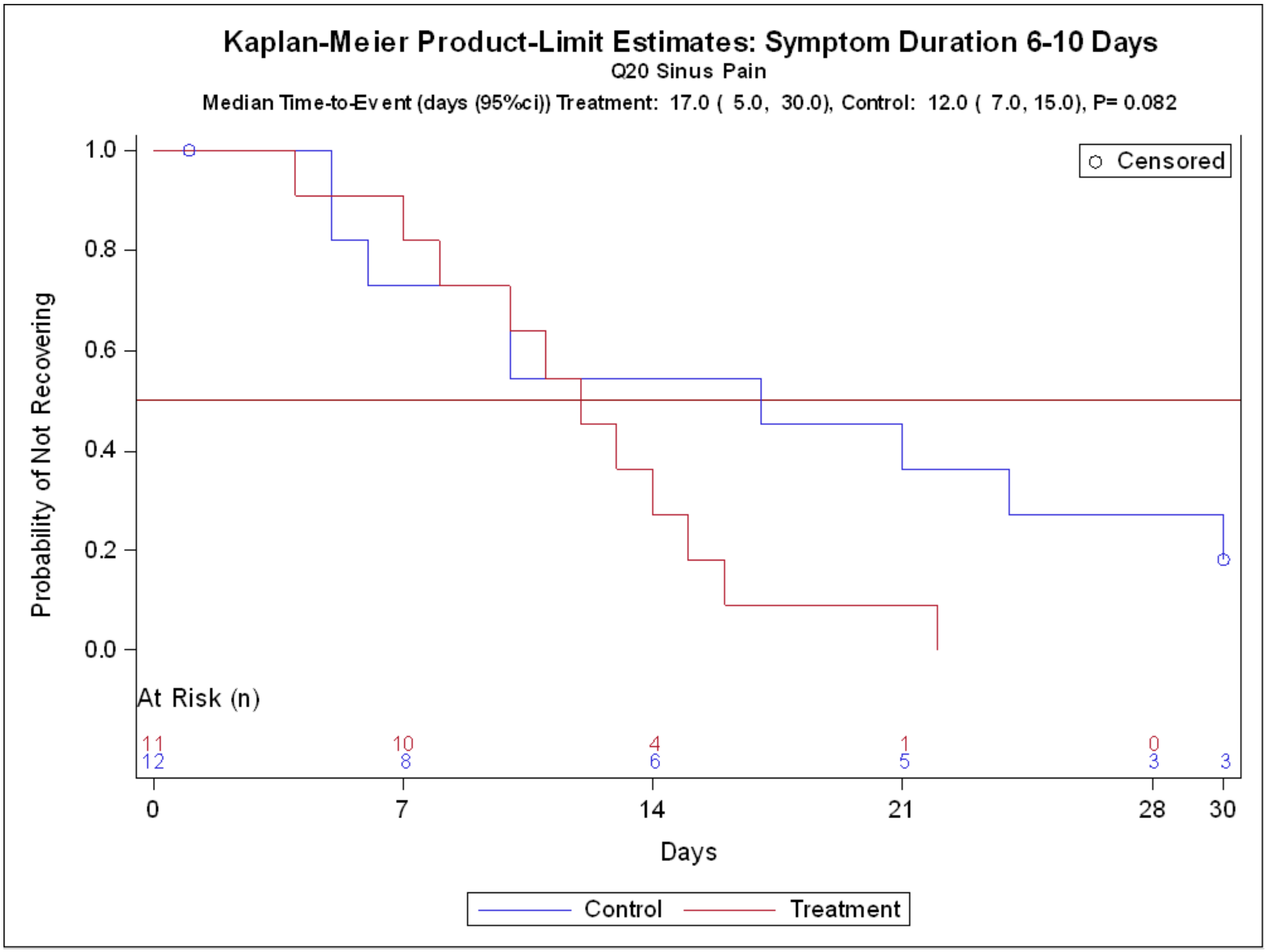
Kaplan-Meier Product-Limit Estimates: Symptom Duration 6-10 Days
Q20 Sinus Pain
Median Time-to-Event (days (95%ci)) Treatment: 17.0 ( 5.0, 30.0), Control: 12.0 ( 7.0, 15.0), P= 0.082
Censored
Probability of Not Recovering
1.0
0.8
0.6
0.4
0.2
0.0
At Risk (n)
11
10
4
1
0
12
8
6
5
3
3
0
7
14
21
28
30
Days
Control
Treatment

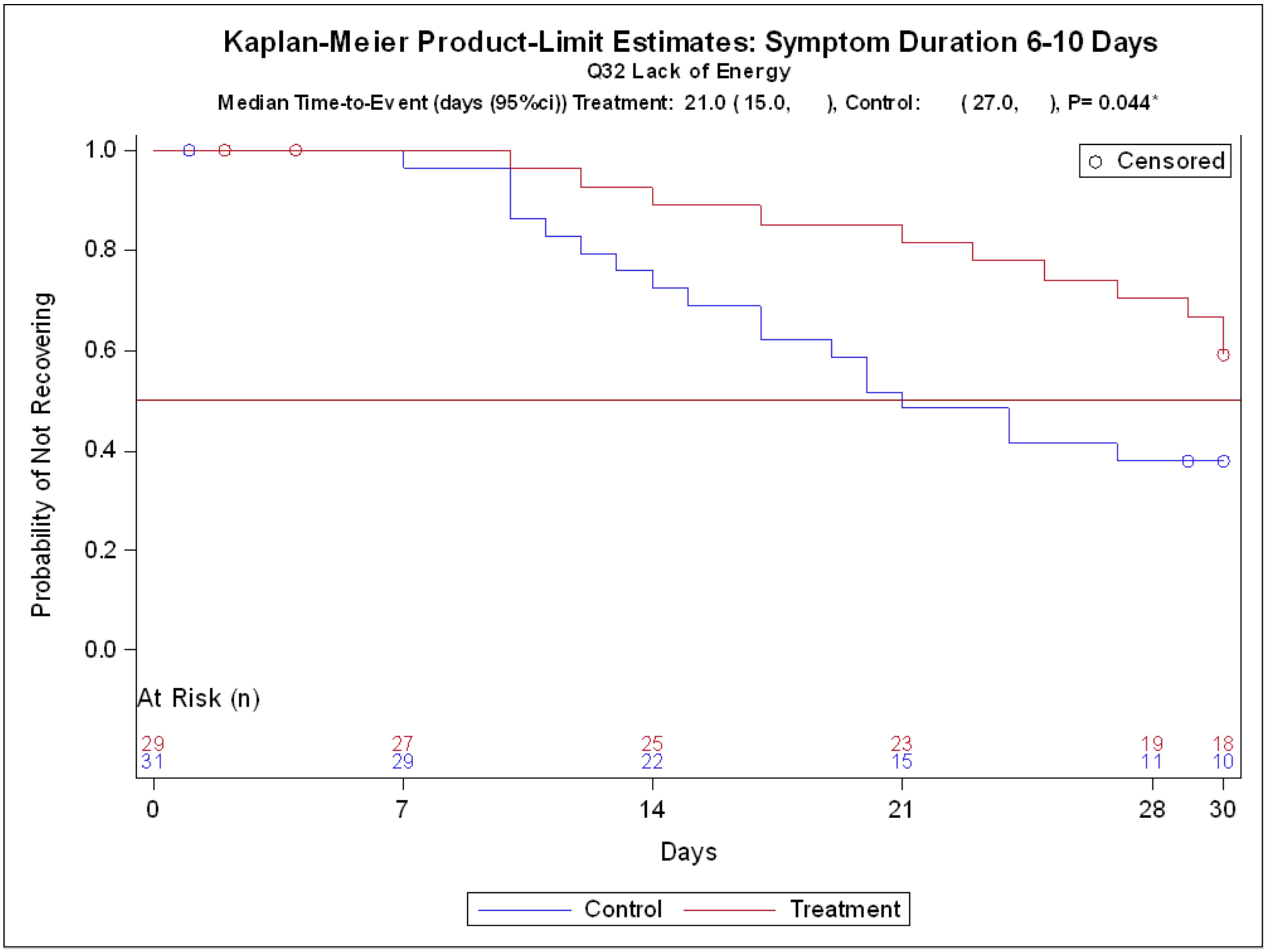

Kaplan-Meier Product-Limit Estimates: Symptom Duration 6-10 Days
Q32 Lack of Energy
Median Time-to-Event (days (95%ci)) Treatment: 21.0 ( 15.0, ), Control: ( 27.0, ), P= 0.044*
Censored
Probability of Not Recovering
1.0
0.8
0.6
0.4
0.2
0.0
At Risk (n)
29 27 25 23 19 18
31 29 22 15 11 10
0 7 14 21 28 30
Days
Control
Treatment
