## Supplementary material for "Home-use Photobiomodulation Device Treatment Outcomes for COVID-19": Cox Proportional Hazards Estimates

### Supplement 7

### Cox Proportional Hazards Estimates

**Table of Contents**

7.1 Full Table for Cox Proportional Hazards Estimates

7.2 Patient Demographics and Baseline Characteristics Response to Treatment - Univariate Cox Proportional Hazards Model in Symptom Duration 0-5 Days at Screening with Baseline Score of 4-7 for WURSS Question 1

### Supplement 7.1

#### Full Table for Cox Proportional Hazards Estimates

#### Success Criteria Event by Treatment Group in Patients with WURSS-44 Q1 Score of 4-7

| | | | | 95% Confidence Interval | | |
|---|---|---|---|---|---|---|
| WURSS Questionnaire Item | Symptom Days on Enrollment | N | Hazard Ratio | Lower | Upper | P-Value |
| 01: How sick do you feel today? | 0-5d | 136 | 1.495 | 0.996 | 2.243 | 0.052 |
| | 6-10d | 63 | 0.803 | 0.425 | 1.517 | 0.499 |
| | 0-10d | 199 | 1.252 | 0.888 | 1.764 | 0.199 |
| 02: Cough | 0-5d | 82 | 1.166 | 0.693 | 1.961 | 0.562 |
| | 6-10d | 36 | 0.726 | 0.319 | 1.654 | 0.446 |
| | 0-10d | 118 | 1.02 | 0.658 | 1.58 | 0.931 |
| 03: Coughing stuff up | 0-5d | 40 | 1.816 | 0.875 | 3.767 | 0.109 |
| | 6-10d | 19 | 1.819 | 0.684 | 4.837 | 0.230 |
| | 0-10d | 59 | 1.817 | 1.013 | 3.261 | 0.045* |
| 04: Cough interfering with sleep | 0-5d | 33 | 2.02 | 0.929 | 4.392 | 0.076 |
| | 6-10d | 15 | 1.273 | 0.399 | 4.062 | 0.684 |
| | 0-10d | 48 | 1.752 | 0.929 | 3.303 | 0.083 |
| 05: Soar throat | 0-5d | 40 | 1.504 | 0.726 | 3.114 | 0.272 |
| | 6-10d | 19 | 1.065 | 0.382 | 2.968 | 0.904 |
| | 0-10d | 59 | 1.339 | 0.747 | 2.399 | 0.327 |
| 06: Scratchy throat | 0-5d | 36 | 1.436 | 0.679 | 3.039 | 0.344 |
| | 6-10d | 15 | 0.543 | 0.155 | 1.9 | 0.339 |
| | 0-10d | 51 | 1.102 | 0.585 | 2.075 | 0.764 |
| 07: Hoarseness | 0-5d | 38 | 1.212 | 0.595 | 2.47 | 0.596 |
| | 6-10d | 18 | 2.56 | 0.884 | 7.414 | 0.083 |

| | | | | | | |
|---|---|---|---|---|---|---|
| | 0-10d | 56 | 1.521 | 0.839 | 2.759 | 0.167 |
| 08: Runny nose | 0-5d | 44 | 0.831 | 0.423 | 1.634 | 0.592 |
| | 6-10d | 17 | 3.619 | 0.924 | 14.179 | 0.065 |
| | 0-10d | 61 | 1.151 | 0.627 | 2.11 | 0.650 |
| 09: Plugged nose | 0-5d | 63 | 1.338 | 0.756 | 2.368 | 0.317 |
| | 6-10d | 26 | 0.986 | 0.417 | 2.335 | 0.975 |
| | 0-10d | 89 | 1.22 | 0.758 | 1.964 | 0.412 |
| 10: Sneezing | 0-5d | 39 | 1.777 | 0.862 | 3.663 | 0.119 |
| | 6-10d | 16 | 2.478 | 0.531 | 11.569 | 0.248 |
| | 0-10d | 55 | 1.896 | 0.99 | 3.628 | 0.054 |
| 11: Headache | 0-5d | 90 | 2.027 | 1.216 | 3.378 | 0.007* |
| | 6-10d | 38 | 0.824 | 0.361 | 1.879 | 0.645 |
| | 0-10d | 128 | 1.586 | 1.036 | 2.429 | 0.034* |
| 12: Body aches | 0-5d | 85 | 1.641 | 0.996 | 2.704 | 0.052 |
| | 6-10d | 34 | 1.681 | 0.767 | 3.684 | 0.195 |
| | 0-10d | 119 | 1.652 | 1.084 | 2.519 | 0.020* |
| 13: Feeling run down | 0-5d | 122 | 1.306 | 0.833 | 2.049 | 0.244 |
| | 6-10d | 54 | 0.506 | 0.245 | 1.046 | 0.066 |
| | 0-10d | 176 | 0.998 | 0.683 | 1.458 | 0.991 |
| 14: Sweats | 0-5d | 57 | 1.277 | 0.731 | 2.228 | 0.390 |
| | 6-10d | 21 | 1.129 | 0.456 | 2.799 | 0.793 |
| | 0-10d | 78 | 1.234 | 0.768 | 1.982 | 0.384 |
| 15: Chills | 0-5d | 44 | 0.674 | 0.355 | 1.279 | 0.227 |
| | 6-10d | 17 | 1.484 | 0.496 | 4.436 | 0.480 |
| | 0-10d | 61 | 0.827 | 0.468 | 1.461 | 0.513 |
| 16: Feeling feverish | 0-5d | 44 | 1.439 | 0.742 | 2.79 | 0.281 |
| | 6-10d | 17 | 1.09 | 0.362 | 3.284 | 0.878 |
| | 0-10d | 61 | 1.338 | 0.76 | 2.358 | 0.313 |
| 17: Feeling dizzy | 0-5d | 59 | 1.418 | 0.796 | 2.524 | 0.236 |
| | 6-10d | 25 | 0.923 | 0.363 | 2.343 | 0.866 |

| | | | | | | |
|---|---|---|---|---|---|---|
| | 0-10d | 84 | 1.261 | 0.768 | 2.07 | 0.359 |
| 18: Feeling tired | 0-5d | 125 | 1.09 | 0.677 | 1.755 | 0.723 |
| | 6-10d | 54 | 0.432 | 0.192 | 0.972 | 0.043* |
| | 0-10d | 179 | 0.849 | 0.566 | 1.275 | 0.431 |
| 19: Irritability | 0-5d | 68 | 1.717 | 0.947 | 3.114 | 0.075 |
| | 6-10d | 29 | 0.957 | 0.395 | 2.319 | 0.923 |
| | 0-10d | 97 | 1.438 | 0.874 | 2.367 | 0.153 |
| 20: Sinus pain | 0-5d | 60 | 1.926 | 1.074 | 3.452 | 0.028* |
| | 6-10d | 21 | 2.26 | 0.792 | 6.452 | 0.128 |
| | 0-10d | 81 | 2.001 | 1.203 | 3.328 | 0.008* |
| 21: Sinus pressure | 0-5d | 65 | 1.702 | 0.952 | 3.044 | 0.073 |
| | 6-10d | 28 | 0.906 | 0.368 | 2.234 | 0.831 |
| | 0-10d | 93 | 1.415 | 0.865 | 2.314 | 0.167 |
| 22: Sinus drainage | 0-5d | 40 | 2.158 | 0.994 | 4.687 | 0.052 |
| | 6-10d | 15 | 2.077 | 0.526 | 8.198 | 0.297 |
| | 0-10d | 55 | 2.138 | 1.088 | 4.203 | 0.028* |
| 23: Swollen glands | 0-5d | 33 | 2.436 | 1.046 | 5.676 | 0.039* |
| | 6-10d | 10 | 0.39 | 0.045 | 3.352 | 0.391 |
| | 0-10d | 43 | 1.764 | 0.861 | 3.616 | 0.121 |
| 24: Plugged ears | 0-5d | 29 | 1.542 | 0.673 | 3.532 | 0.306 |
| | 6-10d | 13 | 2.839 | 0.681 | 11.831 | 0.152 |
| | 0-10d | 42 | 1.812 | 0.895 | 3.67 | 0.099 |
| 25: Ear discomfort | 0-5d | 28 | 2.39 | 0.937 | 6.097 | 0.068 |
| | 6-10d | 17 | 2.217 | 0.653 | 7.527 | 0.202 |
| | 0-10d | 45 | 2.325 | 1.104 | 4.893 | 0.026* |
| 26: Watery eyes | 0-5d | 12 | 2.934 | 0.72 | 11.954 | 0.133 |
| | 6-10d | 12 | 1.45 | 0.291 | 7.22 | 0.650 |
| | 0-10d | 24 | 2.218 | 0.749 | 6.565 | 0.150 |
| 27: Eye discomfort | 0-5d | 23 | 0.747 | 0.298 | 1.869 | 0.533 |
| | 6-10d | 12 | 0.594 | 0.154 | 2.29 | 0.449 |

| | | | | | | |
|---|---|---|---|---|---|---|
| | 0-10d | 35 | 0.695 | 0.323 | 1.492 | 0.350 |
| 28: Head congestion | 0-5d | 71 | 0.873 | 0.48 | 1.586 | 0.655 |
| | 6-10d | 30 | 1.132 | 0.459 | 2.792 | 0.787 |
| | 0-10d | 101 | 0.944 | 0.574 | 1.553 | 0.822 |
| 29: Chest congestion | 0-5d | 59 | 1.847 | 1.008 | 3.384 | 0.047* |
| | 6-10d | 19 | 1.985 | 0.659 | 5.984 | 0.223 |
| | 0-10d | 78 | 1.878 | 1.105 | 3.193 | 0.020* |
| 30: Chest tightness | 0-5d | 46 | 1.247 | 0.599 | 2.593 | 0.555 |
| | 6-10d | 27 | 1.297 | 0.552 | 3.048 | 0.551 |
| | 0-10d | 73 | 1.268 | 0.727 | 2.211 | 0.403 |
| 31: Heaviness in chest | 0-5d | 46 | 1.386 | 0.682 | 2.815 | 0.367 |
| | 6-10d | 24 | 0.819 | 0.344 | 1.953 | 0.653 |
| | 0-10d | 70 | 1.127 | 0.652 | 1.951 | 0.668 |
| 32: Lack of energy | 0-5d | 118 | 1.211 | 0.737 | 1.988 | 0.450 |
| | 6-10d | 52 | 0.459 | 0.207 | 1.016 | 0.055 |
| | 0-10d | 170 | 0.916 | 0.603 | 1.392 | 0.681 |
| 33: Loss of appetite | 0-5d | 74 | 1.202 | 0.705 | 2.052 | 0.499 |
| | 6-10d | 30 | 0.87 | 0.382 | 1.979 | 0.739 |
| | 0-10d | 104 | 1.093 | 0.697 | 1.714 | 0.699 |
| 34: Think clearly | 0-5d | 59 | 2.067 | 1.093 | 3.909 | 0.025* |
| | 6-10d | 26 | 1.514 | 0.563 | 4.072 | 0.411 |
| | 0-10d | 85 | 1.893 | 1.103 | 3.248 | 0.021* |
| 35: Speak clearly | 0-5d | 31 | 1.194 | 0.473 | 3.01 | 0.708 |
| | 6-10d | 16 | 2.797 | 0.734 | 10.657 | 0.132 |
| | 0-10d | 47 | 1.609 | 0.768 | 3.369 | 0.208 |
| 36: Sleep well | 0-5d | 75 | 1.453 | 0.823 | 2.566 | 0.197 |
| | 6-10d | 33 | 0.858 | 0.34 | 2.165 | 0.746 |
| | 0-10d | 108 | 1.261 | 0.777 | 2.046 | 0.348 |
| 37: Breathe easily | 0-5d | 64 | 1.535 | 0.832 | 2.833 | 0.170 |
| | 6-10d | 30 | 0.915 | 0.373 | 2.243 | 0.846 |

| | | | | | | |
|---|---|---|---|---|---|---|
| | 0-10d | 94 | 1.301 | 0.791 | 2.14 | 0.301 |
| 38: Walk, climb stairs, exercise | 0-5d | 86 | 1.313 | 0.779 | 2.213 | 0.307 |
| | 6-10d | 39 | 0.756 | 0.313 | 1.826 | 0.534 |
| | 0-10d | 125 | 1.136 | 0.725 | 1.782 | 0.577 |
| 39: Accomplish daily activities | 0-5d | 90 | 1.336 | 0.802 | 2.227 | 0.266 |
| | 6-10d | 39 | 0.858 | 0.372 | 1.98 | 0.719 |
| | 0-10d | 129 | 1.186 | 0.767 | 1.834 | 0.443 |
| 40: Work outside the home | 0-5d | 88 | 1.315 | 0.784 | 2.205 | 0.299 |
| | 6-10d | 39 | 0.855 | 0.383 | 1.911 | 0.702 |
| | 0-10d | 127 | 1.162 | 0.75 | 1.8 | 0.502 |
| 41: Work inside the home | 0-5d | 86 | 1.313 | 0.786 | 2.193 | 0.298 |
| | 6-10d | 39 | 1.042 | 0.468 | 2.323 | 0.919 |
| | 0-10d | 125 | 1.228 | 0.797 | 1.892 | 0.352 |
| 42: Interact with others | 0-5d | 89 | 1.225 | 0.752 | 1.996 | 0.415 |
| | 6-10d | 38 | 1.033 | 0.49 | 2.176 | 0.932 |
| | 0-10d | 127 | 1.164 | 0.774 | 1.753 | 0.466 |
| 43: Live your personal life | 0-5d | 96 | 1.451 | 0.879 | 2.397 | 0.146 |
| | 6-10d | 40 | 0.839 | 0.388 | 1.816 | 0.656 |
| | 0-10d | 136 | 1.233 | 0.81 | 1.878 | 0.328 |

*p<0.05 by proportional hazards maximum likelihood estimate for 0-5d and 6-10d, and stratified MLE estimate for 0-10d symptom days.

Success (event) criteria defined as first day of 3 consecutive days of a zero score (does not have this symptom, not at all).

17MAR22, T04_cox

### Supplement 7.2

#### Patient Demographics and Baseline Characteristics Response to Treatment

#### Univariate Cox Proportional Hazards Model in Symptom Duration 0-5 Days on Enrollment with Baseline Score of 4-7 for WURSS Question 1

| | | | 95% Confidence Interval | | |
|---|---|---|---|---|---|
| Subgroup | N | Hazard Ratio | Lower | Upper | P-Value |
| Male | 93 | 1.572 | 0.959 | 2.576 | 0.073 |
| Female | 43 | 1.359 | 0.664 | 2.781 | 0.401 |
| Caucasian | 114 | 1.609 | 1.029 | 2.517 | 0.037* |
| Non-Caucasian | 22 | 1.112 | 0.415 | 2.974 | 0.833 |
| Age >30 | 87 | 2.229 | 1.291 | 3.848 | 0.004* |
| Age <=30 | 49 | 0.731 | 0.392 | 1.363 | 0.325 |
| Age >35 median | 67 | 1.99 | 1.078 | 3.675 | 0.028* |
| Age <=35 median | 69 | 1.135 | 0.658 | 1.958 | 0.649 |
| Age >40 | 51 | 1.789 | 0.879 | 3.641 | 0.109 |
| Age <=40 | 85 | 1.414 | 0.857 | 2.332 | 0.175 |
| Age >50 | 18 | 1.492 | 0.436 | 5.105 | 0.523 |
| Age <=50 | 118 | 1.514 | 0.984 | 2.331 | 0.059 |
| Age >60 | 6 | 1.165 | 0.119 | 11.389 | 0.896 |
| Age <=60 | 130 | 1.517 | 1.003 | 2.296 | 0.049* |
| USA Site | 128 | 1.506 | 0.995 | 2.279 | 0.053 |
| Canada Site | 8 | 2.24 | 0.231 | 21.713 | 0.486 |
| Former Smoker | 22 | 1.522 | 0.525 | 4.412 | 0.439 |
| Non-Smoker | 114 | 1.502 | 0.968 | 2.331 | 0.070 |
| Screen 4 or 5 | 44 | 1.532 | 0.71 | 3.307 | 0.277 |
| Screen 6 or 7 | 92 | 1.499 | 0.928 | 2.421 | 0.098 |

| | | | | | |
|---|---|---|---|---|---|
| BMI 24+ | 88 | 1.819 | 1.102 | 3.001 | 0.019* |
| BMI <24 | 48 | 1.029 | 0.508 | 2.085 | 0.936 |
| BMI 27+ Obese | 63 | 1.939 | 1.08 | 3.482 | 0.027* |
| BMI <27 Non-obese | 73 | 1.189 | 0.676 | 2.09 | 0.548 |
| BMI 30+ Very obese | 37 | 2.233 | 1.009 | 4.942 | 0.048* |
| BMI <30 Not very obese | 99 | 1.29 | 0.803 | 2.073 | 0.292 |

*p<0.05 by proportional hazards maximum likelihood estimate. 17MAR22

Success (event) criteria defined as first day of 3 consecutive days of a zero score (does not have this symptom, not at all).
