## Supplementary material for "Home-use Photobiomodulation Device Treatment Outcomes for COVID-19": Full Table ANOVA Mild Symptoms

### Supplement 8

#### Full Table of Analysis of Variance for Days of Mild Symptoms
#### Comparison of Days with WURSS-44 Scores of 0-3 for All Symptoms
#### in Patients with Baseline WURSS-44 Q1 Score of 4-7

| | | | | | | | | Treatment Comparison | | | | |
|---|---|---|---|---|---|---|---|---|---|---|---|---|
| | | | | | | 95% c.i. | | Difference | | 95% c.i. | | |
| Questionnaire Item | Symptom Days | Treatment | N | LS Mean | Std Error | Lower | Upper | LS Mean | Std Error | Lower | Upper | P-Value |
| 01: How sick do you feel today? | 0-5 | Treatment | 68 | 23.78 | 0.77 | 22.25 | 25.31 | | | | | |
| | | Control | 68 | 22.78 | 0.77 | 21.25 | 24.31 | | | | | |
| | | Difference | 136 | | | | | 1.00 | 1.09 | -1.16 | 3.16 | 0.362 |
| | 5-10 | Treatment | 32 | 22.84 | 1.40 | 20.05 | 25.64 | | | | | |
| | | Control | 31 | 22.06 | 1.42 | 19.23 | 24.90 | | | | | |
| | | Difference | 63 | | | | | 0.78 | 1.99 | -3.20 | 4.76 | 0.697 |
| | 0-10 | Treatment | 100 | 23.33 | 0.71 | 21.93 | 24.74 | | | | | |
| | | Control | 99 | 22.40 | 0.72 | 20.99 | 23.82 | | | | | |
| | | Difference | 199 | | | | | 0.93 | 0.97 | -0.99 | 2.85 | 0.341 |
| 02: Cough | 0-5 | Treatment | 44 | 24.52 | 0.88 | 22.77 | 26.27 | | | | | |
| | | Control | 38 | 22.42 | 0.95 | 20.54 | 24.30 | | | | | |
| | | Difference | 82 | | | | | 2.10 | 1.29 | -0.47 | 4.67 | 0.108 |
| | 5-10 | Treatment | 18 | 23.22 | 1.90 | 19.37 | 27.08 | | | | | |
| | | Control | 18 | 20.67 | 1.90 | 16.81 | 24.52 | | | | | |
| | | Difference | 36 | | | | | 2.56 | 2.68 | -2.89 | 8.01 | 0.347 |
| | 0-10 | Treatment | 62 | 23.83 | 0.88 | 22.09 | 25.56 | | | | | |
| | | Control | 56 | 21.59 | 0.90 | 19.79 | 23.38 | | | | | |
| | | Difference | 118 | | | | | 2.24 | 1.21 | -0.15 | 4.63 | 0.066 |
| 03: Coughing stuff up | 0-5 | Treatment | 17 | 23.71 | 1.63 | 20.41 | 27.00 | | | | | |
| | | Control | 23 | 22.17 | 1.40 | 19.34 | 25.01 | | | | | |
| | | Difference | 40 | | | | | 1.53 | 2.15 | -2.82 | 5.88 | 0.480 |
| | 5-10 | Treatment | 10 | 25.60 | 1.77 | 21.86 | 29.34 | | | | | |
| | | Control | 9 | 21.56 | 1.87 | 17.61 | 25.50 | | | | | |
| | | Difference | 19 | | | | | 4.04 | 2.58 | -1.39 | 9.48 | 0.135 |
| | 0-10 | Treatment | 27 | 24.49 | 1.25 | 21.99 | 26.99 | | | | | |
| | | Control | 32 | 22.14 | 1.19 | 19.75 | 24.52 | | | | | |
| | | Difference | 59 | | | | | 2.35 | 1.67 | -1.00 | 5.70 | 0.165 |

*p<0.05 by one-way ANOVA within symptom day strata (0-5, 6-10) and ANOVA with factors treatment and strata for 0-10 days.
17MAR22,T05_mild

### Supplement 8

#### Full Table of Analysis of Variance for Days of Mild Symptoms
#### Comparison of Days with WURSS-44 Scores of 0-3 for All Symptoms
#### in Patients with Baseline WURSS-44 Q1 Score of 4-7

| | | | | | | | | Treatment Comparison | | | | |
|---|---|---|---|---|---|---|---|---|---|---|---|---|
| | | | | | | 95% c.i. | | Difference | | 95% c.i. | | |
| Questionnaire Item | Symptom Days | Treatment | N | LS Mean | Std Error | Lower | Upper | LS Mean | Std Error | Lower | Upper | P-Value |
| 04: Cough interfering with sleep | 0-5 | Treatment | 19 | 24.74 | 1.39 | 21.89 | 27.58 | | | | | |
| | | Control | 14 | 23.36 | 1.62 | 20.04 | 26.67 | | | | | |
| | | Difference | 33 | | | | | 1.38 | 2.14 | -2.99 | 5.75 | 0.524 |
| | 5-10 | Treatment | 6 | 22.83 | 3.26 | 15.80 | 29.87 | | | | | |
| | | Control | 9 | 18.00 | 2.66 | 12.26 | 23.74 | | | | | |
| | | Difference | 15 | | | | | 4.83 | 4.2 | -4.25 | 13.91 | 0.271 |
| | 0-10 | Treatment | 25 | 23.30 | 1.44 | 20.39 | 26.20 | | | | | |
| | | Control | 23 | 20.85 | 1.41 | 18.01 | 23.69 | | | | | |
| | | Difference | 48 | | | | | 2.45 | 1.95 | -1.49 | 6.38 | 0.217 |
| 05: Sore throat | 0-5 | Treatment | 25 | 24.68 | 1.34 | 21.97 | 27.39 | | | | | |
| | | Control | 15 | 22.93 | 1.73 | 19.44 | 26.43 | | | | | |
| | | Difference | 40 | | | | | 1.75 | 2.18 | -2.68 | 6.17 | 0.429 |
| | 5-10 | Treatment | 6 | 26.67 | 3.07 | 20.19 | 33.14 | | | | | |
| | | Control | 13 | 21.85 | 2.09 | 17.45 | 26.25 | | | | | |
| | | Difference | 19 | | | | | 4.82 | 3.71 | -3.01 | 12.65 | 0.211 |
| | 0-10 | Treatment | 31 | 25.12 | 1.39 | 22.33 | 27.90 | | | | | |
| | | Control | 28 | 22.43 | 1.31 | 19.81 | 25.06 | | | | | |
| | | Difference | 59 | | | | | 2.68 | 1.89 | -1.1 | 6.46 | 0.161 |
| 06: Scratchy throat | 0-5 | Treatment | 19 | 25.58 | 1.28 | 22.98 | 28.17 | | | | | |
| | | Control | 17 | 24.65 | 1.35 | 21.90 | 27.39 | | | | | |
| | | Difference | 36 | | | | | 0.93 | 1.86 | -2.84 | 4.71 | 0.619 |
| | 5-10 | Treatment | 6 | 26.50 | 3.15 | 19.71 | 33.29 | | | | | |
| | | Control | 9 | 20.33 | 2.57 | 14.79 | 25.88 | | | | | |
| | | Difference | 15 | | | | | 6.17 | 4.06 | -2.6 | 14.94 | 0.153 |
| | 0-10 | Treatment | 25 | 25.27 | 1.35 | 22.55 | 28.00 | | | | | |
| | | Control | 26 | 22.84 | 1.27 | 20.29 | 25.39 | | | | | |
| | | Difference | 51 | | | | | 2.43 | 1.77 | -1.13 | 5.99 | 0.176 |

*p<0.05 by one-way ANOVA within symptom day strata (0-5, 6-10) and ANOVA with factors treatment and strata for 0-10 days.
17MAR22,T05_mild

### Supplement 8

#### Full Table of Analysis of Variance for Days of Mild Symptoms
#### Comparison of Days with WURSS-44 Scores of 0-3 for All Symptoms
#### in Patients with Baseline WURSS-44 Q1 Score of 4-7

| | | | | | | | | Treatment Comparison | | | | |
|---|---|---|---|---|---|---|---|---|---|---|---|---|
| | | | | | | 95% c.i. | | Difference | | 95% c.i. | | |
| Questionnaire Item | Symptom Days | Treatment | N | LS Mean | Std Error | Lower | Upper | LS Mean | Std Error | Lower | Upper | P-Value |
| 10: Sneezing | 0-5 | Treatment | 22 | 25.86 | 1.23 | 23.37 | 28.35 | | | | | |
| | | Control | 17 | 24.65 | 1.40 | 21.81 | 27.48 | | | | | |
| | | Difference | 39 | | | | | 1.22 | 1.86 | -2.56 | 4.99 | 0.518 |
| | 5-10 | Treatment | 11 | 26.64 | 1.57 | 23.27 | 30.00 | | | | | |
| | | Control | 5 | 16.60 | 2.33 | 11.60 | 21.60 | | | | | |
| | | Difference | 16 | | | | | 10.04 | 2.81 | 4.01 | 16.06 | 0.003* |
| | 0-10 | Treatment | 33 | 25.74 | 1.07 | 23.60 | 27.88 | | | | | |
| | | Control | 22 | 22.20 | 1.35 | 19.50 | 24.90 | | | | | |
| | | Difference | 55 | | | | | 3.54 | 1.63 | 0.27 | 6.82 | 0.035* |
| 11: Headache | 0-5 | Treatment | 50 | 23.84 | 0.91 | 22.04 | 25.64 | | | | | |
| | | Control | 40 | 20.73 | 1.01 | 18.71 | 22.74 | | | | | |
| | | Difference | 90 | | | | | 3.11 | 1.36 | 0.41 | 5.82 | 0.024* |
| | 5-10 | Treatment | 17 | 19.94 | 1.75 | 16.39 | 23.50 | | | | | |
| | | Control | 21 | 19.57 | 1.58 | 16.37 | 22.77 | | | | | |
| | | Difference | 38 | | | | | 0.37 | 2.36 | -4.41 | 5.15 | 0.876 |
| | 0-10 | Treatment | 67 | 22.24 | 0.87 | 20.51 | 23.97 | | | | | |
| | | Control | 61 | 19.94 | 0.88 | 18.21 | 21.68 | | | | | |
| | | Difference | 128 | | | | | 2.30 | 1.19 | -0.05 | 4.65 | 0.055 |
| 12: Body aches | 0-5 | Treatment | 46 | 23.83 | 1.11 | 21.62 | 26.03 | | | | | |
| | | Control | 39 | 21.33 | 1.20 | 18.94 | 23.73 | | | | | |
| | | Difference | 85 | | | | | 2.49 | 1.64 | -0.76 | 5.75 | 0.131 |
| | 5-10 | Treatment | 14 | 24.93 | 2.12 | 20.61 | 29.25 | | | | | |
| | | Control | 20 | 18.95 | 1.77 | 15.34 | 22.56 | | | | | |
| | | Difference | 34 | | | | | 5.98 | 2.76 | 0.35 | 11.61 | 0.038* |
| | 0-10 | Treatment | 60 | 23.86 | 1.07 | 21.74 | 25.98 | | | | | |
| | | Control | 59 | 20.39 | 1.03 | 18.36 | 22.43 | | | | | |
| | | Difference | 119 | | | | | 3.47 | 1.41 | 0.68 | 6.26 | 0.015* |

*p<0.05 by one-way ANOVA within symptom day strata (0-5, 6-10) and ANOVA with factors treatment and strata for 0-10 days.

17MAR22,T05_mild

### Supplement 8

#### Full Table of Analysis of Variance for Days of Mild Symptoms
#### Comparison of Days with WURSS-44 Scores of 0-3 for All Symptoms
#### in Patients with Baseline WURSS-44 Q1 Score of 4-7

| | | | | | | | | Treatment Comparison | | | | |
|---|---|---|---|---|---|---|---|---|---|---|---|---|
| | | | | | | 95% c.i. | | Difference | | 95% c.i. | | |
| Questionnaire Item | Symptom Days | Treatment | N | LS Mean | Std Error | Lower | Upper | LS Mean | Std Error | Lower | Upper | P-Value |
| 13: Feeling run down | 0-5 | Treatment | 61 | 21.41 | 1.08 | 19.27 | 23.55 | | | | | |
| | | Control | 61 | 19.56 | 1.08 | 17.42 | 21.70 | | | | | |
| | | Difference | 122 | | | | | 1.85 | 1.53 | -1.17 | 4.88 | 0.228 |
| | 5-10 | Treatment | 26 | 19.00 | 1.89 | 15.21 | 22.79 | | | | | |
| | | Control | 28 | 18.54 | 1.82 | 14.89 | 22.19 | | | | | |
| | | Difference | 54 | | | | | 0.46 | 2.62 | -4.80 | 5.73 | 0.860 |
| | 0-10 | Treatment | 87 | 20.35 | 0.99 | 18.40 | 22.29 | | | | | |
| | | Control | 89 | 18.92 | 0.97 | 17.01 | 20.83 | | | | | |
| | | Difference | 176 | | | | | 1.43 | 1.33 | -1.19 | 4.04 | 0.283 |
| 14: Sweats | 0-5 | Treatment | 31 | 24.32 | 0.95 | 22.42 | 26.23 | | | | | |
| | | Control | 26 | 25.92 | 1.04 | 23.85 | 28.00 | | | | | |
| | | Difference | 57 | | | | | -1.60 | 1.41 | -4.42 | 1.22 | 0.260 |
| | 5-10 | Treatment | 10 | 23.10 | 2.29 | 18.31 | 27.89 | | | | | |
| | | Control | 11 | 24.18 | 2.18 | 19.61 | 28.75 | | | | | |
| | | Difference | 21 | | | | | -1.08 | 3.16 | -7.70 | 5.54 | 0.736 |
| | 0-10 | Treatment | 41 | 23.64 | 0.98 | 21.68 | 25.61 | | | | | |
| | | Control | 37 | 25.10 | 1.00 | 23.11 | 27.10 | | | | | |
| | | Difference | 78 | | | | | -1.46 | 1.32 | -4.09 | 1.17 | 0.272 |
| 15: Chills | 0-5 | Treatment | 28 | 24.46 | 1.21 | 22.03 | 26.90 | | | | | |
| | | Control | 16 | 24.69 | 1.60 | 21.47 | 27.91 | | | | | |
| | | Difference | 44 | | | | | -0.22 | 2.00 | -4.26 | 3.82 | 0.912 |
| | 5-10 | Treatment | 8 | 22.25 | 3.37 | 15.07 | 29.43 | | | | | |
| | | Control | 9 | 21.11 | 3.18 | 14.34 | 27.88 | | | | | |
| | | Difference | 17 | | | | | 1.14 | 4.63 | -8.73 | 11.01 | 0.809 |
| | 0-10 | Treatment | 36 | 23.18 | 1.35 | 20.48 | 25.87 | | | | | |
| | | Control | 25 | 23.00 | 1.49 | 20.02 | 25.97 | | | | | |
| | | Difference | 61 | | | | | 0.18 | 1.92 | -3.66 | 4.02 | 0.927 |

*p<0.05 by one-way ANOVA within symptom day strata (0-5, 6-10) and ANOVA with factors treatment and strata for 0-10 days.
17MAR22,T05_mild

### Supplement 8

#### Full Table of Analysis of Variance for Days of Mild Symptoms
#### Comparison of Days with WURSS-44 Scores of 0-3 for All Symptoms
#### in Patients with Baseline WURSS-44 Q1 Score of 4-7

| | | | | | | | | Treatment Comparison | | | | |
|---|---|---|---|---|---|---|---|---|---|---|---|---|
| | | | | | | 95% c.i. | | Difference | | 95% c.i. | | |
| Questionnaire Item | Symptom Days | Treatment | N | LS Mean | Std Error | Lower | Upper | LS Mean | Std Error | Lower | Upper | P-Value |
| 16: Feeling feverish | 0-5 | Treatment | 26 | 24.08 | 1.19 | 21.67 | 26.49 | | | | | |
| | | Control | 18 | 24.72 | 1.44 | 21.83 | 27.62 | | | | | |
| | | Difference | 44 | | | | | -0.65 | 1.87 | -4.41 | 3.12 | 0.731 |
| | 5-10 | Treatment | 8 | 22.25 | 3.36 | 15.08 | 29.42 | | | | | |
| | | Control | 9 | 20.67 | 3.17 | 13.91 | 27.43 | | | | | |
| | | Difference | 17 | | | | | 1.58 | 4.62 | -8.27 | 11.44 | 0.737 |
| | 0-10 | Treatment | 34 | 22.87 | 1.33 | 20.20 | 25.54 | | | | | |
| | | Control | 27 | 22.88 | 1.41 | 20.06 | 25.70 | | | | | |
| | | Difference | 61 | | | | | -0.01 | 1.84 | -3.70 | 3.68 | 0.995 |
| 17: Feeling dizzy | 0-5 | Treatment | 27 | 23.30 | 1.33 | 20.63 | 25.96 | | | | | |
| | | Control | 32 | 23.19 | 1.22 | 20.74 | 25.63 | | | | | |
| | | Difference | 59 | | | | | 0.11 | 1.81 | -3.51 | 3.72 | 0.952 |
| | 5-10 | Treatment | 13 | 22.31 | 2.57 | 16.99 | 27.62 | | | | | |
| | | Control | 12 | 19.83 | 2.67 | 14.30 | 25.37 | | | | | |
| | | Difference | 25 | | | | | 2.47 | 3.71 | -5.20 | 10.15 | 0.511 |
| | 0-10 | Treatment | 40 | 22.60 | 1.25 | 20.11 | 25.08 | | | | | |
| | | Control | 44 | 21.78 | 1.22 | 19.35 | 24.21 | | | | | |
| | | Difference | 84 | | | | | 0.82 | 1.67 | -2.51 | 4.14 | 0.627 |
| 18: Feeling tired | 0-5 | Treatment | 62 | 20.89 | 1.06 | 18.79 | 22.98 | | | | | |
| | | Control | 63 | 19.11 | 1.05 | 17.03 | 21.19 | | | | | |
| | | Difference | 125 | | | | | 1.78 | 1.49 | -1.18 | 4.73 | 0.236 |
| | 5-10 | Treatment | 25 | 17.44 | 1.99 | 13.45 | 21.43 | | | | | |
| | | Control | 29 | 17.90 | 1.84 | 14.19 | 21.60 | | | | | |
| | | Difference | 54 | | | | | -0.46 | 2.71 | -5.90 | 4.98 | 0.867 |
| | 0-10 | Treatment | 87 | 19.41 | 1.00 | 17.45 | 21.38 | | | | | |
| | | Control | 92 | 18.31 | 0.96 | 16.42 | 20.20 | | | | | |
| | | Difference | 179 | | | | | 1.11 | 1.32 | -1.50 | 3.71 | 0.404 |

*p<0.05 by one-way ANOVA within symptom day strata (0-5, 6-10) and ANOVA with factors treatment and strata for 0-10 days.
17MAR22,T05_mild

### Supplement 8

#### Full Table of Analysis of Variance for Days of Mild Symptoms
#### Comparison of Days with WURSS-44 Scores of 0-3 for All Symptoms
#### in Patients with Baseline WURSS-44 Q1 Score of 4-7

| | | | | | | | | Treatment Comparison | | | | |
|---|---|---|---|---|---|---|---|---|---|---|---|---|
| | | | | | | 95% c.i. | | Difference | | 95% c.i. | | |
| Questionnaire Item | Symptom Days | Treatment | N | LS Mean | Std Error | Lower | Upper | LS Mean | Std Error | Lower | Upper | P-Value |
| 19: Irritability | 0-5 | Treatment | 33 | 23.06 | 1.33 | 20.41 | 25.71 | | | | | |
| | | Control | 35 | 19.49 | 1.29 | 16.92 | 22.06 | | | | | |
| | | Difference | 68 | | | | | 3.57 | 1.85 | -0.11 | 7.26 | 0.057 |
| | 5-10 | Treatment | 16 | 21.19 | 2.22 | 16.62 | 25.75 | | | | | |
| | | Control | 13 | 16.85 | 2.47 | 11.78 | 21.91 | | | | | |
| | | Difference | 29 | | | | | 4.34 | 3.32 | -2.47 | 11.16 | 0.202 |
| | 0-10 | Treatment | 49 | 22.06 | 1.18 | 19.72 | 24.40 | | | | | |
| | | Control | 48 | 18.26 | 1.22 | 15.84 | 20.68 | | | | | |
| | | Difference | 97 | | | | | 3.80 | 1.62 | 0.58 | 7.02 | 0.021* |
| 20: Sinus pain | 0-5 | Treatment | 28 | 23.86 | 1.33 | 21.20 | 26.52 | | | | | |
| | | Control | 32 | 22.84 | 1.24 | 20.35 | 25.33 | | | | | |
| | | Difference | 60 | | | | | 1.01 | 1.82 | -2.63 | 4.66 | 0.580 |
| | 5-10 | Treatment | 11 | 24.82 | 2.38 | 19.84 | 29.79 | | | | | |
| | | Control | 10 | 19.50 | 2.49 | 14.28 | 24.72 | | | | | |
| | | Difference | 21 | | | | | 5.32 | 3.44 | -1.89 | 12.53 | 0.139 |
| | 0-10 | Treatment | 39 | 23.88 | 1.23 | 21.42 | 26.33 | | | | | |
| | | Control | 42 | 21.75 | 1.22 | 19.31 | 24.18 | | | | | |
| | | Difference | 81 | | | | | 2.13 | 1.62 | -1.09 | 5.35 | 0.192 |
| 21: Sinus pressure | 0-5 | Treatment | 28 | 23.71 | 1.35 | 21.02 | 26.41 | | | | | |
| | | Control | 37 | 23.19 | 1.17 | 20.85 | 25.53 | | | | | |
| | | Difference | 65 | | | | | 0.53 | 1.79 | -3.04 | 4.09 | 0.770 |
| | 5-10 | Treatment | 13 | 23.77 | 2.35 | 18.94 | 28.60 | | | | | |
| | | Control | 15 | 21.13 | 2.19 | 16.64 | 25.63 | | | | | |
| | | Difference | 28 | | | | | 2.64 | 3.21 | -3.96 | 9.23 | 0.419 |
| | 0-10 | Treatment | 41 | 23.53 | 1.21 | 21.12 | 25.94 | | | | | |
| | | Control | 52 | 22.36 | 1.10 | 20.17 | 24.56 | | | | | |
| | | Difference | 93 | | | | | 1.17 | 1.57 | -1.95 | 4.29 | 0.460 |

*p<0.05 by one-way ANOVA within symptom day strata (0-5, 6-10) and ANOVA with factors treatment and strata for 0-10 days.

17MAR22,T05_mild

### Supplement 8

#### Full Table of Analysis of Variance for Days of Mild Symptoms
#### Comparison of Days with WURSS-44 Scores of 0-3 for All Symptoms
#### in Patients with Baseline WURSS-44 Q1 Score of 4-7

| | | | | | | | | Treatment Comparison | | | | |
|---|---|---|---|---|---|---|---|---|---|---|---|---|
| | | | | | | 95% c.i. | | Difference | | 95% c.i. | | |
| Questionnaire Item | Symptom Days | Treatment | N | LS Mean | Std Error | Lower | Upper | LS Mean | Std Error | Lower | Upper | P-Value |
| 22: Sinus drainage | 0-5 | Treatment | 19 | 21.58 | 1.79 | 17.95 | 25.2 | | | | | |
| | | Control | 21 | 22.81 | 1.7 | 19.36 | 26.26 | | | | | |
| | | Difference | 40 | | | | | -1.23 | 2.47 | -6.23 | 3.77 | 0.621 |
| | 5-10 | Treatment | 9 | 26.44 | 2.42 | 21.21 | 31.68 | | | | | |
| | | Control | 6 | 16.50 | 2.97 | 10.09 | 22.91 | | | | | |
| | | Difference | 15 | | | | | 9.94 | 3.83 | 1.66 | 18.23 | 0.022* |
| | 0-10 | Treatment | 28 | 23.15 | 1.57 | 19.99 | 26.31 | | | | | |
| | | Control | 27 | 21.41 | 1.68 | 18.04 | 24.79 | | | | | |
| | | Difference | 55 | | | | | 1.73 | 2.17 | -2.63 | 6.09 | 0.429 |
| 23: Swollen glands | 0-5 | Treatment | 20 | 25.05 | 1.28 | 22.45 | 27.65 | | | | | |
| | | Control | 13 | 25.00 | 1.58 | 21.77 | 28.23 | | | | | |
| | | Difference | 33 | | | | | 0.05 | 2.03 | -4.10 | 4.20 | 0.981 |
| | 5-10 | Treatment | 3 | 20.67 | 4.92 | 9.32 | 32.02 | | | | | |
| | | Control | 7 | 19.14 | 3.22 | 11.71 | 26.57 | | | | | |
| | | Difference | 10 | | | | | 1.52 | 5.88 | -12.04 | 15.09 | 0.802 |
| | 0-10 | Treatment | 23 | 22.51 | 1.58 | 19.32 | 25.7 | | | | | |
| | | Control | 20 | 22.15 | 1.46 | 19.21 | 25.09 | | | | | |
| | | Difference | 43 | | | | | 0.36 | 2.00 | -3.68 | 4.40 | 0.858 |
| 24: Plugged ears | 0-5 | Treatment | 12 | 23.92 | 1.75 | 20.33 | 27.5 | | | | | |
| | | Control | 17 | 22.53 | 1.47 | 19.52 | 25.54 | | | | | |
| | | Difference | 29 | | | | | 1.39 | 2.28 | -3.30 | 6.07 | 0.548 |
| | 5-10 | Treatment | 8 | 26.00 | 2.26 | 21.02 | 30.98 | | | | | |
| | | Control | 5 | 17.80 | 2.86 | 11.5 | 24.1 | | | | | |
| | | Difference | 13 | | | | | 8.20 | 3.65 | 0.17 | 16.23 | 0.046* |
| | 0-10 | Treatment | 20 | 24.65 | 1.42 | 21.78 | 27.53 | | | | | |
| | | Control | 22 | 21.19 | 1.46 | 18.24 | 24.15 | | | | | |
| | | Difference | 42 | | | | | 3.46 | 1.98 | -0.54 | 7.46 | 0.088 |

*p<0.05 by one-way ANOVA within symptom day strata (0-5, 6-10) and ANOVA with factors treatment and strata for 0-10 days.
17MAR22,T05_mild

### Supplement 8

#### Full Table of Analysis of Variance for Days of Mild Symptoms
#### Comparison of Days with WURSS-44 Scores of 0-3 for All Symptoms
#### in Patients with Baseline WURSS-44 Q1 Score of 4-7

| | | | | | | | | Treatment Comparison | | | | |
|---|---|---|---|---|---|---|---|---|---|---|---|---|
| | | | | | | 95% c.i. | | Difference | | 95% c.i. | | |
| Questionnaire Item | Symptom Days | Treatment | N | LS Mean | Std Error | Lower | Upper | LS Mean | Std Error | Lower | Upper | P-Value |
| 25: Ear discomfort | 0-5 | Treatment | 14 | 25.50 | 1.40 | 22.63 | 28.37 | | | | | |
| | | Control | 14 | 21.79 | 1.40 | 18.91 | 24.66 | | | | | |
| | | Difference | 28 | | | | | 3.71 | 1.98 | -0.35 | 7.78 | 0.072 |
| | 5-10 | Treatment | 10 | 25.30 | 2.04 | 20.95 | 29.65 | | | | | |
| | | Control | 7 | 21.71 | 2.44 | 16.51 | 26.92 | | | | | |
| | | Difference | 17 | | | | | 3.59 | 3.18 | -3.20 | 10.37 | 0.278 |
| | 0-10 | Treatment | 24 | 25.40 | 1.16 | 23.06 | 27.75 | | | | | |
| | | Control | 21 | 21.74 | 1.27 | 19.18 | 24.29 | | | | | |
| | | Difference | 45 | | | | | 3.67 | 1.69 | 0.25 | 7.08 | 0.036* |
| 26: Watery eyes | 0-5 | Treatment | 7 | 21.71 | 3.51 | 13.89 | 29.54 | | | | | |
| | | Control | 5 | 19.40 | 4.15 | 10.15 | 28.65 | | | | | |
| | | Difference | 12 | | | | | 2.31 | 5.44 | -9.80 | 14.43 | 0.679 |
| | 5-10 | Treatment | 8 | 25.25 | 2.17 | 20.42 | 30.08 | | | | | |
| | | Control | 4 | 18.25 | 3.07 | 11.42 | 25.08 | | | | | |
| | | Difference | 12 | | | | | 7.00 | 3.76 | -1.37 | 15.37 | 0.092 |
| | 0-10 | Treatment | 15 | 23.54 | 2.01 | 19.36 | 27.72 | | | | | |
| | | Control | 9 | 18.99 | 2.60 | 13.59 | 24.39 | | | | | |
| | | Difference | 24 | | | | | 4.55 | 3.29 | -2.29 | 11.40 | 0.181 |
| 27: Eye discomfort | 0-5 | Treatment | 10 | 25.50 | 1.85 | 21.66 | 29.34 | | | | | |
| | | Control | 13 | 24.00 | 1.62 | 20.63 | 27.37 | | | | | |
| | | Difference | 23 | | | | | 1.50 | 2.46 | -3.61 | 6.61 | 0.548 |
| | 5-10 | Treatment | 7 | 23.86 | 2.78 | 17.65 | 30.06 | | | | | |
| | | Control | 5 | 21.00 | 3.30 | 13.66 | 28.34 | | | | | |
| | | Difference | 12 | | | | | 2.86 | 4.31 | -6.76 | 12.47 | 0.523 |
| | 0-10 | Treatment | 17 | 24.62 | 1.54 | 21.49 | 27.75 | | | | | |
| | | Control | 18 | 22.66 | 1.56 | 19.48 | 25.85 | | | | | |
| | | Difference | 35 | | | | | 1.96 | 2.15 | -2.41 | 6.33 | 0.367 |

*p<0.05 by one-way ANOVA within symptom day strata (0-5, 6-10) and ANOVA with factors treatment and strata for 0-10 days.
17MAR22,T05_mild

### Supplement 8

#### Full Table of Analysis of Variance for Days of Mild Symptoms
#### Comparison of Days with WURSS-44 Scores of 0-3 for All Symptoms
#### in Patients with Baseline WURSS-44 Q1 Score of 4-7

| | | | | | | | | Treatment Comparison | | | | |
|---|---|---|---|---|---|---|---|---|---|---|---|---|
| | | | | | | 95% c.i. | | Difference | | 95% c.i. | | |
| Questionnaire Item | Symptom Days | Treatment | N | LS Mean | Std Error | Lower | Upper | LS Mean | Std Error | Lower | Upper | P-Value |
| 28: Head congestion | 0-5 | Treatment | 32 | 22.59 | 1.25 | 20.11 | 25.08 | | | | | |
| | | Control | 39 | 23.18 | 1.13 | 20.93 | 25.43 | | | | | |
| | | Difference | 71 | | | | | -0.59 | 1.68 | -3.94 | 2.77 | 0.729 |
| | 5-10 | Treatment | 13 | 23.92 | 2.12 | 19.57 | 28.27 | | | | | |
| | | Control | 17 | 22.35 | 1.86 | 18.55 | 26.16 | | | | | |
| | | Difference | 30 | | | | | 1.57 | 2.82 | -4.21 | 7.35 | 0.582 |
| | 0-10 | Treatment | 45 | 23.00 | 1.13 | 20.77 | 25.24 | | | | | |
| | | Control | 56 | 22.95 | 1.01 | 20.94 | 24.96 | | | | | |
| | | Difference | 101 | | | | | 0.05 | 1.44 | -2.82 | 2.92 | 0.972 |
| 29: Chest congestion | 0-5 | Treatment | 32 | 23.69 | 1.26 | 21.16 | 26.21 | | | | | |
| | | Control | 27 | 22.26 | 1.37 | 19.51 | 25.01 | | | | | |
| | | Difference | 59 | | | | | 1.43 | 1.86 | -2.30 | 5.16 | 0.447 |
| | 5-10 | Treatment | 11 | 25.82 | 1.25 | 23.18 | 28.45 | | | | | |
| | | Control | 8 | 22.63 | 1.46 | 19.54 | 25.71 | | | | | |
| | | Difference | 19 | | | | | 3.19 | 1.92 | -0.87 | 7.25 | 0.115 |
| | 0-10 | Treatment | 43 | 24.57 | 1.08 | 22.41 | 26.72 | | | | | |
| | | Control | 35 | 22.72 | 1.20 | 20.33 | 25.10 | | | | | |
| | | Difference | 78 | | | | | 1.85 | 1.49 | -1.11 | 4.82 | 0.217 |
| 30: Chest tightness | 0-5 | Treatment | 25 | 21.84 | 1.56 | 18.70 | 24.98 | | | | | |
| | | Control | 21 | 21.19 | 1.70 | 17.76 | 24.62 | | | | | |
| | | Difference | 46 | | | | | 0.65 | 2.31 | -4.01 | 5.30 | 0.780 |
| | 5-10 | Treatment | 16 | 25.06 | 1.21 | 22.56 | 27.56 | | | | | |
| | | Control | 11 | 23.73 | 1.46 | 20.71 | 26.74 | | | | | |
| | | Difference | 27 | | | | | 1.34 | 1.90 | -2.58 | 5.25 | 0.489 |
| | 0-10 | Treatment | 41 | 23.42 | 1.08 | 21.26 | 25.58 | | | | | |
| | | Control | 32 | 22.52 | 1.24 | 20.06 | 24.98 | | | | | |
| | | Difference | 73 | | | | | 0.90 | 1.61 | -2.32 | 4.12 | 0.579 |

*p<0.05 by one-way ANOVA within symptom day strata (0-5, 6-10) and ANOVA with factors treatment and strata for 0-10 days.
17MAR22,T05_mild

### Supplement 8

#### Full Table of Analysis of Variance for Days of Mild Symptoms Comparison of Days with WURSS-44 Scores of 0-3 for All Symptoms in Patients with Baseline WURSS-44 Q1 Score of 4-7

| | | | | | | | | Treatment Comparison | | | | |
|---|---|---|---|---|---|---|---|---|---|---|---|---|
| | | | | | | 95% c.i. | | Difference | | 95% c.i. | | |
| Questionnaire Item | Symptom Days | Treatment | N | LS Mean | Std Error | Lower | Upper | LS Mean | Std Error | Lower | Upper | P-Value |
| 31: Heaviness in chest | 0-5 | Treatment | 24 | 21.42 | 1.62 | 18.16 | 24.67 | | | | | |
| | | Control | 22 | 21.09 | 1.69 | 17.69 | 24.49 | | | | | |
| | | Difference | 46 | | | | | 0.33 | 2.34 | -4.39 | 5.04 | 0.890 |
| | 5-10 | Treatment | 14 | 24.64 | 1.35 | 21.84 | 27.45 | | | | | |
| | | Control | 10 | 23.9 | 1.6 | 20.58 | 27.22 | | | | | |
| | | Difference | 24 | | | | | 0.74 | 2.10 | -3.61 | 5.09 | 0.726 |
| | 0-10 | Treatment | 38 | 23.01 | 1.17 | 20.68 | 25.33 | | | | | |
| | | Control | 32 | 22.54 | 1.29 | 19.97 | 25.11 | | | | | |
| | | Difference | 70 | | | | | 0.47 | 1.69 | -2.91 | 3.85 | 0.784 |
| 32: Lack of energy | 0-5 | Treatment | 59 | 20.49 | 1.11 | 18.29 | 22.69 | | | | | |
| | | Control | 59 | 19.59 | 1.11 | 17.39 | 21.79 | | | | | |
| | | Difference | 118 | | | | | 0.90 | 1.57 | -2.22 | 4.01 | 0.569 |
| | 5-10 | Treatment | 25 | 17.6 | 1.96 | 13.67 | 21.53 | | | | | |
| | | Control | 27 | 17.44 | 1.88 | 13.66 | 21.23 | | | | | |
| | | Difference | 52 | | | | | 0.16 | 2.72 | -5.30 | 5.61 | 0.955 |
| | 0-10 | Treatment | 84 | 19.12 | 1.02 | 17.12 | 21.13 | | | | | |
| | | Control | 86 | 18.45 | 1 | 16.48 | 20.42 | | | | | |
| | | Difference | 170 | | | | | 0.67 | 1.37 | -2.03 | 3.37 | 0.624 |
| 33: Loss of appetite | 0-5 | Treatment | 36 | 22.97 | 1.14 | 20.7 | 25.24 | | | | | |
| | | Control | 38 | 22.63 | 1.11 | 20.42 | 24.84 | | | | | |
| | | Difference | 74 | | | | | 0.34 | 1.59 | -2.83 | 3.51 | 0.831 |
| | 5-10 | Treatment | 15 | 24.67 | 2.16 | 20.25 | 29.09 | | | | | |
| | | Control | 15 | 20.6 | 2.16 | 16.18 | 25.02 | | | | | |
| | | Difference | 30 | | | | | 4.07 | 3.05 | -2.18 | 10.32 | 0.193 |
| | 0-10 | Treatment | 51 | 23.43 | 1.07 | 21.3 | 25.56 | | | | | |
| | | Control | 53 | 22.02 | 1.06 | 19.91 | 24.12 | | | | | |
| | | Difference | 104 | | | | | 1.42 | 1.43 | -1.43 | 4.26 | 0.326 |

*p<0.05 by one-way ANOVA within symptom day strata (0-5, 6-10) and ANOVA with factors treatment and strata for 0-10 days.
17MAR22,T05_mild

### Supplement 8

#### Full Table of Analysis of Variance for Days of Mild Symptoms
#### Comparison of Days with WURSS-44 Scores of 0-3 for All Symptoms
#### in Patients with Baseline WURSS-44 Q1 Score of 4-7

| | | | | | | | | Treatment Comparison | | | | |
|---|---|---|---|---|---|---|---|---|---|---|---|---|
| | | | | | | 95% c.i. | | Difference | | 95% c.i. | | |
| Questionnaire Item | Symptom Days | Treatment | N | LS Mean | Std Error | Lower | Upper | LS Mean | Std Error | Lower | Upper | P-Value |
| 34: Think clearly | 0-5 | Treatment | 29 | 21.62 | 1.53 | 18.56 | 24.68 | | | | | |
| | | Control | 30 | 22.70 | 1.50 | 19.69 | 25.71 | | | | | |
| | | Difference | 59 | | | | | -1.08 | 2.14 | -5.37 | 3.21 | 0.616 |
| | 5-10 | Treatment | 15 | 22.20 | 2.31 | 17.43 | 26.97 | | | | | |
| | | Control | 11 | 15.45 | 2.70 | 9.88 | 21.03 | | | | | |
| | | Difference | 26 | | | | | 6.75 | 3.56 | -0.59 | 14.08 | 0.070 |
| | 0-10 | Treatment | 44 | 21.35 | 1.34 | 18.70 | 24.01 | | | | | |
| | | Control | 41 | 20.08 | 1.42 | 17.25 | 22.91 | | | | | |
| | | Difference | 85 | | | | | 1.28 | 1.87 | -2.45 | 5.00 | 0.498 |
| 35: Speak clearly | 0-5 | Treatment | 15 | 18.07 | 2.34 | 13.28 | 22.85 | | | | | |
| | | Control | 16 | 22.87 | 2.26 | 18.24 | 27.51 | | | | | |
| | | Difference | 31 | | | | | -4.81 | 3.25 | -11.47 | 1.85 | 0.150 |
| | 5-10 | Treatment | 9 | 25.44 | 2.67 | 19.71 | 31.18 | | | | | |
| | | Control | 7 | 17.57 | 3.03 | 11.07 | 24.07 | | | | | |
| | | Difference | 16 | | | | | 7.87 | 4.04 | -0.79 | 16.54 | 0.072 |
| | 0-10 | Treatment | 24 | 21.02 | 1.90 | 17.18 | 24.86 | | | | | |
| | | Control | 23 | 21.55 | 1.99 | 17.54 | 25.56 | | | | | |
| | | Difference | 47 | | | | | -0.53 | 2.68 | -5.94 | 4.87 | 0.843 |
| 36: Sleep well | 0-5 | Treatment | 39 | 22.59 | 1.16 | 20.28 | 24.90 | | | | | |
| | | Control | 36 | 21.22 | 1.21 | 18.82 | 23.63 | | | | | |
| | | Difference | 75 | | | | | 1.37 | 1.67 | -1.97 | 4.71 | 0.417 |
| | 5-10 | Treatment | 16 | 20.19 | 2.30 | 15.49 | 24.88 | | | | | |
| | | Control | 17 | 15.88 | 2.23 | 11.33 | 20.44 | | | | | |
| | | Difference | 33 | | | | | 4.31 | 3.21 | -2.24 | 10.85 | 0.189 |
| | 0-10 | Treatment | 55 | 21.08 | 1.12 | 18.87 | 23.29 | | | | | |
| | | Control | 53 | 18.81 | 1.12 | 16.59 | 21.04 | | | | | |
| | | Difference | 108 | | | | | 2.27 | 1.52 | -0.74 | 5.27 | 0.138 |

*p<0.05 by one-way ANOVA within symptom day strata (0-5, 6-10) and ANOVA with factors treatment and strata for 0-10 days.

17MAR22,T05_mild

### Supplement 8

#### Full Table of Analysis of Variance for Days of Mild Symptoms
#### Comparison of Days with WURSS-44 Scores of 0-3 for All Symptoms
#### in Patients with Baseline WURSS-44 Q1 Score of 4-7

| | | | | | | | | Treatment Comparison | | | | |
|---|---|---|---|---|---|---|---|---|---|---|---|---|
| | | | | | | 95% c.i. | | Difference | | 95% c.i. | | |
| Questionnaire Item | Symptom Days | Treatment | N | LS Mean | Std Error | Lower | Upper | LS Mean | Std Error | Lower | Upper | P-Value |
| 37: Breathe easily | 0-5 | Treatment | 35 | 21.31 | 1.33 | 18.65 | 23.98 | | | | | |
| | | Control | 29 | 22.48 | 1.46 | 19.56 | 25.41 | | | | | |
| | | Difference | 64 | | | | | -1.17 | 1.98 | -5.12 | 2.79 | 0.557 |
| | 5-10 | Treatment | 13 | 21.15 | 2.68 | 15.66 | 26.65 | | | | | |
| | | Control | 17 | 20.94 | 2.35 | 16.13 | 25.75 | | | | | |
| | | Difference | 30 | | | | | 0.21 | 3.57 | -7.09 | 7.52 | 0.953 |
| | 0-10 | Treatment | 48 | 21.07 | 1.29 | 18.50 | 23.63 | | | | | |
| | | Control | 46 | 21.80 | 1.27 | 19.28 | 24.32 | | | | | |
| | | Difference | 94 | | | | | -0.73 | 1.75 | -4.21 | 2.75 | 0.678 |
| 38: Walk, climb stairs, exercise | 0-5 | Treatment | 39 | 21.97 | 1.22 | 19.55 | 24.40 | | | | | |
| | | Control | 47 | 20.83 | 1.11 | 18.62 | 23.04 | | | | | |
| | | Difference | 86 | | | | | 1.14 | 1.65 | -2.14 | 4.43 | 0.490 |
| | 5-10 | Treatment | 20 | 20.15 | 2.15 | 15.79 | 24.51 | | | | | |
| | | Control | 19 | 18.42 | 2.21 | 13.95 | 22.89 | | | | | |
| | | Difference | 39 | | | | | 1.73 | 3.08 | -4.51 | 7.97 | 0.578 |
| | 0-10 | Treatment | 59 | 21.01 | 1.10 | 18.83 | 23.20 | | | | | |
| | | Control | 66 | 19.69 | 1.07 | 17.57 | 21.81 | | | | | |
| | | Difference | 125 | | | | | 1.33 | 1.48 | -1.60 | 4.26 | 0.371 |
| 39: Accomplish daily activities | 0-5 | Treatment | 45 | 22.00 | 1.13 | 19.75 | 24.25 | | | | | |
| | | Control | 45 | 19.96 | 1.13 | 17.70 | 22.21 | | | | | |
| | | Difference | 90 | | | | | 2.04 | 1.60 | -1.14 | 5.23 | 0.206 |
| | 5-10 | Treatment | 21 | 19.52 | 2.04 | 15.39 | 23.66 | | | | | |
| | | Control | 18 | 19.22 | 2.20 | 14.76 | 23.69 | | | | | |
| | | Difference | 39 | | | | | 0.30 | 3.00 | -5.78 | 6.39 | 0.921 |
| | 0-10 | Treatment | 66 | 20.91 | 1.04 | 18.85 | 22.97 | | | | | |
| | | Control | 63 | 19.39 | 1.08 | 17.26 | 21.53 | | | | | |
| | | Difference | 129 | | | | | 1.52 | 1.43 | -1.32 | 4.36 | 0.291 |

*p<0.05 by one-way ANOVA within symptom day strata (0-5, 6-10) and ANOVA with factors treatment and strata for 0-10 days.
17MAR22,T05_mild

### Supplement 8

#### Full Table of Analysis of Variance for Days of Mild Symptoms
#### Comparison of Days with WURSS-44 Scores of 0-3 for All Symptoms
#### in Patients with Baseline WURSS-44 Q1 Score of 4-7

| | | | | | | | | Treatment Comparison | | | | |
|---|---|---|---|---|---|---|---|---|---|---|---|---|
| | | | | | | 95% c.i. | | Difference | | 95% c.i. | | |
| Questionnaire Item | Symptom Days | Treatment | N | LS Mean | Std Error | Lower | Upper | LS Mean | Std Error | Lower | Upper | P-Value |
| 40: Work outside the home | 0-5 | Treatment | 41 | 20.37 | 1.31 | 17.77 | 22.96 | | | | | |
| | | Control | 47 | 18.28 | 1.22 | 15.85 | 20.70 | | | | | |
| | | Difference | 88 | | | | | 2.09 | 1.79 | -1.46 | 5.64 | 0.246 |
| | 5-10 | Treatment | 22 | 19.27 | 2.05 | 15.11 | 23.43 | | | | | |
| | | Control | 17 | 16.06 | 2.33 | 11.33 | 20.79 | | | | | |
| | | Difference | 39 | | | | | 3.21 | 3.11 | -3.09 | 9.51 | 0.308 |
| | 0-10 | Treatment | 63 | 19.74 | 1.13 | 17.51 | 21.97 | | | | | |
| | | Control | 64 | 17.31 | 1.16 | 15.01 | 19.61 | | | | | |
| | | Difference | 127 | | | | | 2.43 | 1.56 | -0.65 | 5.51 | 0.121 |
| 41: Work inside the home | 0-5 | Treatment | 41 | 21.34 | 1.32 | 18.72 | 23.96 | | | | | |
| | | Control | 45 | 20.64 | 1.26 | 18.14 | 23.14 | | | | | |
| | | Difference | 86 | | | | | 0.70 | 1.82 | -2.92 | 4.32 | 0.703 |
| | 5-10 | Treatment | 20 | 19.15 | 2.08 | 14.93 | 23.37 | | | | | |
| | | Control | 19 | 19.47 | 2.14 | 15.14 | 23.80 | | | | | |
| | | Difference | 39 | | | | | -0.32 | 2.98 | -6.37 | 5.72 | 0.914 |
| | 0-10 | Treatment | 61 | 20.33 | 1.15 | 18.06 | 22.61 | | | | | |
| | | Control | 64 | 19.96 | 1.14 | 17.70 | 22.21 | | | | | |
| | | Difference | 125 | | | | | 0.38 | 1.55 | -2.70 | 3.45 | 0.808 |
| 42: Interact with others | 0-5 | Treatment | 43 | 21.16 | 1.28 | 18.63 | 23.70 | | | | | |
| | | Control | 46 | 19.13 | 1.23 | 16.68 | 21.58 | | | | | |
| | | Difference | 89 | | | | | 2.03 | 1.77 | -1.50 | 5.56 | 0.255 |
| | 5-10 | Treatment | 20 | 20.10 | 1.99 | 16.06 | 24.14 | | | | | |
| | | Control | 18 | 18.22 | 2.10 | 13.96 | 22.48 | | | | | |
| | | Difference | 38 | | | | | 1.88 | 2.90 | -3.99 | 7.75 | 0.521 |
| | 0-10 | Treatment | 63 | 20.65 | 1.11 | 18.44 | 22.85 | | | | | |
| | | Control | 64 | 18.66 | 1.12 | 16.44 | 20.88 | | | | | |
| | | Difference | 127 | | | | | 1.99 | 1.51 | -1.00 | 4.97 | 0.191 |

*p<0.05 by one-way ANOVA within symptom day strata (0-5, 6-10) and ANOVA with factors treatment and strata for 0-10 days.
17MAR22,T05_mild

### Supplement 8

#### Full Table of Analysis of Variance for Days of Mild Symptoms
#### Comparison of Days with WURSS-44 Scores of 0-3 for All Symptoms
#### in Patients with Baseline WURSS-44 Q1 Score of 4-7

| | | | | | | | | Treatment Comparison | | | | |
|---|---|---|---|---|---|---|---|---|---|---|---|---|
| | | | | | | 95% c.i. | | Difference | | 95% c.i. | | |
| Questionnaire Item | Symptom Days | Treatment | N | LS Mean | Std Error | Lower | Upper | LS Mean | Std Error | Lower | Upper | P-Value |
| 43: Live your personal life | 0-5 | Treatment | 45 | 20.16 | 1.24 | 17.70 | 22.61 | | | | | |
| | | Control | 51 | 19.02 | 1.16 | 16.71 | 21.33 | | | | | |
| | | Difference | 96 | | | | | 1.14 | 1.70 | -2.24 | 4.51 | 0.505 |
| | 5-10 | Treatment | 20 | 17.75 | 1.94 | 13.82 | 21.68 | | | | | |
| | | Control | 20 | 19.45 | 1.94 | 15.52 | 23.38 | | | | | |
| | | Difference | 40 | | | | | -1.70 | 2.75 | -7.26 | 3.86 | 0.540 |
| | 0-10 | Treatment | 65 | 19.23 | 1.09 | 17.08 | 21.38 | | | | | |
| | | Control | 71 | 18.93 | 1.06 | 16.84 | 21.02 | | | | | |
| | | Difference | 136 | | | | | 0.30 | 1.44 | -2.56 | 3.16 | 0.836 |

*p<0.05 by one-way ANOVA within symptom day strata (0-5, 6-10) and ANOVA with factors treatment and strata for 0-10 days.
17MAR22,T05_mild
