## Supplementary material for "Home-use Photobiomodulation Device Treatment Outcomes for COVID-19": Oxygen Saturation Analysis

### Supplement 9

### Oxygen Saturation Analysis

**Table of Contents**

9.1 Summary of Oxygen Saturation Analysis

9.2 Mixed Model Repeated Measures Analysis of Covariance Treatment Comparison of Oxygen Saturation (%) in in All Patients with Oxygen Saturation Data from Enrollment

9.3 Chart of Change in Oxygen Levels with Standard Error Bars

### Supplement 9.1

#### Summary of Oxygen Saturation Analysis

##### Introduction

We used the Mixed Model of Repeated Measures Analysis of Covariance (MMRM) model to analyze the oxygen saturation levels (SpO2) on the change from baseline in oxygen saturation (%) on days 7, 14, 21 and 28. Model terms include treatment, day, treatment-by-day interaction, symptom duration days and Baseline/Screening covariate.

All available data in percentage changes in SpO2 levels were read from Enrollment (n=267) as part of the safety monitoring. The loss of some patients who improved by Day 1 of treatment, and not considered for efficacy analyses, had no bearing on this exercise Least-square (LS) means for Treatment and Control and the differences between treatments included the 95% confidence interval (CI).

##### Results

Supplement 9.2 presents MMRM comparison of SpO2 percentage in the change from Enrollment in all patients with SpO2 data. Among 232 out of 267 patients with available SpO2 data, the LS mean difference of 0.32 % was statistically significant (P-value 0.018). in favor of the device.

The Chart on Changes in Oxygen Levels with Standard Error Bars is presented in Supplement 9.3.

##### Conclusion

In tracking SpO2 level changes of all patients from Enrollment, the Treatment group produced statistically significant (P=0.018) improvements.

#### Supplement 9.2

##### Mixed Model Repeated Measures Analysis of Covariance
##### Treatment Comparison of Oxygen Saturation (%) in the Change
##### in All Patients with Oxygen Saturation Data from Enrollment

| Treatment Estimate Change from Enrollment | | | | | | Treatment Comparison Change from Enrollment | | | | |
|---|---|---|---|---|---|---|---|---|---|---|
| | | | | 95% CI | | Difference | | 95% CI | | |
| Treatment | N | LS Mean | Std Error | Lower | Upper | LS Mean | Std Error | Lower | Upper | P-Value |
| Treatment | 117 | 0.54 | 0.098 | 0.35 | 0.73 | | | | | |
| Control | 115 | 0.22 | 0.1 | 0.02 | 0.41 | | | | | |
| Difference | 232 | | | | | 0.32 | 0.134 | 0.05 | 0.58 | 0.018* |

*p<0.05 by mixed model repeated measures analysis of covariance for the change from Enrollment in oxygen saturation (%) to days 7, 14, 21 and 28.

Model terms include treatment (Treatment, Control), day (7,14,21,28), treatment-by-day interaction, symptoms duration days on Enrollment (0-5, 6-10) and oxygen on Enrollment saturation. Least-squares (LS) mean, 95% confidence interval (CI) 04APR22,T08_o2sat_all

#### Supplement 9.3

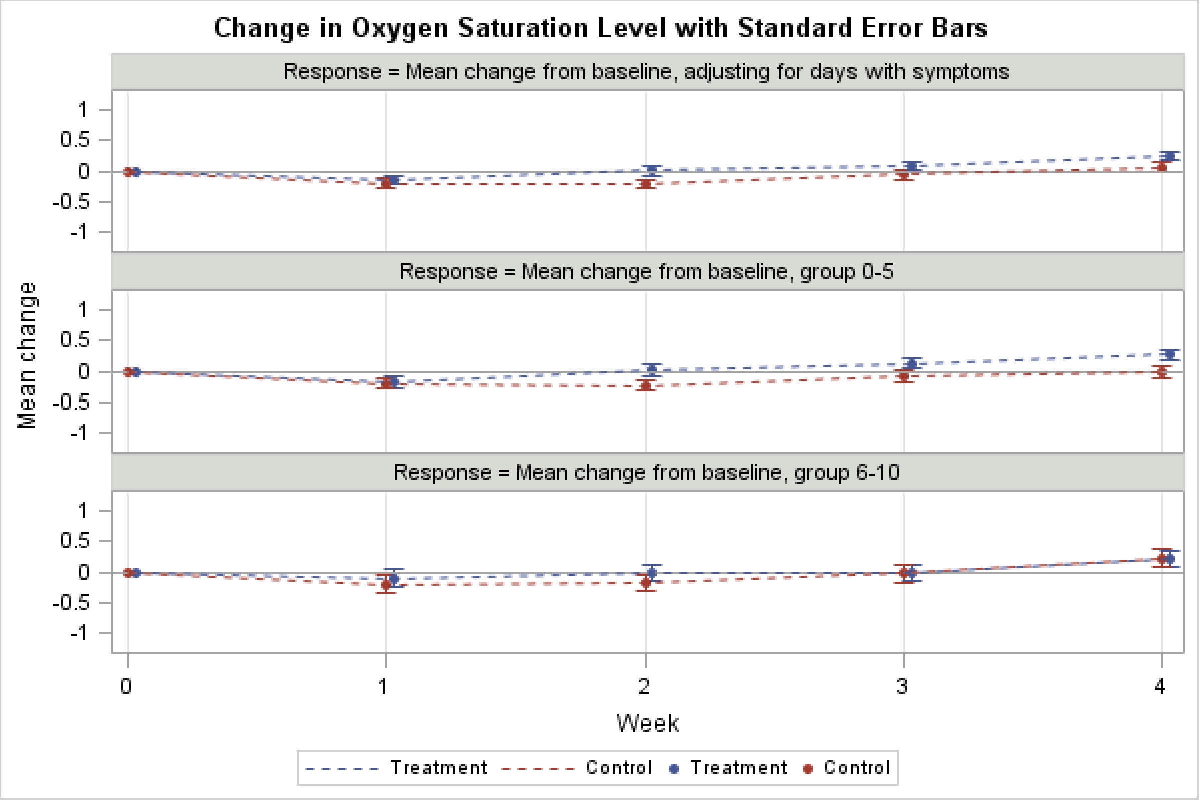
