## Supplementary material for "Home-use Photobiomodulation Device Treatment Outcomes for COVID-19": Adverse Events

### Supplement 10

### Adverse Events

**Table of Contents**

10.1 Notes on Adverse Events

10.2 Non-severe Adverse Events Affecting More than 5% of Patients

10.3 Severe Adverse Events Report by Medical Monitor

### Supplement 10.1

#### Notes on Adverse Events

Safety and adverse events were monitored for all patients upon enrollment (n=267) and over 30 days of assessment. Some improved before the baseline for efficacy analysis, hence excluded for efficacy analysis but this had no bearing on remaining eligible for adverse events monitoring.

Three tables summarizing adverse events are reported as laid out in the Statistical Analysis Plan (Supplement 3.2 as amended in Supplement 3.3):

1 All-cause Mortality: A table of all anticipated and unanticipated deaths, listing the diagnoses on expiration.

2 Serious Adverse Events requiring Hospitalization (Excluding Deaths): A table of all anticipated and unanticipated listing of patients, citing the main symptoms.

3 Non-serious Adverse Events: A table of anticipated and unanticipated listing of patients that exceed 5% of total of monitored patients.

**Table 1. All-cause Mortality**

| No of patients | Treatment or Control[1] | Diagnoses on expiration |
|---|---|---|
| 1 | Control | - septic shock<br>- COVID-19 pneumonia<br>- bilateral pneumonia<br>- hypoxic respiratory failure<br>- left-sided pneumothorax<br>- severe subcutaneous emphysema<br>- Type 2 diabetes<br>- (history of) hypertension<br>- hyperlipidemia<br>- history of coronary artery disease (post 2 stents placement to the left anterior descending artery)<br>- sick sinus syndrome (post permanent pacemaker placement)<br>- moderate protein-calorie malnutrition due to acute illness<br>- acute kidney injury, likely due to acute tubular necrosis due to septic shock |

**Table 2. Serious Adverse Events Requiring Hospitalization (Excluding Deaths)**

| No of patients | Treatment or Control[1] | Diagnosis |
|---|---|---|
| 1 | Control | COVID pneumonia |
| 1 | Control | Respiratory decomposition |
| 1 | Control | Unspecified COVID symptoms |

#### Table 3. Adverse Events Not Requiring Hospitalization[2 3 4 5]

| | Monitored from Enrollment | | | | | | | | | |
|---|---|---|---|---|---|---|---|---|---|---|
| | Treatment Group, N=135 | | | Control Group, N=132 | | | Differences in % of Patients, Treatment-Control | | | |
| | | | | | | | | 95% CI | | |
| Adverse Event | Number of Patients | % in Treatment | Number of Events | Number of Patients | % in Control | Number of Events | Difference in % | Lower | Upper | P-value |
| **Significant Symptom** | | | | | | | | | | |
| Tachycardia | 40 | 29.63 | 107 | 58 | 43.94 | 180 | -14.30 | -25.90 | -1.87 | 0.016 |
| Dysgeusia | 7 | 5.19 | 7 | 24 | 18.18 | 24 | -13.00 | -21.20 | -4.63 | 0.001 |
| **Other Symptoms >5% of Patients** | | | | | | | | | | |
| Bradycardia | 38 | 28.15 | 100 | 38 | 28.79 | 102 | -0.64 | -11.60 | 10.51 | 0.940 |
| Diarhea | 19 | 14.07 | 23 | 24 | 18.18 | 27 | -4.11 | -13.30 | 4.89 | 0.529 |
| Nausea | 16 | 11.85 | 18 | 27 | 20.45 | 31 | -8.60 | -17.90 | 0.36 | 0.058 |
| Anosmia/parosmia | 11 | 8.15 | 11 | 15 | 14.35 | 19 | -6.25 | -14.30 | 1.55 | 0.126 |
| Emesis | 8 | 5.93 | 8 | 10 | 7.58 | 12 | -1.65 | -8.28 | 4.75 | 0.683 |

Notes:

1. Events in Control would be unrelated to the investigational agent.

2. "Adverse Events Not Requiring Hospitalization" collectively include mild and moderate levels of adverse events not requiring hospitalization.

3. Symptoms affecting >5% of monitored patients are listed. The full table is shown in Supplement 4.

4. Patients were monitored for safety and adverse events from the time of Enrollment and after initial Follow-up. As the result 267 were monitored, 135 in Treatment and 132 in Control.

5. Upon Enrollment all these patients qualified with WURSS-44 severity scores of 4-7. They continued to be monitored throughout their 30-day assessment period although some would fail to be included in the Baseline for Final Analysis due to improvements.

---

These are presented as Table 4 in the main manuscript.

Severe Adverse Events (SAEs) as reported by the Medical Monitor is set out in Supplement 10.5.

In summary, there were four cases requiring hospitalization, including one death. All the SAEs occurred in the Control group, and none was reported in the Treatment group. The conditions of three patients deteriorated rapidly after Enrollment and before Baseline; from moderate-to-severe (WURSS-44 Q1 score of 4-7) to SAEs requiring hospitalization, one of whom died.

#### Supplement 10.2 Non-severe Adverse Events Affecting More than 5% of Patients

| | Monitored from Enrollment | | | | | | | | | |
|---|---|---|---|---|---|---|---|---|---|---|
| | Treatment Group, N=135 | | | Control Group, N=132 | | | Differences in % of Patients, Treatment-Control | | | |
| | | | | | | | | 95% CI | | |
| Adverse Event | Number of Patients | % in Treatment | Number of Events | Number of Patients | % in Control | Number of Events | Difference in % | Lower | Upper | P-value |
| **Significant Symptom** | | | | | | | | | | |
| Tachycardia | 40 | 29.63 | 107 | 58 | 43.94 | 180 | -14.30 | -25.90 | -1.87 | 0.016 |
| Dysgeusia | 7 | 5.19 | 7 | 24 | 18.18 | 24 | -13.00 | -21.20 | -4.63 | 0.001 |
| **Other Symptoms >5% of Patients** | | | | | | | | | | |
| Bradycardia | 38 | 28.15 | 100 | 38 | 28.79 | 102 | -0.64 | -11.60 | 10.51 | 0.940 |
| Diarhea | 19 | 14.07 | 23 | 24 | 18.18 | 27 | -4.11 | -13.30 | 4.89 | 0.529 |
| Nausea | 16 | 11.85 | 18 | 27 | 20.45 | 31 | -8.60 | -17.90 | 0.36 | 0.058 |
| Anosmia/parosmia | 11 | 8.15 | 11 | 15 | 14.35 | 19 | -6.25 | -14.30 | 1.55 | 0.126 |
| Emesis | 8 | 5.93 | 8 | 10 | 7.58 | 12 | -1.65 | -8.28 | 4.75 | 0.683 |

#### A Randomized Study Evaluating the Efficacy of the Vielight RX in the Treatment of COVID-19 Respiratory Symptoms

##### Medical Monitor Safety Summary

Interim adverse experience (AE) reports were reviewed from October 28, 2020 through July 20, 2021 during the conduct of the clinical trial "A Randomized Study Evaluating the Efficacy of the Vielight RX in the Treatment of COVID-19 Respiratory Symptoms." Serious adverse event (SAE) reports were also reviewed during this period as the reports became available. A final summary of AE events was received on September 12, 2021.

The clinical trial enrolled patients with coronavirus disease 2019 (COVID-19) and was an open label trial. Subjects were randomized as untreated controls or receiving treatment with Vielight RX.

There were 4 SAEs reviewed during the study. All of these SAEs represented hospitalization, and there was 1 death reported. All SAEs were attributed to COVID-19. All of the SAEs were in the control group. The subject that died was in the control group. Due to the potential severity of COVID-19, hospitalizations and deaths were expected. No SAEs were attributed to study participation, and this was confirmed by independent review. Since these events were not unexpected and not attributed to study participation, study continuation was advised.

The occurrence of at least 1 AE was reported for 209 study subjects. The vast majority of the AEs were reported as mild, with a few moderate AEs also reported. There was a large number of objective reports of tachycardia and bradycardia that arose from pulse oximeter readouts. These readouts met numerical criteria for tachycardia (over 100 beats per minute) or bradycardia (less than 60 beats per minute), and the duration of these events were largely indeterminate due to the nature of the data collection. These objective events appeared to be equally distributed between subjects in the control and treatment groups. This was expected, as bradycardia and tachycardia are known to be associated with COVID-19. Subjective reports of high or low heart rate were rare. The high number of tachycardia and bradycardia reports were thought to be an artifact of the pulse oximetry readouts, and were either spurious findings or attributed to COVID-19.

Gastrointestinal AEs (nausea, diarrhea) were reported in both control and treated subjects at an expected frequency, and there were no trends that suggested that these AEs were associated with study treatment. Similarly, AEs attributable to COVID-19, such as dysgeusia and hyposmia, occurred with approximately equal frequency in both control and treated subjects, and there were no trends that suggested that these AEs were associated with study treatment.

There were 5 AEs that appeared to be related to study treatment: erythema on the chest (11 treatment subjects); headache (6 treatment subjects); irritation of the nostril (5 treated subjects); eye discomfort (4 treated subjects); and rhinorrhea (4 treated subjects). The timing of the events, as well as the location of the events relative to the treatment application (e.g. eye discomfort on the same side as the treatment application) suggest an association between these AEs and treatment with Vielight RX. It is important to note that all of these AEs were reported as mild.

In summary, there were no safety signals or trends that suggested that study treatment or study participation represented an undue risk to study participants. Reported SAEs were expected in terms of their nature and frequency and were unrelated to study participation. The high number of reports of tachycardia and bradycardia appear to be a result of reporting due to pulse oximetry readouts. While some of the common expected AEs did not appear to be attributable to study treatment, 5 AEs (chest erythema, headache, nostril irritation, rhinorrhea, and eye discomfort) appear to be associated with the application of study treatment. Almost all AEs were mild. Overall, treatment with Vielight RX appears to be safe and well tolerated.

Russell G Clayton DO 13 MAR 2022

Russell G. Clayton Sr., D.O.
Medical Monitor

Date
